# Five-Decade Prevalence of Delirium in Pneumonia, Risk Factors, and Associated Mortality: A Systematic Review and Meta-Analysis

**DOI:** 10.1101/2025.06.01.25328725

**Authors:** Erika L. Juarez-Martinez, Aida Araia, Dillan Prasad, Shreya Dhar, Khizar Nandoliya, Ian G. Sherrington, Catherine Zhao, Annie Wescott, Chiagozie I. Pickens, Richard G. Wunderink, Eyal Y. Kimchi

**Affiliations:** Ken & Ruth Davee Department of Neurology, Feinberg School of Medicine, Northwestern University. Chicago, Illinois, 60611; Galter Health Sciences Library & Learning Center, Feinberg School of Medicine, Northwestern University, Chicago, IL, 60611; Division of Pulmonary and Critical Care, Department of Medicine. Feinberg School of Medicine. Northwestern University. Chicago, Illinois, 60611

**Keywords:** pneumonia, hospitalization, delirium, encephalopathy, systematic review

## Abstract

**Background:** Delirium can occur in patients with pneumonia, but its prevalence is inconsistent across studies. Unreliable estimates and uncertainty regarding the significance of patient-specific vs. microbiological risk factors hinder delirium management and prognosis. Here, we provide robust estimates of delirium prevalence in patients with pneumonia, associated risk factors, and association with mortality.

**Methods:** We searched five databases (MEDLINE, Cochrane Library, Embase, PsycINFO, and Scopus), from inception to August 6, 2024. We included studies in adults hospitalized with pneumonia reporting delirium, encephalopathy, or altered mental status. Two investigators extracted data and assessed risk of bias. Summary rates were calculated using random-effects models. We performed prespecified analyses for diagnostic methods, microbiologic factors, clinical factors, and mortality, with sensitivity analysis among studies at low risk of bias. Registration: PROSPERO-CRD42023385571.

**Results:** Delirium prevalence across 126 studies was 22% (95% CI [18%–26%]), and higher in studies at low risk of bias (40% [24%–58%], n=11). Standardized assessments yielded higher rates than symptom-or ICD code-based assessments (p<0.05). Surprisingly, delirium rates did not differ by microbiological etiology (p*=*0.63), including COVID-19, nor by pneumonia origin (p=0.14). Predisposing factors included older age and neurologic and systemic comorbidities. Delirium was associated with increased mortality (OR 4.3 [3.24–5.76], p<0.001), without change over five decades (p = 0.32).

**Interpretation:** Delirium is highly prevalent and enduring in pneumonia. Our results emphasize patient-and care-related factors over microbiological causes, including COVID-19. Delirium’s entrenched association with mortality, even considering covariates, reinforces the need to manage delirium as a convergent syndrome in pneumonia.

**Take-home message (shareable abstract):** Delirium rates in pneumonia vary widely across studies. This meta-analysis establishes that delirium is common in pneumonia, driven by patient and care related factors rather than microbiology including COVID-19, and consistently associated with mortality.

## Introduction

Pneumonia is one of the most frequent causes of hospitalization in the USA and the leading cause of infectious mortality worldwide.^1,2^ Delirium, an acute neuropsychiatric disorder characterized by disturbances in attention and awareness, is variably observed in patients hospitalized with pneumonia and has been associated with increased length of hospital stay and mortality.^3–6^ To date, there is no consensus on delirium prevalence in patients with pneumonia across studies, with reported rates from lower than 10% to as high as 80%, raising concerns for potential under-or over-diagnosis.^7–11^ Furthermore, the importance of key risk factors remains largely undetermined, given delirium’s multifactorial origin and the complex interplay between infection-specific vs. patient-specific risk factors. This lack of consensus hampers efforts to anticipate, prevent, and manage delirium effectively in patients with pneumonia.

In general, older age, cognitive impairment, and severe illness are known risk factors for delirium across many different clinical conditions.^12^ But for pneumonia specifically, infection-related factors such as the setting in which pneumonia is acquired, microbiological origin, and the extent of lung involvement, may play important roles. Notably, during the COVID-19 pandemic, several studies reported increased rates of delirium in severe COVID-19 infections, suggesting a microbiology-related contribution. ^13,14^ However, comparisons of delirium rates between COVID-19 and non-COVID pneumonia etiologies are wanting.

This gap could result in an underappreciation of delirium in non-COVID pneumonia populations.

Establishing reliable prevalence estimates of delirium in patients hospitalized with pneumonia and identifying the key factors influencing its occurrence and recognition are essential to guide healthcare providers in implementing targeted, multimodal strategies within the broader care of patients with pneumonia, to reduce delirium incidence, improve prognosis, and mitigate mortality. This systematic review and meta-analysis addresses this knowledge gap by quantifying delirium prevalence across a wide population range, hospital settings and time periods, including pre-and post-COVID era, evaluates the relevance of associated predisposing and precipitating risk factors for both infection and patient features, and robustly determines its association with mortality over time, even when accounting for other important clinical covariates.

## Methods

We followed the Preferred Reporting in Systematic Review and Meta-Analysis guidance (PRIMSA).^15^ The protocol was registered in PROSPERO, the international prospective register of systematic reviews (CRD42023385571).

### Search strategy and eligibility criteria

We searched five databases: MEDLINE, Cochrane Library, Embase, PsycINFO, and Scopus, without filters or language limits, from inception to August 2024. We used controlled vocabulary and keywords related to the topics of (1) pneumonia, (2) delirium, encephalopathy, altered mental status, or confusion, and (3) hospitalization. Multiple terms were used for delirium given fragmentation of nomenclature in the clinical literature.^16,17^ References were downloaded and deduplicated, and unique records were uploaded to Covidence for screening. Search strategies are detailed in Supplementary Table S1 and the numbers of studies initially obtained per database are detailed in Supplementary Table S2.

Our preregistered inclusion criteria were studies describing adult patients (>18 years old) hospitalized with pneumonia reporting rates of delirium or related keywords. We only included studies from peer-reviewed journals. We *a priori* excluded studies with <20 patients with pneumonia, delirium tremens, chronic or non-delirium-related encephalopathies, and case-control or randomized-controlled designs, as in the former, the prevalence of delirium cannot be estimated from preselected populations, while in the latter intervention studies, rates of delirium may be less reflective of natural clinical progression. Two reviewers independently screened records at each stage and discrepancies were resolved through consensus. PRISMA study flow diagram is shown in Figure 1.

**Figure 1.**
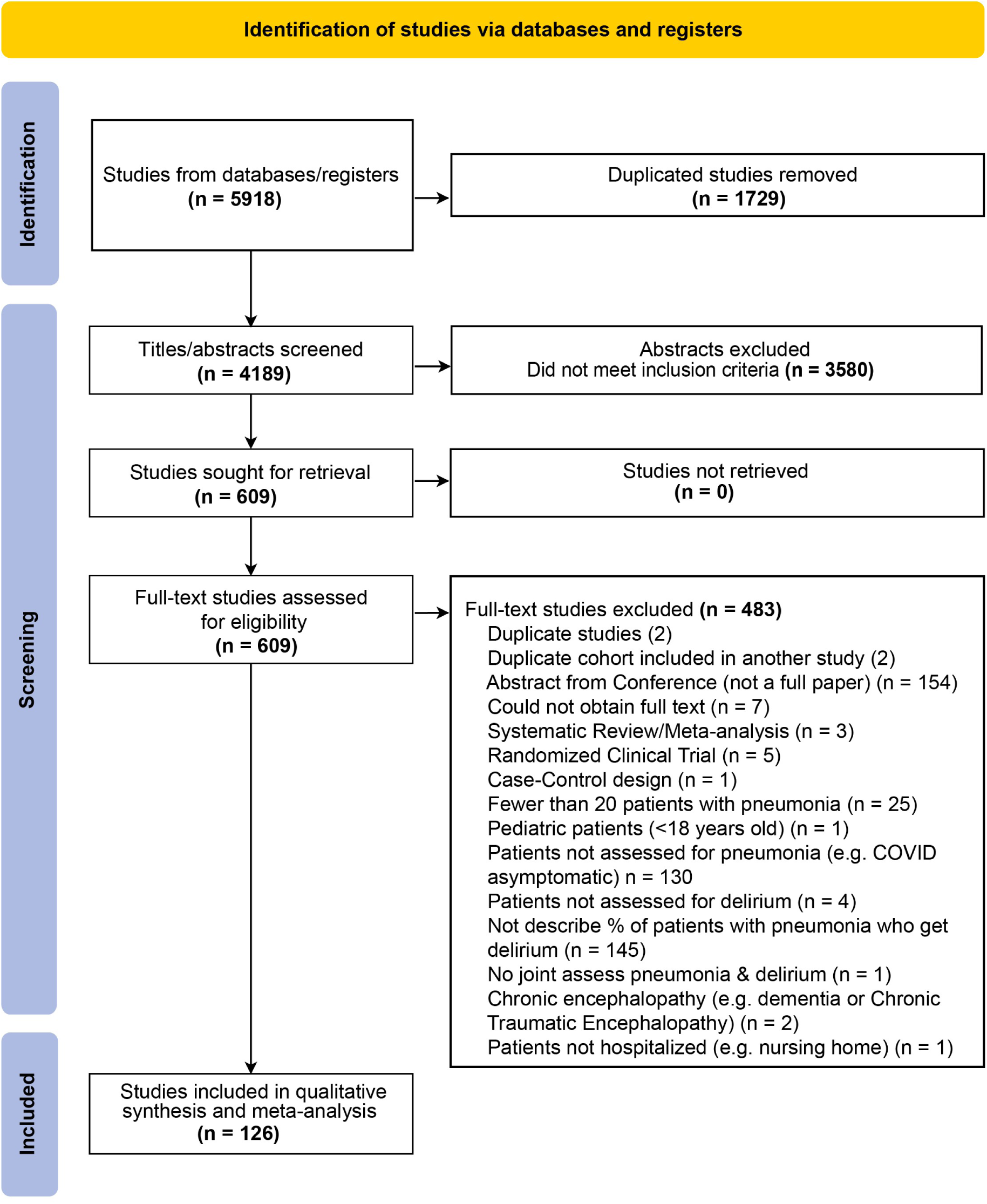
Preferred Reporting in Systematic Review and Meta-Analysis (PRISMA) Flowchart

### Data extraction and management

Two authors independently extracted data from included studies (E.J.M., D.P., or E.Y.K.). Discrepancies were resolved through consensus. Extracted data fields are presented in the Supplementary Methods and summarized here: general study information; population demographics and comorbidities;^18^ pneumonia diagnostic criteria, imaging, infection origin (e.g., community vs. hospital-acquired), and microbiological etiology; clinical severity indicators (CURB-65 scores,^19^ Pneumonia severity index (PSI) scores,^20^ mechanical ventilation, ICU admission); delirium/encephalopathy assessment methods (Diagnostic and Statistics Manual-based assessment (DSM),^21^ validated delirium scales (e.g., the confusion assessment method framework, CAM^22^ or CAM-ICU^23^); non-delirium standardized assessments of mental status (e.g. Glasgow Coma Scale, GCS),^24^ International Classification of Diseases (ICD) codes, and non-structured, symptom-based reports (e.g. tracking of individual symptoms collection such as confusion, altered mental status, or altered consciousness); and clinical outcomes (length of hospitalization and stay in the ICU, length of ventilation, and mortality). For mortality, we recorded odds ratios from univariate analysis, as well as odds ratios from multivariable analyses when performed while controlling for clinical covariates.

### Meta-analysis methods

Meta-analyses were performed in R (4.2.3) using the *metafor* package.^25^ All primary analyses included all eligible studies and were performed at the study level, since individual patient data were not available across this broad set of studies. We used random-effects models for summary estimates of delirium prevalence given expected heterogeneity. The restricted maximum likelihood estimator (REML)^26^ was used to calculate the heterogeneity variance τ^2^. We also assessed between-study heterogeneity using Higgin I^2^, since it is robust to changes in the number of studies in the analysis, unlike Cochrane’s Q. We used Knapp-Hartung adjustments to calculate the confidence interval around the pooled effect.^27^ Effect measures were calculated using the *metaprop*, *metabin*, *metacont* functions and the *metamedian*^28^ package (Hozo/Wan/Bland method), and were expressed as proportions, odds ratios, and difference of means (DOM), respectively. Proportions were logit-transformed automatically in R before they were pooled. We created forest plots of all estimates with 95% confidence intervals (CI). The contribution of each study to the pooled proportion was given by its weight based on the inverse variance of the study’s proportion estimate. Publication bias was assessed using funnel plots^29^ and supplemented with Eggers’ regression test. The effects of publication bias were assessed using Rucker’s limit meta-analysis method (*metasens* package).^30^

We performed prespecified subgroup analyses based on infection origin, microbiological agent, clinical care setting, and delirium assessment method, including all studies that reported relevant data to minimize potential selection bias. Meta-regression using random-effects models was used to calculate R^2^, i.e., the proportion of variance in the dependent variable (delirium rate) explained by each model, and subgroup effects. Post-hoc comparisons were performed to test for subgroup differences compared to a gold standard subgroup when applicable, or otherwise to the average proportion across all groups. *p-*values were adjusted for multiple comparisons using Holm’s procedure. We ran separate analyses for studies that split cohorts into groups <65 and>65 years old, and cohorts that assessed delirium in groups based on gender, comorbidities, pneumonia severity factors, or clinical care measures. *p-*values *<* 0.05 were considered significant and corrected for multiple comparisons where indicated.

### Quality and risk of bias assessment

Risk of bias was assessed by two independent reviewers using the JBI manual for evidence synthesis tool (Supplementary Table S3).^31^ Disagreements were resolved through consensus. The overall risk of bias was classified as “low” if studies met ≥6/8 domains, including validated evaluation of both pneumonia and delirium. We conducted sensitivity analyses in studies at low risk of bias.

## Results

### Included Studies

We identified 4,189 articles after deduplication, of which 126 met inclusion criteria and were used for synthesis and meta-analysis (Figure 1). These studies represent a total of 8,379,648 patients with pneumonia and included cohorts from Africa, Asia, Europe, North America, and South America. Paper languages included English (n =113), Spanish (n=7), Chinese (n=2), Dutch (n=1), German (n=1), Japanese (n=1), and Portuguese (n=1). Study characteristics are shown in Supplementary Table S4 and S5, and risk of bias assessments in Supplementary Figure S1.

### Summary Prevalence

The summary meta-analytic estimate for delirium was 22% (95% CI [18%; 26%], Figure 2). There was statistical heterogeneity in the results (I^2^ = 100%, T^2^ = 1.62 [1.4; 2.4]), which was expected and explored below.^29^ Funnel plot analysis suggested the presence of true heterogeneity rather than non-reporting bias (Supplementary Figure S2), despite a suggestive Eggers’ regression test^32^ (test 22.2 [18.3 – 26.1], t = 11.1, p<0.0001). Regardless, addressing publication bias using Rucker’s limit meta-analysis method yielded a similar adjusted proportion delirium occurrence estimate of 19% [16%; 23%].

**Figure 2.**
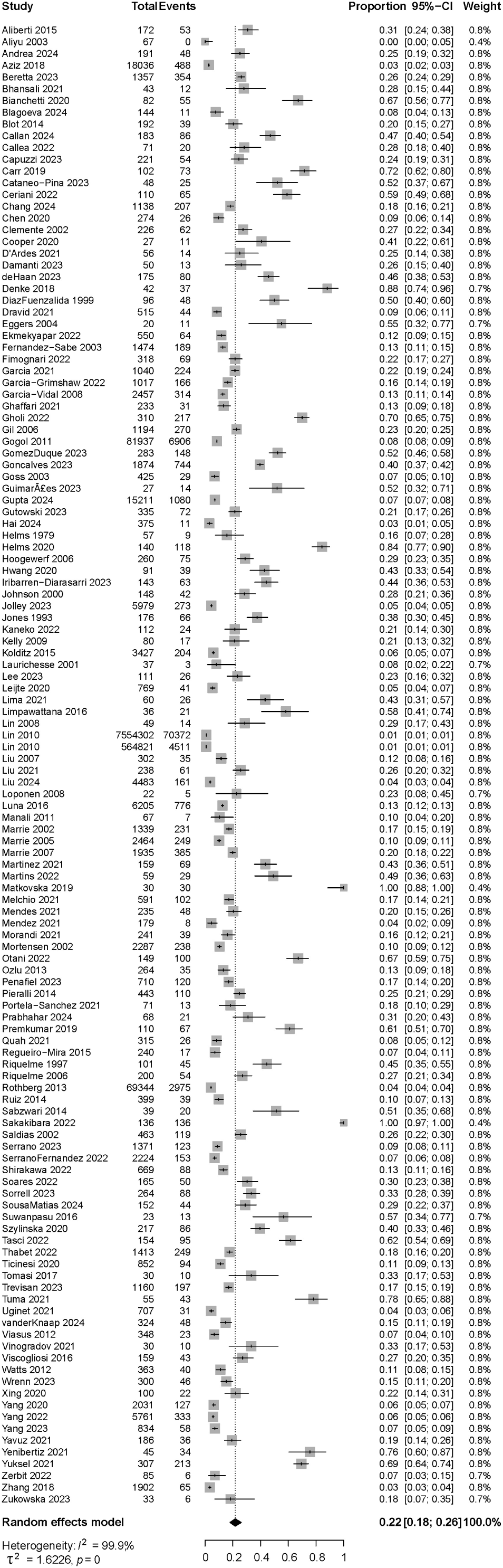
Summary estimate for occurrence of delirium in patients hospitalized with pneumonia. Effect measures were calculated using random-effects models and expressed as proportions. 126 studies were included, and the overall summary rate was 0.22 (95% CI [0.18-0.26]).

The quality of the studies (risk of bias assessment) influenced delirium rates, explaining 6.2% of delirium rates variance (R^2^ = 6.21%, F_2,123_ = 4.35, p=0.015). Eleven studies had a low risk of bias, including both validated pneumonia and delirium assessment (2,517 patients). In those studies, the reported occurrence of delirium was 40% [24%; 58%] and higher compared to those at moderate risk (n=44 studies, 16% [12%; 20%], p=0.005) and high risk of bias (n=71 studies, 24% [18%; 31%], p=0.0002). Forest plots per risk of bias group with delirium rates, 95% CI and measures of heterogeneity are provided in Supplementary Figure S3.

### Delirium Assessment

Given the heterogeneity across studies, we investigated factors that may contribute to reported variability in delirium rates (Supplementary Figures S4–S7). Delirium rates were significantly influenced by assessment method (Figure 3A, R^2^ = 36.7%, F_4,120_ = 15.2, p<0.0001). Compared to gold standard DSM diagnosis (n=12 studies, 34% [24%, 45%]), delirium rates were similar when studies used validated delirium scales (n=20 studies, 40% [28%; 53%], p=0.73 compared to DSM) or standardized assessments of mental status (AMS scales, n=7 studies, 45% [24%; 69%], p=0.73 compared to DSM). In contrast, compared to DSM diagnosis, delirium rates were lower when studies used non-standardized symptom assessments (symptom collection, n=78 studies, 18% [15%; 22%], p=0.03) or ICD codes (n=8 studies, 3% [2%; 6%], p<0.001). Forest plots for each subgroup analysis, including delirium rates and 95% CI are shown in Supplementary Figure S4. Sensitivity analysis in studies at low risk of bias (in which ascertainment of delirium through validated methods was a criterion) showed similar results, with no difference in delirium rates when assessed using the DSM (n=3 studies, 30% [9%; 65%]), standardized assessments of mental status (AMS Scales) (n=2 studies, 59% [18%; 91%], and validated delirium scales (n=6 studies, 39% [13%; 74%]) (Supplementary Figure S8A, R^2^ = 0.0%, F_2, 8_ = 0.54, p=0.60). Forest plot for the sensitivity analysis per delirium assessment method is shown in Supplementary Figure S9A.

**Figure 3.**
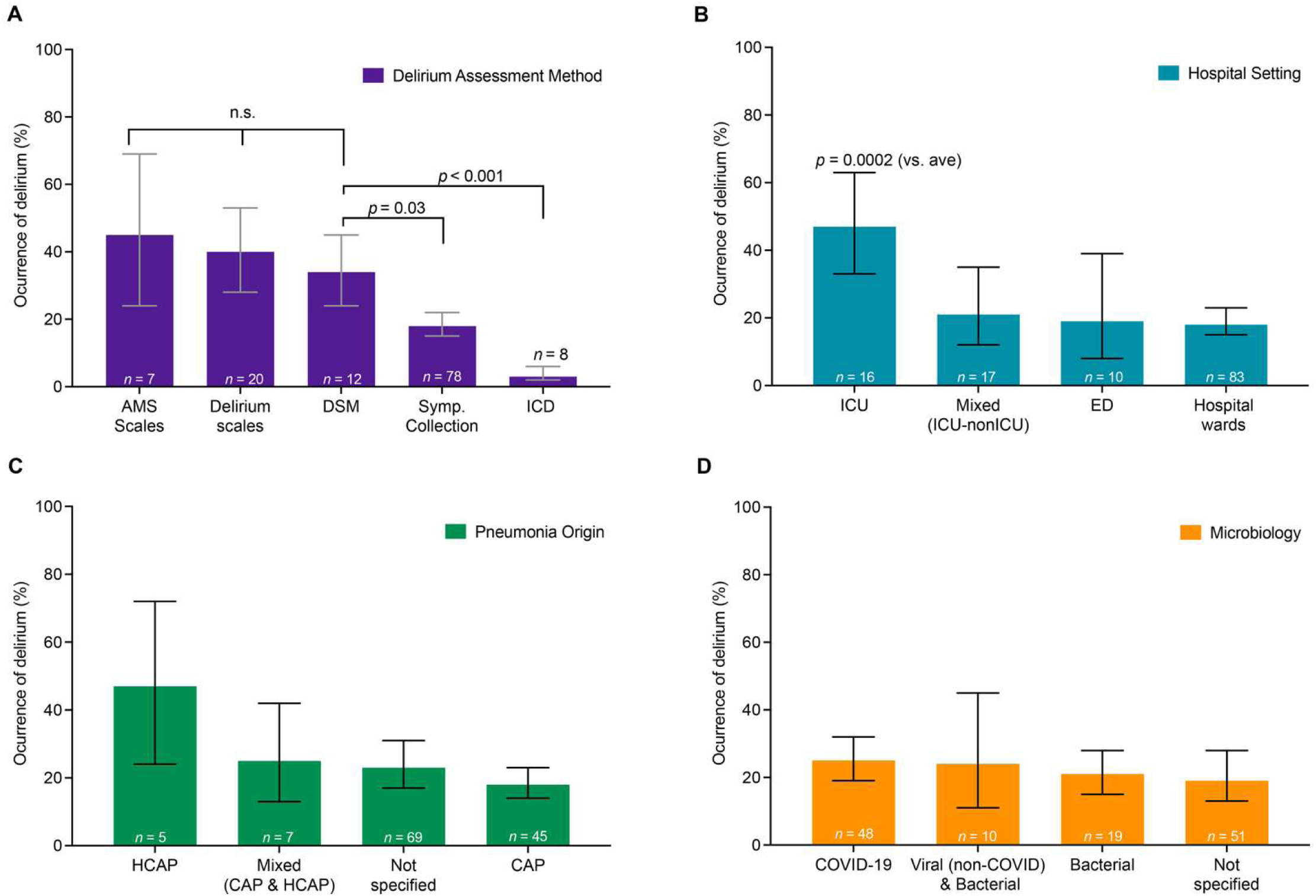
Delirium rates differ based on assessment methods and are higher in studies performed in intensive care units, but do not differ based on pneumonia origin or microbiological etiology. Delirium rates varied significantly according to the assessment method used, which explained 37% of variance in delirium rates across studies (R^2^ = 36.8%, *p*<0.0001). Given that the Diagnostic and Statistical Manual (DSM) assessment is considered the gold standard for delirium assessment, we compared delirium rates to those of the DSM. Delirium rates were similar to the DSM assessment for standardized assessments of mental status (AMS Scales, such as Glasgow Coma Scale or Richmond Agitation Sedation Scale, *p*=0.73) or validated Delirium scales (such as CAM, CAM-ICU, ICSDC, 4AT, see abbreviations in Supplementary eMethods, *p*=0.73). In contrast, rates of delirium were lower when assessed using nonstructured symptom assessments (i.e., symptom collection, *p=*0.03) or ICD codes (*p<*0.001). Symp. Collection = symptom collection: identification of relevant symptoms, e.g. chart report of altered mental status. Each bar represents a meta-analytic estimate of delirium rates, with the calculated 95% confidence interval. Post-hoc *p-*values in all panels are adjusted for multiple comparisons (Holm). Forest plots of all studies with n’s, meta-analytic proportion, CI, and heterogeneity measures for each subgroup analysis are provided in supplementary material (Supplementary Figure S4). **A.** Delirium rates varied significantly according to study setting, which explained 12% of variance in delirium rates across studies (R^2^ 12.1%, *p=*0.002). Compared to the average of all studies, delirium rates were significantly higher for studies performed in the ICU (*p=*0.0002). (ICU = Intensive Care Unit, ED = Emergency Department). Conventions as in A, with forest plots in Supplementary Figure S5). **B.** Delirium rates did not vary significantly according to pneumonia origin, which explained only 2.4% of variance in delirium rates across studies (R^2^ = 2.4%, *p*=0.15). (HCAP=Healthcare Acquired Pneumonia, CAP=Community Acquired Pneumonia). Conventions as in A, with forest plots in Supplementary Figure S6). **C.** Delirium rates did not vary significantly according to microbiological etiologies, which explained 0.0% of variance in delirium rates across studies (R^2^ = 0.0%, *p=*0.63). Conventions as in A, with forest plots in Supplementary Figure S7).

### Hospital Care Setting

The hospital care setting in which the study was conducted was significantly associated with delirium rate (R^2^ 12.1%, F_3,122_ = 5.2, p=0.002). Post-hoc comparisons showed that delirium occurrence was greater than average in studies conducted in the ICU (Figure 3B, n=16 studies, 47% [33%; 63%], p=0.0002). Forest plots for subgroup by hospital care setting, delirium rate and 95% CI are shown in Supplementary Figure S5. Sensitivity analysis in studies at low risk of bias showed similar results, with delirium rates varying significantly according to the setting and explaining 68% of the variance (R^2^ = 68.3, F_1, 9_ = 20.06, p=0.0015). Delirium rates were also higher in the ICU studies (n=3 studies, 73% [36%; 93%], p=0.001) than in hospital ward studies (Supplementary Figure 8B, n=8 studies, 27% [18%; 39%]). Forest plot for the sensitivity analysis per hospital study setting is shown in Supplementary Figure S9B.

### Microbiological Factors

Delirium rates did not differ significantly based on the origin of the infection (e.g., community-acquired vs. healthcare-acquired pneumonia) (Figure 3C, R^2^ = 2.4%, F_3,122_ = 1.8, p=0.146). Delirium rates also did not differ significantly based on the type of microbiological agent causing pneumonia, whether COVID-19, or other viral and/or bacterial causes (Figure 3D, R^2^ = 0.0%, F_3,124_ = 0.57, p=0.630). Forest plots, delirium rates, and 95% CI are shown for infection origin in Supplementary Figure S6 and for microbiological agents in Supplementary Figure S7. Sensitivity analysis in studies at low risk of bias showed similar results, with no difference in delirium rates based on infection origin (Supplementary Figure 8C, R^2^ = 0%, F_2, 8_ = 0.26, p=0.77), or the type of microbiological agent (Supplementary Figure 8D, R^2^ = 0%, F_3, 7_ = 0.04, p=0.98). Forest plot for the sensitivity analysis for infection origin are shown in Supplementary Figure S9C and for microbiological agents in Supplementary Figure S9D.

### Predisposing Factors

Next, we investigated potential predisposing factors for delirium (Table 1 and Supplementary Figures S10–S12). Patients with pneumonia and delirium were older than those without delirium (Supplementary Figure S10A, n=18 studies, difference of means (DOM) +6.5 years [4.2; 8.7], p<0.0001). Studies stratifying patients into those younger and older than 65 years (n=7 studies) found that age >65 was associated with delirium (Supplementary Figure S10B, OR 2.9 [2.4; 3.5], p<0.0001). Gender was not associated with delirium, but alcohol intake was (Supplementary Figure S11, n=3 studies, OR 2.1 [1.02; 4.16], p=0.047). Surprisingly, delirium was somewhat less likely in patients with a history of smoking (n=6 studies, OR 0.8 [0.7; 0.96], p=0.021).

**Table 1.**
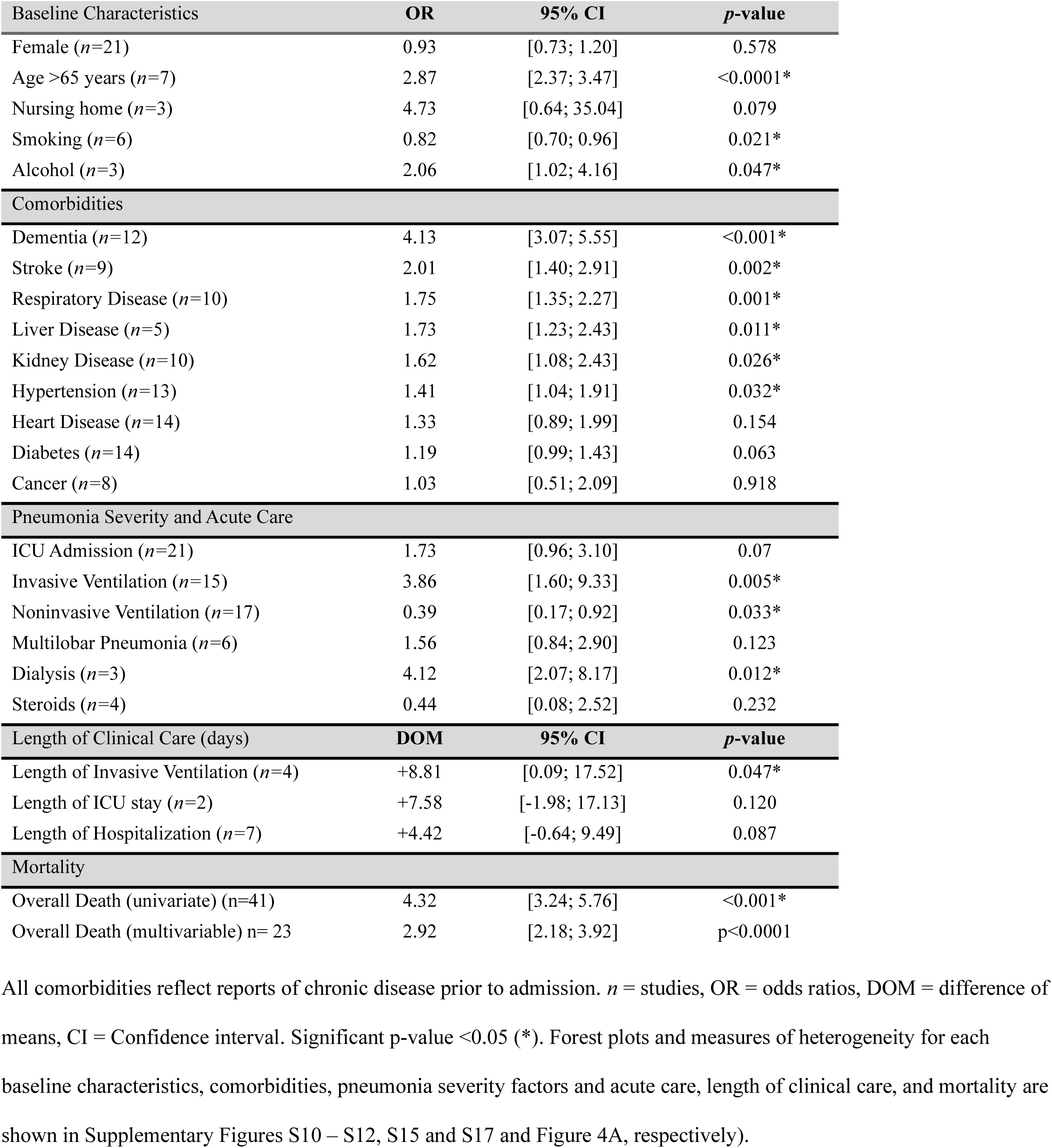
Relationship of delirium to clinical factors, including baseline characteristics, comorbidities, measures of pneumonia severity and acute care, mortality, and Length of Clinical Care.

Multiple comorbidities were associated with delirium (Table 1 and Supplementary Figure S12), including neurologic disorders such as dementia (OR 4.1, [3.1; 5.5], p<0.001) and stroke (OR 2.0 [1.4; 2.9], p=0.002), and chronic respiratory disease (COPD OR 1.7 [1.3; 2.3], p=0.001). Delirium was also associated with liver disease (OR=1.7 [1.2; 2.4], p=0.011), kidney disease (OR 1.6 [1.08; 2.43], p=0.026), and hypertension (OR 1.4 [1.04; 1.91], p=0.032). We found no association between delirium and diabetes, heart disease, or cancer (p<0.05).

Sensitivity analysis in studies at low risk of bias corroborated that patients with delirium were older than those without delirium (n=5 studies, DOM +6.2 years [1.5; 10.9], p=0.01) (Supplementary Table S6 with forest plots in Supplementary Figure S13). Neurological comorbidities remained associated with delirium such as dementia (n=3 studies, OR 4.1, [1.9; 8.9], p=0.015) and stroke (n=4 studies, OR 2.2 [1.3; 3.7], p=0.017), but other systemic comorbidities did not show a significant association (Supplementary Table S6 with forest plots in Supplementary Figure S14).

### Pneumonia and Acute Clinical Severity

We next analyzed if factors related to pneumonia severity were associated with delirium (Table 1 and Supplementary Figure S15). Delirium was more likely in patients treated with invasive ventilation (OR 3.9 [1.60; 9.33], p=0.005) and less likely in patients treated with non-invasive ventilation (OR 0.4 [0.17; 0.92], p=0.033). The association between delirium and ICU admission did not reach statistical significance (OR 1.7 [0.96; 3.1], p=0.07). Delirium was more likely with renal replacement therapy (OR 4.1 [2.1; 8.2], p=0.012). Pneumonia severity scores (e.g., PSI or CURB-65) were not consistently reported for patients with and without delirium and none of the identified papers split patients by delirium status for septic shock, acute respiratory distress syndrome (ARDS), or pleural effusion. Sensitivity analysis in studies at low risk of bias did not show a significant association with pneumonia severity factors, albeit very few studies reported them, including ICU admission (n=3 studies, OR 2.7 [0.57:12.49], p=0.11) and invasive ventilation (n=1 study, OR 5.3 [0.10; 272.3], p=0.41) (Supplementary Figure S16).

### Clinical Outcomes

We investigated if there was an association between delirium and clinical outcomes (Table 1 and Supplementary Figure S17). Delirium was associated with a significantly increased length of invasive ventilation (n=4 studies, DOM +8.8 days [0.1; 17.5], p=0.047), but not ICU stay (n=2 studies, DOM = +7.6 days [-1.9;17.1], p=0.120) nor length of hospitalization (n=7 studies, DOM +4.4 days, [-0.6; 9.5], p=0.087). Most importantly, delirium was associated with increased mortality (Figure 4A, n=41 studies, OR 4.3 [3.2; 5.7], p<0.001). This association between delirium and mortality remained significant in studies applying multivariable models to control for clinical covariates (n=23 studies, OR 2.9 [2.2; 3.9], p<0.0001, Table 1). Sensitivity analysis showed similar results (Supplementary Table S6). Delirium remained associated with increased length of invasive ventilation, though this was available only in one study (DOM +6 [1.54; 10.46], p= 0.008, Supplementary Figure S18). Importantly, delirium continued to be associated with increased mortality in studies at low risk of bias for both univariate analyses (n=6 studies, OR 3.7 [1.14;12.05], p=0.036, Supplementary Figure S19A) and in studies that controlled for clinical covariates with multivariable analyses (n=5 studies, OR 2.16 [1.2;3.9], p=0.023, Supplementary Figure S19B).

**Figure 4.**
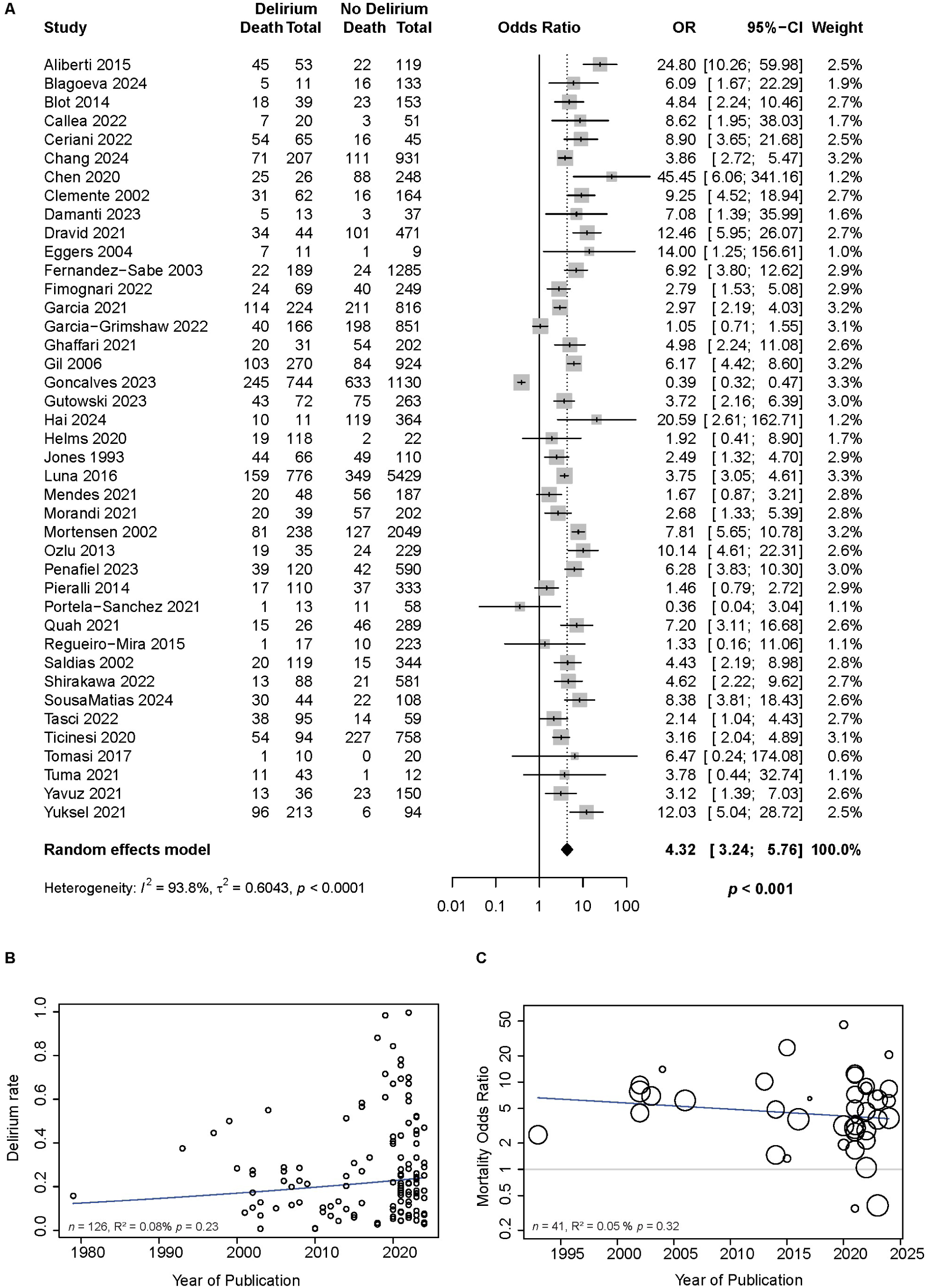
Delirium is associated with significantly increased mortality in patients with pneumonia and has not changed over time. A. Pooled effect of studies demonstrating the association between delirium and mortality using univariate analysis (*n=*41 studies, OR 4.3, 95% CI [3.24; 5.76], p<0.001). This association remained significant in studies applying multivariable models to quantify the relationship of delirium and mortality while controlling for clinical covariates (Table 1). B. Meta-regression analysis measuring the prevalence of delirium in patients hospitalized with pneumonia over time shows steady rates including during the COVID-19 era (R^2^ = 0.08, F_1,124_ = 1.45, p=0.23). C. Notably, the association of delirium and mortality over time remains unchanged (R^2^ = 0.05, F_1,39_ = 1.01, p=0.32). Each circle represents one study, and the size of the circles represents the study weight (studies with a higher precision, i.e., with a smaller standard error, receive a greater weight).

### Delirium prevalence and mortality association over time

Finally, we analyzed if there has been a change in delirium prevalence and its association with mortality over time. Meta-regression analysis of delirium prevalence using year of publication as a continuous variable showed no difference in rates of delirium throughout five decades, including the COVID-19 era (Figure 4B, n=126 studies, R^2^ = 0.08, F_1,124_ = 1.45, p=0.23). Additionally, the association of delirium and mortality over five decades also remained unchanged (Figure 4C, n=126 studies, R^2^ = 0.05, F_1,39_ = 1.01, p=0.32).

## Discussion

To resolve inconsistencies in the occurrence and relationship of delirium with pneumonia, we performed a systematic review and meta-analysis yielding over 100 studies from over five decades of research across multiple continents and languages, diverse hospital settings, and varied pneumonia etiologies, including pre-and post-COVID eras. We provide a robust estimate of delirium, determining that it occurs in approximately one in every five patients with pneumonia (22%). The prior lack of reliable estimates, including uncertainty regarding the roles of patient-specific vs. microbiological risk factors, has hindered delirium management and prognosis. Included studies displayed an expected amount of heterogeneity, however, our adjusted estimate of delirium occurrence –after accounting for small-study effects and between study heterogeneity– yielded similar rates of delirium (19%). Interestingly, our results are in line with systematic reviews on delirium occurrence in hospitalized patients for other common conditions,^33^ e.g., urinary tract infections,^34^ surgical and oncological procedures,^35–37^ and stroke^38^ (17%–25%). This suggests that across diverse conditions, delirium might be more strongly associated with patient-and care-specific factors, rather than unique pathologies or infectious etiologies. The large number of studies in our systemic review allowed us to evaluate these hypotheses directly within patients with pneumonia.

### Delirium assessment methods influence delirium rates

There was substantial variation in how studies defined and assessed delirium, which significantly contributed to delirium rates variability. ICD determination of delirium and less formal and less validated symptom assessments likely underestimate delirium compared to gold standard DSM diagnosis, whereas validated delirium or other mental status assessment scales yield similar, higher estimates. Our sensitivity analysis, which showed nearly double the delirium rate—up to 40%—in studies at low risk of bias, reinforces the latter finding and underscores the need for standardized assessments in clinical care.

Critically, only 30% of the studies used validated delirium methods or standardized assessments of mental status, being more common in studies conducted in the ICU (70%) than hospital wards (30%). Thus, implementation strategies are needed to ensure that standardized assessments are adopted consistently across all care settings to determine the prevalence and impact of delirium reliably.

### Infection-related factors were not associated with delirium rates

We hypothesized that delirium rates might be influenced by pneumonia-specific factors, such as microbiological etiology or infection origin. Our results showed that microbiological etiology, including COVID-19, was not significantly associated with delirium rates. While delirium is a highly recognized neuropsychiatric disorder accompanying COVID-19 infection,^13,14^ we show that in a broader pneumonia context, other groups of etiological agents, such as bacteria (e.g., *Streptococcus pneumoniae* or *Staphylococcus aureus*) or other viruses (e.g., influenza A H1N1), are similarly associated with delirium. Notably, many studies did not document a specific microorganism (*n* = 52), which reflects the difficulty in determining pneumonia etiology using standard culture-based diagnosis. Our findings, however, endorse a mechanism for delirium common to diverse infectious organisms, e.g., an exacerbated inflammatory and immune responses, increased procoagulant states or altered cerebral perfusion, or general hypoxia.^39–42^ Efforts to shed a light into pathophysiological mechanisms related to delirium in severe illness support a key role of inflammation, reflected by increased levels of IL-6, IL-8, IL-10, TNF-α, TNFR1, protein S100β, and pathological pathway convergence.^43–47^ Additionally, infection contracted in healthcare settings compared to community acquired pneumonia had similar rates of delirium, suggesting that the clinical impact and severity of pneumonia may drive delirium more than its cause.

### Care-specific factors were related to delirium rates

Non-microbiologic, case-specific factors significantly influenced delirium rates. Delirium was more common in studies conducted in the ICU setting.^48,49^ And across all studies, care for acute organ failure such as invasive ventilation and dialysis were associated with increased rates of delirium. Unfortunately care information was only provided in a small set of studies at low risk of bias. These data currently cannot determine whether decreased sedation and accelerated weaning from ventilation can decrease delirium,^50–52^ but highlight the importance of studying clinical support for organ failure in high quality studies of delirium and pneumonia, following the criteria identified (e.g. standardized assessment of mental status). Overall, our analysis indicates that delirium in pneumonia is more strongly associated with care-specific factors and illness severity rather than specific infectious etiologies. Our results suggest opportunities for broadly oriented, systems-level delirium prevention and management, which is critical given that up to 40% of delirium could be prevented.^50,53–55^ Such measures are likely to remain important well beyond any particular pandemic (such as COVID-19), given the ongoing evolution of respiratory threats.^56^

### The role of comorbidities in predisposition to delirium

Delirium is thought to have a multifactorial pathophysiology including predisposing and precipitating factors.^57^ In other patient populations, predisposing vulnerabilities include older age, cognitive impairment, critical illness, vision impairment, lower functional status, and dehydration.^58,59^ Within the context of pneumonia, older age, dementia, and stroke continued to be the strongest predisposing factors for delirium, consistent with the frequent superimposition of delirium on severe brain disorders such as dementia and stroke.^38,60,61^ Our results highlight that *pre-established* brain disorders, in which resilience and cognitive reserve are hampered by either neurodegeneration or a localized lesion, increases delirium vulnerability in the context of respiratory infection.^62^

Across all studies, the next strongest predisposing risk factor for delirium, among those with pneumonia, was chronic respiratory illness, including COPD.^63^ A superimposed lung infection in patients with limited lung function may lead more easily to respiratory failure, mechanical ventilation and ICU admission, suggesting decreased resilience across the organ system most affected by pneumonia. Extensive literature suggests a link between smoking and increased risk of delirium in critically ill or post-operative settings, albeit a direct association remains uncertain.^64,65^ Notably, this association seems rather linked to acute smoking cessation and nicotine withdrawal.^64^ Surprisingly, in our study, smoking appeared to be associated with a decreased risk of delirium. Nicotine, as a cholinergic agonist, may have a plausible protective effect given the hypothesis on altered cholinergic activity (deficits) as a mechanism of delirium.^66–68^ However, the effectiveness of nicotine replacement therapy in reducing delirium in critically ill smoking patients is not consistent,^69^ and requires further study.

Other systemic comorbidities were also associated with delirium, including liver disease, kidney disease, and hypertension, remarking the importance of close monitoring of three organ systems that can be directly associated with acute metabolic encephalopathy.^70–72^ However, studies at lower risk of bias did not routinely report rates of systemic predispositions, suggesting the importance of future work to verify which organ system dysfunction are most reliably associated with delirium in pneumonia. Systemic disorders may play an important role in delirium through diverse mechanisms and delirium should be assessed consistently in studies of other organ interventions. For instance, for hepatic encephalopathy, hyperammonemia can induce astrocyte and microglia reactivity, a pro-inflammatory state, altered neurotransmission and oxidative stress, contributing to (acute) impairments in cognitive function.^70^ Similarly, renal failure may exacerbate delirium through an upregulated inflammatory processes that contribute to blood-brain-barrier disruption and release of proinflammatory cytokines.^71^ Blood pressure variability as seen with hypertension in critical illness has also been associated with delirium and linked to microvascular damage and blood-brain-barrier disruption,^73^ which may be a modifiable factor that could be incorporated into delirium-prevention. These predisposing factors should be added or continue to be integrated into predictive models to identify high-risk patients and implement preventive strategies.^74^

### Delirium is associated with worse clinical outcomes in patients with pneumonia

Delirium is consistently associated with worse clinical outcomes including increased mortality across diverse conditions.^60^ Here, delirium was associated with increased length of invasive ventilation, though length of ICU stay and hospitalization were not statistically significantly different. Most importantly, delirium was associated with significantly increased mortality in patients with pneumonia, even in studies controlling for factors that would be independently related to increased death (e.g., age, ventilation status, and comorbidities). We corroborated these findings also in studies at low risk of bias. While delirium may reflect illness severity not fully accounted for by covariates, and its causal relationship with mortality remains undetermined, delirium may contribute to increased mortality in at least some cases through complications such as aspiration, inappropriate antipsychotic use, and falls.^75–77^

### The association between delirium and mortality remains unchanged over the past five decades

Despite increased research into delirium’s pathophysiology and essential guidelines on its prevention and management, ^50–55^ mortality outcomes have seen little improvement. As we discussed, this persistent gap may stem from fundamental issues—such as under-recognition of delirium, misconceptions about its true prevalence and thus, its clinical relevance. Our study addresses this gap by providing a reliable estimate of delirium prevalence in pneumonia and key insights into factors influencing its occurrence and recognition. Ultimately, the entrenched link between delirium and mortality underscores the urgent need for renewed efforts in research, implementation, and clinical practice to change delirium’s prognosis in patients with pneumonia.

### Limitations

General limitations inherent to large systematic reviews were the high heterogeneity across studies, reflecting methodological differences, which we accounted for through pre-planned subgroup analyses. Pharmacologic therapies including sedatives and even antibiotics are important factors associated with delirium, but their relationship was not explored in the studies included, possibly due in part to the difficulty of assessing confounding by indication. We identified an association between delirium and pneumonia severity factors; however, there was surprisingly limited data on the timing of delirium and important complications such as ARDS and lung protective ventilation strategies preventing us from inferring a causal relationship. Further studies are needed to understand the independent relationship between delirium and the risk factors we identified, guided by the above results.

## Conclusions

Delirium is highly prevalent and persistent in pneumonia and identified at higher rates when standardized diagnostic tools are used. Our results emphasize patient and care specific factors, over microbiological causes, including COVID-19. Delirium’s entrenched association with mortality, even controlling for covariates, reinforces the urgent need of improving its prevention and management as a convergent syndrome in pneumonia.

## Data Availability

All data produced in the present study are available upon reasonable request to the authors

## Author contributions

Juarez-Martinez and Kimchi had full access to all the data in the study and take responsibility for the integrity of the data and the accuracy of the data analysis. Concept and design: Kimchi and Juarez-Martinez. Acquisition, analysis, or interpretation of data: All authors. Drafting of the manuscript: Juarez-Martinez and Kimchi. Critical revision of the manuscript for important intellectual content: Kimchi, Juarez-Martinez, Pickens, and Wunderink. Statistical analysis: Juarez-Martinez.

Administrative, technical, or material support: All authors. Supervision: Kimchi

## Data sharing statement

Data available upon request from qualified investigators

## Points for clinical practice

1. Delirium is common in patients with pneumonia, affecting approximately one in five patients.
2. Standardized and validated methods to assess delirium are essential for reliable diagnosis, with consistent use necessary across all care settings.
3. Delirium in pneumonia is primarily associated with care-specific and illness severity factors rather than infectious etiologies, endorsing shared mechanisms across multiple causes of pneumonia.
4. Our results suggest the importance of broadly oriented, systems-level delirium prevention and management beyond the context of any specific respiratory pandemic, such as COVID-19.
5. Delirium’s persistent association with mortality calls for renewed efforts in research, implementation, and clinical efforts to improve delirium prognosis in patients with pneumonia.

