## Supplement for "Five-Decade Prevalence of Delirium in Pneumonia, Risk Factors, and Associated Mortality: A Systematic Review and Meta-Analysis"

### Supplementary Online Content

Erika L. Juarez-Martinez[1], Aida Araia[1], Dillan Prasad[1], Shreya Dhar[1], Khizar Nandoliya[1], Ian G. Sherrington[1], Catherine Zhao[1], Annie Wescott[2], Chiagozie I. Pickens[3], Richard G. Wunderink[3], Eyal Y. Kimchi[1]

[1] Ken & Ruth Davee Department of Neurology. Feinberg School of Medicine. Northwestern University. Chicago, Illinois, 60611

[2] Galter Health Sciences Library & Learning Center, Feinberg School of Medicine, Northwestern University, Chicago, IL, 60611

[3] Division of Pulmonary and Critical Care, Department of Medicine. Feinberg School of Medicine. Northwestern University. Chicago, Illinois, 60611

Methods. Data extraction and management

Table S1. Search Strategies across databases
Table S2 Number of papers retrieved from the databases searched
Table S3. Sensitivity Analysis Grading Criteria. JBI Critical Appraisal Checklist for Analytical Cross-Sectional Studies
Table S4. Included studies and their characteristics (*part 1*)
Table S5. Included studies and their characteristics *(part 2)*
Table S6. Relationship of delirium to Baseline characteristics, Comorbidities, measures of Pneumonia Severity and Acute Care, Mortality, and Length of Clinical Care in studies at low risk of bias.

Figure S1. Traffic light plot for risk of bias in individual studies
Figure S2. Funnel plots for all the included studies suggests the presence of publication bias
Figure S3. Forest plot by risk of bias assessment.
Figure S4. Forest plot by Delirium assessment method
Figure S5. Forest plot by Hospital Setting
Figure S6. Forest plot by Pneumonia Infection Origin
Figure S7. Forest plot by Microbiological etiology
Figure S8. Subgroup analysis in studies at low risk of bias
Figure S9. Forest plots per subgroup analysis in studies at low risk of bias
Figure S10. Older age is a predisposing factor for delirium.
Figure S11. Forest plots by demographics as predisposing factors of delirium
Figure S12. Forest plots by comorbidities as predisposing factors of delirium
Figure S13. Forest plots by demographics as predisposing factors of delirium in studies at low risk of bias
Figure S14. Forest plots by comorbidities as predisposing factors of delirium in studies at low risk of bias.

Figure S15. Forest plots by pneumonia-severity factors
Figure S16. Forest plots by pneumonia-severity factors in studies at low risk of bias.
Figure S17. Forest plots of the associations between delirium and clinical course
Figure S18. Forest plots of the associations between delirium and clinical course in studies at low risk of bias
Figure S19. Delirium is associated with significantly increased mortality in patients with pneumonia in studies at low risk of bias.

#### Supplementary Methods

**Data extraction and management**

Two out of three authors independently extracted data from studies that fulfilled inclusion criteria (E.K., E.J-M., D.P.). Data was extracted and organized by delirium/encephalopathy status. Any discrepancies were resolved through consensus. The following data were extracted from each study when available:

*General study information.* Language, study location, start and end date, type of study, and hospital care setting.

*Study population*. Age, sex, race and ethnicity, comorbidities, Charlson Comorbidity Index,[1] and admission diagnosis.

*Pneumonia diagnosis characteristics.* Pneumonia diagnostic criteria (i.e., respiratory and infectious signs or symptoms, Chest X-ray, CT scan, chart-based, or International Classification of Disease (ICD) codes), pneumonia origin (i.e., community-acquired (CAP), hospital acquired (HAP), healthcare-associated (HCAP), ventilation associated (VAP)). We integrated VAP and HAP to HCAP given the low numbers of studies that reported these origins. Microbiological etiology (i.e., bacterial, COVID-19, other viral or mixed).

*Pneumonia and clinical severity*. CURB-65 scores,[2] Pneumonia severity index (PSI) scores,[3] multilobar pneumonia, pleural effusion, mechanical ventilation, ICU admission, steroid treatment, sepsis, severe sepsis, septic shock, acute respiratory distress syndrome (ARDS), and dialysis.

*Delirium/Encephalopathy assessment methods:* The Diagnostic and Statistics Manual-based assessment (DSM-IV or DSM-5).[4] Validated delirium scales included the confusion assessment method framework (CAM, CAM-ICU or other CAM-based method),[5] the Intensive Care Delirium Screening Checklist (ICDSC),[6] the Delirium Rating Scale-revised (DRS-R-98),[7] the 4 A's delirium assessment tool (4AT), the Nursing Delirium Screening Scale (NuDESC),[8] the Delirium Observation Scale (DOS),[9] and CHART-DEL[10]. Other non-delirium standardized assessments of mental status (AMS) included: the Richmond Agitation-Sedation Scale (RASS),[11] the Glasgow Coma Scale,[12] the Abbreviated Mental Test,[13] and the West-Haven criteria for hepatic encephalopathy.[14] We separately identified mental status ascertainment based on International Classification of Diseases (ICD) codes or if delirium was identified through non-structured, symptom-based reports (e.g. symptoms collection such as confusion, altered mental status, or altered consciousness).

*Delirium characteristics:* periods of delirium assessment, delirium subtypes (hyperactive, hypoactive, or mixed), delirium onset, or other factors associated with delirium.

*Clinical outcomes:* Length of hospitalization and stay in the ICU, length of ventilation, and mortality. For mortality we recorded the odds ratio results of any univariate or multivariable analyses.

#### Supplementary Tables

#### Table S1. Search Strategies across databases.

| | | |
|---|---|---|
| | | **Ovid Medline** |
| **#** | **Search** | **Result** |
| 1 | exp Pneumonia/ | 374039 |
| 2 | (Pneumonia* or Lung-inflammation or Inflammatory-lung-disease or pneumoniae* or Pneumonitis or Pneumonic or Respiratory-tract-infection* or Respiratory-infection* or Bronchopneumonia* or Bronchopneumoniae*).ti,ab. | 289087 |
| 3 | 1 or 2 | 577636 |
| 4 | exp Delirium/ | 13577 |
| 5 | (delirium* or deliria or delirious-state* or clouding-of-consciousness or Encephalopathy).ti,ab. | 78854 |
| 6 | ((acute or organic) adj2 (confusion or brain-syndrome or brain-dysfunction or brain-failure)).ti,ab. | 1473 |
| 7 | ((altered) adj2 (mental-state or mental-status)).ti,ab. | 5108 |
| 8 | 4 or 5 or 6 or 7 | 86201 |
| 9 | exp Hospitalization/ | 304394 |
| 10 | (inpatient* or in-patient* or hospitalise* or hospitalisation* or hospitalize* or hospitalization*).ti,ab. | 2603655 |
| 11 | 9 or 10 | 2754518 |
| 12 | 3 and 8 and 11 | 1349 |
| | | **Cochrane CENTRAL** |
| #1 | MeSH descriptor: [Pneumonia] explode all trees | 12925 |
| #2 | (Pneumonia* or Lung-inflammation or Inflammatory-lung-disease or pneumoniae* or Pneumonitis or Pneumonic or Respiratory-tract-infection* or Respiratory-infection* or Bronchopneumonia* or Bronchopneumoniae*):ti,ab,kw | 36311 |
| #3 | #1 OR #2 | 43138 |
| #4 | MeSH descriptor: [Delirium] explode all trees | 1613 |
| #5 | (delirium* or deliria or delirious-state* or clouding-of-consciousness or Encephalopathy):ti,ab,kw | 10501 |
| #6 | ((acute or organic) NEAR/2 (confusion or brain-syndrome or brain-dysfunction or brain-failure)):ti,ab,kw | 207 |
| #7 | ((altered) NEAR/2 (mental-state or mental-status)):ti,ab,kw | 238 |
| #8 | #4 OR #5 OR #6 OR #7 | 10860 |
| #9 | MeSH descriptor: [Hospitalization] explode all trees | 20543 |
| #10 | (inpatient* or in-patient* or hospitalise* or hospitalisation* or hospitalize* or hospitalization*):ti,ab,kw | 514079 |
| #11 | #9 OR #10 | 520134 |
| #12 | #3 AND #8 AND #11 | 400 |
| | | **Embase (Elsevier)** |
| 1 | 'pneumonia'/exp | |
| 2 | (Pneumonia* or Lung-inflammation or Inflammatory-lung-disease or pneumoniae* or Pneumonitis or Pneumonic or Respiratory-tract-infection* or Respiratory-infection* or Bronchopneumonia* or Bronchopneumoniae*):ti,ab | 440183 |
| 3 | #1 OR #2 | 641745 |
| 4 | 'delirium'/exp | 47236 |
| 5 | (delirium* or deliria or delirious-state* or clouding-of-consciousness or Encephalopathy):ti,ab | 125904 |
| 6 | ((acute or organic) NEAR/2 (confusion or brain-syndrome or brain-dysfunction or brain-failure)):ti,ab | 2359 |
| 7 | ((altered) NEAR/2 (mental-state or mental-status)):ti,ab | 11002 |

| | | |
|---|---|---|
| 8 | #4 OR #5 OR #6 OR #7 | 152921 |
| 9 | 'hospital patient'/exp | 254138 |
| 10 | (inpatient* or in-patient* or hospitalise* or hospitalisation* or hospitalize* or hospitalization):ti,ab | 4132780 |
| 11 | #9 OR #10 | 4159905 |
| 12 | 3 and 8 and 11 | 3136 |
| | | **PsycINFO (EBSCO)** |
| S1 | DE "Pneumonia" | 1015 |
| S2 | TI ( Pneumonia* or Lung-inflammation or Inflammatory-lung-disease or pneumoniae* or Pneumonitis or Pneumonic or Respiratory-tract-infection* or Respiratory-infection* or Bronchopneumonia* or Bronchopneumonia* ) OR AB ( Pneumonia* or Lung-inflammation or Inflammatory-lung-disease or pneumoniae* or Pneumonitis or Pneumonic or Respiratory-tract-infection* or Respiratory-infection* or Bronchopneumonia* or Bronchopneumoniae* ) | 4153 |
| S3 | S1 OR S2 | 4921 |
| S4 | DE "Delirium" | 14199 |
| S5 | TI ( delirium* or deliria or delirious-state* or clouding-of-consciousness or Encephalopathy ) OR AB ( delirium* or deliria or delirious-state* or clouding-of-consciousness or Encephalopathy ) | 1160 |
| S6 | TI ( (acute or organic) N2 (confusion or brain-syndrome or brain-dysfunction or brain-failure) ) OR AB ( (acute or organic) N2 (confusion or brain-syndrome or brain-dysfunction or brain-failure) ) | 474 |
| S7 | TI ( (altered) N2 (mental-state or mental-status) ) OR AB ( (altered) N2 (mental-state or mental-status) ) | 15937 |
| S8 | S4 OR S5 OR S6 OR S7 | 15164 |
| S9 | DE "Hospitalized Patients" | 852258 |
| S10 | TI ( inpatient* or in-patient* or hospitalise* or hospitalisation* or hospitalize* or hospitalization ) OR AB ( inpatient* or in-patient* or hospitalise* or hospitalisation* or hospitalize* or hospitalization ) | |
| S11 | S9 OR S10 | 853076 |
| S12 | S3 AND S8 AND S11 | 165 |
| | | **Scopus (Elsevier)** |
| 1 | ( TITLE-ABS ( pneumonia* OR lung-inflammation OR inflammatory-lung-disease OR pneumoniae* OR pneumonitis OR pneumonic OR respiratory-tract-infection* OR respiratory-infection* OR bronchopneumonia* OR bronchopneumoniae* ) ) AND ( ( TITLE-ABS ( delirium* OR deliria OR delirious-state* OR clouding-of-consciousness OR encephalopathy ) ) OR ( TITLE-ABS ( ( altered ) W/2 ( mental-state OR mental-status ) ) ) OR ( TITLE-ABS ( ( acute OR organic ) W/2 ( confusion OR brain-syndrome OR brain-dysfunction OR brain-failure ) ) ) ) AND ( TITLE-ABS ( inpatient* OR in-patient* OR hospitalise* OR hospitalisation* OR hospitalize* OR hospitalization* ) ) | 868 |

**Table S2. Number of papers retrieved from the databases searched.**

| Databases searched | Date searched | Results |
|---|---|---|
| Ovid MEDLINE | Aug/06/2024 | 1349 |
| Cochrane Central Register of Controlled Trials | Aug06/2024 | 400 |
| Embase | Aug/06/2024 | 3136 |
| PsycINFO | Aug/06/2024 | 165 |
| Scopus | Aug/06/2024 | 868 |
| Total | | 5918 |
| After de-duplication | | 4189 |

**Table S3. Sensitivity Analysis Grading Criteria. JBI Critical Appraisal Checklist for Analytical Cross-Sectional Studies.**

| | |
|---|---|
| **1.** | **Were the criteria for inclusion in the sample clearly defined?**<br>*Description: The authors should provide clear inclusion and exclusion criteria that they developed prior to recruitment of the study participants. The inclusion/exclusion criteria should be specified (e.g., risk, stage of disease progression) with sufficient detail and all the necessary information critical to the study.* |
| **Yes**: | As close to all pneumonia patients as possible. OK if limited to certain patients such as specific pneumonia subtype (COVID-19, influenza, etc.), OK if limited to specific setting (ICU, Respiratory ward, postoperative), and OK if limited to certain age, but if there were too many other exclusions downgrade to “No” |
| **2.** | **Were the study subjects and the setting described in detail?**<br>*Description: The study sample should be described in sufficient detail so that other researchers can determine if it is comparable to the population of interest to them. The authors should provide a clear description of the population from which the study participants were selected or recruited, including demographics, location, and time period.* |
| **Yes**: | Clearly indicated demographics for study cohort, as well as where and when study was conducted. |
| **3.** | **Was the exposure (Pneumonia) measured in a valid and reliable way?**<br>*Description: The study should clearly describe the method of measurement of exposure. Assessing validity requires that a ‘gold standard’ is available to which the measure can be compared. The validity of exposure measurement usually relates to whether a current measure is appropriate or whether a measure of past exposure is needed. Reliability refers to the processes included in an epidemiological study to check repeatability of measurements of the exposures. These usually include intra-observer reliability and inter-observer reliability.* |
| **Yes:** | Clear process for diagnosing pneumonia, combination of respiratory signs/symptoms, signs/symptoms of infection, and corroboration by imaging (CXR or CT). |
| **4.** | **Were objective, standard criteria used for measurement of the condition (pneumonia)?**<br>*Description: It is useful to determine if patients were included in the study based on either a specified diagnosis or definition. This is more likely to decrease the risk of bias. Characteristics are another useful approach to matching groups, and studies that did not use specified diagnostic methods or definitions should provide evidence on matching by key characteristics.* |
| **Yes:** | Clear description of chest imaging (CXR or CT). |
| **5.** | **Were confounding factors identified?**<br>*Description: Confounding has occurred where the estimated intervention exposure effect is biased by the presence of some difference between the comparison groups (apart from the exposure investigated/of interest). Typical confounders include baseline characteristics, prognostic factors, or concomitant exposures (e.g., smoking). A confounder is a difference between the comparison groups, and it influences the direction of the study results. A high-quality study at the level of cohort design will identify the potential confounders and measure them (where possible). This is difficult for studies where behavioral, attitudinal or lifestyle factors may impact on the results.* |
| **Yes**: | Report of at least two of the following: Age, Dementia, Intubation/Mechanical Ventilation, ICU. |
| **6.** | **Were strategies to deal with confounding factors stated?**<br>*Description: Strategies to deal with effects of confounding factors may be dealt within the study design or in data analysis. By matching or stratifying sampling of participants, effects of confounding factors can be adjusted for. When dealing with adjustment in data analysis, assess the statistics used in the study. Most will be some form of multivariate regression analysis to account for the confounding factors measured.* |
| **Yes**: | Mulitvariable model or stratified approach with confounders included as independent variables and delirium as the dependent variable. |
| **7.** | **Were the outcomes (Delirium) measured in a valid and reliable way?**<br>*Description: If for e.g., lung cancer is assessed based on existing definitions or diagnostic criteria, then the answer to this question is likely to be yes. If lung cancer is assessed using observer reported, or self-reported scales, the risk of over- or under-reporting is increased, and objectivity is compromised. Importantly, determine if the measurement tools used were validated instruments as this has a significant impact on outcome assessment validity. Having established the objectivity of the outcome measurement (e.g., lung cancer) instrument, it’s* |

| | |
|---|---|
| | *important to establish how the measurement was conducted. Were those involved in collecting data trained or educated in the use of the instrument/s? (e.g., radiologists). If there was more than one data collector, were they similar in terms of level of education, clinical or research experience, or level of responsibility in the piece of research being appraised?* |
| **Yes:** | Either prospective DSM based diagnosis by expert or validated clinical scale (CAM, CAM-ICU, ICSDC, 4AT, DRS, DOS, West Haven Criteria, CHART-DEL). |
| **8.** | **Was appropriate statistical analysis used?**<br>*Description: As with any consideration of statistical analysis, consideration should be given to whether there was a more appropriate alternate statistical method that could have been used. The methods section should be detailed enough for reviewers to identify which analytical techniques were used (in particular, regression or stratification) and how specific confounders were measured. For studies utilizing regression analysis, it is useful to identify if the study identified which variables were included and how they related to the outcome. If stratification was the analytical approach used, were the strata of analysis defined by the specified variables? Additionally, it is also important to assess the appropriateness of the analytical strategy in terms of the assumptions associated with the approach as differing methods of analysis are based on differing assumptions about the data and how it will respond.* |
| **Yes**: | If included report of how statistical analysis was used, given "Yes" if good description with multivariable regression modeling or at least basic counts. |

*Adapted from*: Moola S, Munn Z, Tufanaru C, Aromataris E, Sears K, Sfetcu R, Currie M, Qureshi R, Mattis P, Lisy K, Mu P-F. Chapter 7: Systematic reviews of etiology and risk.[15]

**Table S4. Included studies and their characteristics** *(part 1)*

| Study ID | Location | Sample size (n) | Hospital Setting | PNA Origin | Microbiology | Delirium Assessment | Age Mean ± SD or Median (IQR 25-75) | Delirium cases |
|---|---|---|---|---|---|---|---|---|
| Aliberti 2015[16] | Europe | 172 | Hospital wards | NS | Bacterial & Viral non-COVID | Delirium Scale | 75 (66– 81) | 53 |
| Aliyu 2003[17] | North America | 67 | ED | CAP | NS | Symp. Collection | 37 ± 8.76 Min, Max (18, 49) | 0 |
| Andrea 2024[18] | South America | 191 | mixed (ICU/non-ICU) | NS | Viral COVID | Symp. Collection | 70.42 ± 7.7 | 48 |
| Aziz 2018[19] | North America | 18036 | Hospital wards | NS | NS | ICD | | 488 |
| Beretta 2023[20] | Europe | 1357 | Hospital wards | NS | Viral COVID | Symp. Collection | | 354 |
| Bhansali 2021[21] | Asia | 43 | Hospital wards | NS | Viral COVID | Symp. Collection | Min 60 | 12 |
| Bianchetti 2020[22] | Europe | 82 | Hospital wards | CAP | Viral COVID | Symp. Collection | 82.6 ± 5.3 | 55 |
| Blagoeva 2024[23] | Europe | 144 | Hospital wards | NS | Viral COVID | Symp. Collection | 67 ± 14.7 Min 25, Max 92 | 11 |
| Blot 2014[24] | Europe | 192 | Hospital wards | Mixed | Bacterial & Viral non-COVID | Symp. Collection | 68.9 ± 18.8 | 39 |
| Callan 2024[25] | North America | 183 | Hospital wards | HCAP | NS | Symp. Collection | | 86 |
| Callea 2022[26] | Europe | 71 | Hospital wards | NS | Viral COVID | DSM | 77 (68–82) | 20 |
| Capuzzi 2023[27] | Europe | 221 | Hospital wards | NS | Viral COVID | Symp. Collection | 67.4 ± 14.5 | 54 |
| Carr 2019[28] | North America | 102 | ICU | HCAP | NS | Symp. Collection | 48 ± 17 | 73 |
| Cataneo-Pina 2023[29] | North America | 48 | Hospital wards | CAP | Viral COVID; NA (CAP not specified) | Delirium Scale | | 25 |
| Ceriani 2022[30] | Europe | 110 | Hospital wards | NS | Viral COVID | Delirium Scale | 81 (79–84) | 65 |
| Chang 2024[31] | North America | 1138 | Hospital wards | NS | Viral COVID | Symp. Collection | 65.4 ± 16.5 | 207 |
| Chen 2020[32] | Asia | 274 | Hospital wards | NS | Viral COVID | Symp. Collection | 62 (44–70) | 26 |
| Clemente 2002[33] | Europe | 226 | ED | CAP | NS | Symp. Collection | 78.71 Min, Max (65, 96) | 62 |
| Cooper 2020[34] | North America | 27 | ICU | NS | Viral COVID | Delirium Scale | 70 (54–76) | 11 |
| D'Ardes 2021[35] | Europe | 56 | Hospital wards | CAP | Viral COVID | Delirium Scale | | 14 |
| Damanti 2023[36] | Europe | 50 | Hospital wards | NS | Viral COVID | Delirium Scale | 79 (73-85) | 13 |

| Study ID | Location | Sample size (n) | Hospital Setting | PNA Origin | Microbiology | Delirium Assessment | Age Mean ± SD or Median (IQR 25-75) | Delirium cases |
|---|---|---|---|---|---|---|---|---|
| deHaan 2023[37] | Europe | 175 | Hospital wards | NS | NS | DSM | | 80 |
| Denke 2018[38] | Europe | 42 | ICU | CAP | Bacterial & Viral non-COVID | Delirium Scale | Md 42 Min, Max (18, 65) | 37 |
| Diaz Fuenzalida 1999[39] | South America | 96 | Hospital wards | CAP | Bacterial | Symp. Collection | 82.3 ± 8.3 | 48 |
| Dravid 2021[40] | Asia | 515 | mixed (ICU/non-ICU) | CAP | Viral COVID | Symp. Collection | 57 (46.5–66) | 44 |
| Eggers 2004[41] | Europe | 20 | ICU | HCAP | NS | DSM | | 11 |
| Ekmekyapar 2022[42] | Asia | 550 | ICU | NS | Viral COVID | Delirium Scale | | 64 |
| Fernandez-Sabe 2003[43] | Europe | 1474 | Hospital wards | CAP | Bacterial & Viral non-COVID | Symp. Collection | 65.17 Min, max (16, 97) | 189 |
| Fimognari 2022[44] | Europe | 318 | Hospital wards | Mixed | NS | Symp. Collection | | 69 |
| Garcia 2021[45] | North America | 1040 | Hospital wards | NS | Viral COVID | Symp. Collection | 55.48 ± 14.47 | 224 |
| Garcia-Grimshaw 2022[46] | North America | 1017 | mixed (ICU/non-ICU) | NS | Viral COVID | Delirium Scale | | 166 |
| Garcia-Vidal 2008[47] | Europe | 2457 | Hospital wards | CAP | Bacterial | Symp. Collection | 65.38 ± 16.85 | 314 |
| Ghaffari 2021[48] | Asia | 233 | Hospital wards | NS | Viral COVID | Symp. Collection | 65.21 ± 16.51 | 31 |
| Gholi 2022[49] | Asia | 310 | ICU | NS | Viral COVID | Delirium Scale | 73 ± 7 | 217 |
| Gil 2006[50] | South America | 1194 | Hospital wards | CAP | NS | Symp. Collection | 68 ± 17 | 270 |
| Gogol 2011[51] | Europe | 81937 | Hospital wards | CAP | NS | Symp. Collection | | 6906 |
| GomezDuque 2023[52] | South America | 283 | ICU | NS | Viral COVID | Delirium Scale | 61.31 ± 13.8 | 148 |
| Goncalves 2023[53] | South America | 1874 | mixed (ICU/non-ICU) | NS | Viral COVID | Symp. Collection | 66.7 ± 10.4 | 744 |
| Goss 2003[54] | North America | 425 | Hospital wards | CAP | NS | Symp. Collection | 46 (18–100) | 29 |
| Guimaraes 2023[55] | South America | 27 | Hospital wards | Mixed | NS | AMS | | 14 |
| Gupta 2024[56] | North America | 15211 | Hospital wards | CAP | NS | Symp. Collection | 69.5 (58-81) | 1080 |
| Gutowski 2023[57] | Europe | 335 | mixed (ICU/non-ICU) | NS | Viral COVID | Symp. Collection | 65.9 ± 15.2 | 72 |

| Study ID | Location | Sample size (n) | Hospital Setting | PNA Origin | Microbiology | Delirium Assessment | Age Mean ± SD or Median (IQR 25-75) | Delirium cases |
|---|---|---|---|---|---|---|---|---|
| Hai 2024[58] | Asia | 375 | Hospital wards | NS | Viral COVID | Symp. Collection | 59.4 ± 16.3 | 11 |
| Helms 1979[59] | North America | 57 | Hospital wards | CAP | Bacterial | Symp. Collection | 43.05 ± 3.58 Min, Max (19, 76) | 9 |
| Helms 2020[60] | Europe | 140 | ICU | NS | Viral COVID | Delirium Scale | 62 (52–70) | 118 |
| Hoogewerf 2006[61] | Europe | 260 | Hospital wards | CAP | Bacterial | Symp. Collection | | 75 |
| Hwang 2020[62] | Asia | 91 | Hospital wards | Mixed | Bacterial | Symp. Collection | 67.42 ± 14.58 | 39 |
| Iribarren-Diarasarri 2023[63] | Europe | 143 | ICU | NS | Viral COVID | Delirium Scale | 61.1 ± 16.2 | 63 |
| Johnson 2000[64] | North America | 148 | Hospital wards | NS | Bacterial | Symp. Collection | M 72.9 | 42 |
| Jolley 2023[65] | North America | 5979 | mixed (ICU/ non-ICU) | NS | Viral COVID | ICD | 61.1 ± 17.02 | 273 |
| Jones 1993[66] | North America | 176 | Hospital wards | NS | Bacterial | Symp. Collection | 58 ± 17.34 | 66 |
| Kaneko 2022[67] | Asia | 112 | ICU | NS | Viral COVID | Symp. Collection | 59.3 ± 13.4 | 24 |
| Kelly 2009[68] | Europe | 80 | ED | CAP | NS | Symp. Collection | 74 (18–95) | 17 |
| Kolditz 2015[69] | Europe | 3427 | Hospital wards | CAP | NS | Symp. Collection | 67.08 Min, Max (18, 102) | 204 |
| Laurichesse 2001[70] | Europe | 37 | Hospital wards | CAP | NS | Symp. Collection | Min, Max (3, 93) | 3 |
| Lee 2023[71] | Asia | 111 | ICU | NS | Viral COVID | DSM | 64.1 ± 13.1 | 26 |
| Leijte 2020[72] | Europe | 769 | Hospital wards | NS | Viral COVID | Symp. Collection | 69.8 ± 13.8 | 41 |
| Lima 2021[73] | South America | 60 | ED | NS | NS | Symp. Collection | | 26 |
| Limpawattana 2016[74] | Asia | 36 | ICU | NS | NS | Delirium Scale | | 21 |
| Lin 2008[75] | Asia | 49 | ICU | NS | NS | Delirium Scale | | 14 |
| Lin 2010[76] | North America | 7554302 | Hospital wards | NS | NS | ICD | | 70372 |
| Lin 2010[77] | North America | 564821 | Hospital wards | NS | NS | ICD | | 4511 |
| Liu 2007[78] | Asia | 302 | Hospital wards | CAP | Bacterial | Symp. Collection | 67.74 ± 10.6 | 35 |
| Liu 2021[79] | Asia | 238 | Hospital wards | NS | Viral COVID | Symp. Collection | 72 (67–80) | 61 |
| Liu 2024[80] | North America | 4483 | Hospital wards | NS | NS | ICD | | 161 |

| Study ID | Location | Sample size (n) | Hospital Setting | PNA Origin | Microbiology | Delirium Assessment | Age Mean ± SD or Median (IQR 25-75) | Delirium cases |
|---|---|---|---|---|---|---|---|---|
| Loponen 2008[81] | Europe | 22 | Hospital wards | NS | NS | Symp. Collection | | 5 |
| Luna 2016[82] | Multilocation | 6205 | Hospital wards | CAP | Bacterial & Viral non-COVID | Symp. Collection | 66.5 ± 17.9 | 776 |
| Manali 2011[83] | Europe | 67 | Hospital wards | CAP | NS | Symp. Collection | 58.8 ± 18.1 | 7 |
| Marrie 2002[84] | North America | 1339 | Hospital wards | CAP | NS | Symp. Collection | 64.28 ± 17.33 | 231 |
| Marrie 2005[85] | North America | 2464 | Hospital wards | CAP | Bacterial | Symp. Collection | 67.69 ± 16.75 | 249 |
| Marrie 2007[86] | North America | 1935 | Hospital wards | CAP | Bacterial | Symp. Collection | 78 ± 11.8 | 385 |
| Martinez 2021[87] | Multilocation | 159 | Hospital wards | HCAP | Bacterial | AMS | 58.9 ± 12.2 | 69 |
| Martins 2022[88] | South America | 59 | ICU | NS | Viral COVID | Symp. Collection | Md 65<br>Min, Max (24, 81) | 29 |
| Matkovska 2019[89] | Europe | 30 | Hospital wards | NS | NS | NS | | 30 |
| Melchio 2021[90] | Europe | 591 | Hospital wards | CAP | NS | AMS | | 102 |
| Mendes 2021[91] | Europe | 235 | Hospital wards | NS | Viral COVID | DSM | 86.3 ± 6.5 | 48 |
| Mendez 2021[92] | Europe | 179 | mixed (ICU/non-ICU) | NS | Viral COVID | Symp. Collection | 57 (49–67) | 8 |
| Morandi 2021[93] | Europe | 241 | Hospital wards | NS | Viral COVID | Delirium Scale | 77.5 (65.6–85) | 39 |
| Mortensen 2002[94] | North America | 2287 | Hospital wards | CAP | NS | Symp. Collection | | 238 |
| Otani 2022[95] | Asia | 149 | ED | NS | Viral COVID | Symp. Collection | | 100 |
| Ozlu 2013[96] | Asia | 264 | mixed (ICU/non-ICU) | NS | Bacterial & Viral non-COVID | Symp. Collection | 47.74 ± 18.67 | 35 |
| Penafiel 2023[97] | South America | 710 | mixed (ICU/non-ICU) | CAP | Viral COVID | Symp. Collection | 59.5 (48-70)<br>Min 18, Max 100 | 120 |
| Pieralli 2014[98] | Europe | 443 | Hospital wards | CAP | NS | Delirium Scale | 81.8 ± 7.5 | 110 |
| Portela-Sanchez 2021[99] | Europe | 71 | mixed (ICU/non-ICU) | NS | Viral COVID | Symp. Collection | Md 69<br>Min, Max (23, 91) | 13 |
| Prabhahar 2024[100] | Asia | 68 | ED | NS | NS | Symp. Collection | | 21 |
| Premkumar 2019[101] | Asia | 110 | ICU | NS | Bacterial & Viral non-COVID | AMS | 47.7 ± 13.31 | 67 |
| Quah 2021[102] | Asia | 315 | Hospital wards | CAP | Bacterial & Viral COVID | Symp. Collection | 71.4 ± 18 | 26 |
| Regueiro-Mira 2015[103] | Europe | 240 | Hospital wards | Mixed | Bacterial | Symp. Collection | 57.2 ± 15.4<br>54.5 (55–59) | 17 |

| Study ID | Location | Sample size (n) | Hospital Setting | PNA Origin | Microbiology | Delirium Assessment | Age Mean ± SD or Median (IQR 25-75) | Delirium cases |
|---|---|---|---|---|---|---|---|---|
| Riquelme 1997[104] | South America | 101 | Hospital wards | CAP | Bacterial | DSM | 78 ± 8 | 45 |
| Riquelme 2006[105] | South America | 200 | Hospital wards | CAP | Bacterial | Symp. Collection | 63 ± 19 | 54 |
| Rothberg 2013[106] | North America | 69344 | Hospital wards | CAP | NS | Symp. Collection | | 2975 |
| Ruiz 2014[107] | Europe | 399 | Hospital wards | NS | Bacterial | Symp. Collection | | 39 |
| Sabzwari 2014[108] | Asia | 39 | Hospital wards | NS | NS | Symp. Collection | | 20 |
| Sakakibara 2022[109] | Asia | 136 | Hospital wards | NS | NS | Symp. Collection | M 75<br>Min, max (3, 98) | 136 |
| Saldias 2002[110] | South America | 463 | Hospital wards | CAP | Bacterial & Viral non-COVID | Symp. Collection | 68.8 ± 18.7<br>Min, max (16, 101) | 119 |
| Serrano 2023[111] | Europe | 1371 | Hospital wards | CAP | Bacterial | Symp. Collection | M 56.7 | 123 |
| Serrano Fernandez 2022[112] | Europe | 2224 | Hospital wards | CAP | Bacterial & Viral COVID | Symp. Collection | | 153 |
| Shirakawa 2022[113] | Asia | 669 | Hospital wards | NS | NS | Symp. Collection | 78 (71–82) | 88 |
| Soares 2022[114] | Europe | 165 | mixed (ICU/non-ICU) | NS | NS | Symp. Collection | | 50 |
| Sorrell 2023[115] | Europe | 264 | Hospital wards | Mixed | Viral COVID | Symp. Collection | | 88 |
| SousaMatias 2024[116] | South America | 152 | Hospital wards | CAP | Bacterial | Symp. Collection | Md 58 | 44 |
| Suwanpasu 2016[117] | Asia | 23 | Hospital wards | CAP | NS | Delirium Scale | M 81.3 | 13 |
| Szylinska 2020[118] | Europe | 217 | Hospital wards | NS | NS | DSM | | 86 |
| Tasci 2022[119] | Asia | 154 | mixed (ICU/non-ICU) | NS | Viral COVID | AMS | 72.3 ± 15 | 95 |
| Thabet 2022[120] | Asia | 1413 | Hospital wards | Mixed | Bacterial | AMS | | 249 |
| Ticinesi 2020[121] | Europe | 852 | Hospital wards | NS | Viral COVID | Delirium Scale | 73 ± 14 | 94 |
| Tomasi 2017[122] | South America | 30 | Hospital wards | CAP | NS | Delirium Scale | | 10 |
| Trevisan 2023[123] | Europe | 1160 | Hospital wards | NS | Viral COVID | DSM | | 197 |
| Tuma 2021[124] | South America | 55 | mixed (ICU/ non-ICU) | NS | Viral COVID | AMS | 59.84 ± 13.8 | 43 |
| Uginet 2021[125] | Europe | 707 | mixed (ICU/non-ICU) | NS | NS | Symp. Collection | | 31 |

| Study ID | Location | Sample size (n) | Hospital Setting | PNA Origin | Microbiology | Delirium Assessment | Age Mean ± SD or Median (IQR 25-75) | Delirium cases |
|---|---|---|---|---|---|---|---|---|
| vanderKnaap 2024[126] | Europe | 324 | ICU | NS | Viral COVID | Symp. Collection | 64 (57-72) | 48 |
| Viasus 2012[127] | Europe | 348 | Hospital wards | NS | Bacterial & Viral non-COVID | Symp. Collection | 44 (33–55) | 23 |
| Vinogradov 2021[128] | Multilocation | 30 | mixed (ICU/non-ICU) | CAP | Viral COVID | DSM | 48.93 ± 8.47 | 10 |
| Viscogliosi 2016[129] | Europe | 159 | Hospital wards | CAP | NS | DSM | 80 ± 9.1 | 43 |
| Watts 2012[130] | North America | 363 | ED | CAP | Bacterial & Viral non-COVID | Symp. Collection | 63.03 ± 17.96 | 40 |
| Wrenn 2023[131] | North America | 300 | ED | CAP | NS | Symp. Collection | 60 (43-72) | 46 |
| Xing 2020[132] | Asia | 100 | Hospital wards | NS | NS | DSM | 66.5 ± 3.93 | 22 |
| Yang 2020[133] | North America | 2031 | Hospital wards | NS | NS | ICD | | 127 |
| Yang 2022[134] | North America | 5761 | Hospital wards | NS | NS | ICD | | 333 |
| Yang 2023[135] | North America | 834 | Hospital wards | NS | NS | ICD | | 58 |
| Yavuz 2021[136] | Asia | 186 | ED | CAP | NS | Symp. Collection | 79.4 ± 8.7<br>Med 79, Range 65-104 | 36 |
| Yenibertiz 2021[137] | Asia | 45 | Hospital wards | NS | NS | DSM | | 34 |
| Yuksel 2021[138] | Asia | 307 | mixed (ICU/non-ICU) | NS | Viral COVID | Symp. Collection | 68.02 ± 15.64 | 213 |
| Zerbit 2022[139] | Europe | 85 | mixed (ICU/non-ICU) | NS | Viral COVID | Symp. Collection | 60 (49-69) | 6 |
| Zhang 2018[140] | Asia | 1902 | ED | CAP | NS | Symp. Collection | 73 (61–82) | 65 |
| Zukowska 2023[141] | Europe | 33 | Hospital wards | HCAP | NS | Symp. Collection | | 6 |

Abbreviations: Intensive Care Unit (ICU), Emergency Department (ED). Community Acquired Pneumonia (CAP), Healthcare Acquired Pneumonia (HCAP), Not Specified (NS). Diagnostic and Statistical Manual (DSM); Delirium Scales: Confusion Assessment Method (CAM, CAM-ICU), Delirium Observation Scale (DOS), 4 A's Delirium Assessment Tool (4AT), Intensive Care Delirium Screening Checklist (ICDSC), and chart-validated scale (CHART-DEL);AMS scales: Richmond Agitation-Sedation Scale (RASS), Glasgow Coma Scale (GCS), Abbreviated Mental Test, and West-Haven Criteria for Hepatic Encephalopathy. International Classification of Diseases (ICD code). Retrospective (Retros.) and Prospective (Pros.) ascertainment of symptoms.

**Table S5. Included studies and their characteristics** *(part 2)*

| Study | Study type | Pneumonia Diagnosis Criteria | Delirium Assessment Method specifications | Delirium onset (*n*=patients) | |
|---|---|---|---|---|---|
| | | | | On admission | During hosp. |
| Aliberti 2015 | Retros. | Respiratory signs/symptoms (clinical); Infectious signs/symptoms (clinical); Chest XRay | DeliriumScale: CHART-DEL | | |
| Aliyu 2003 | Retros. | ICD code only | Symp. Collection | | |
| Andrea 2024 | Retros. | CT scan | Symp. Collection | | |
| Aziz 2018 | Retros. | ICD code only | ICD | | |
| Beretta 2023 | Mixed | Respiratory signs/symptoms (clinical); Infectious signs/symptoms (clinical); Chest XRay | Symp. Collection | | |
| Bhansali 2021 | Retros. | Respiratory signs/symptoms (clinical); Infectious signs/symptoms (clinical); Chest XRay | Symp. Collection | | |
| Bianchetti 2020 | Retros. | Respiratory signs/symptoms (clinical); Infectious signs/symptoms (clinical) | Symp. Collection | 55 | |
| Blagoeva 2024 | Retros. | Respiratory signs/symptoms (clinical); Infectious signs/symptoms (clinical); Chest XRay; CT scan | Symp. Collection | 11 | |
| Blot 2014 | Mixed | Respiratory signs/symptoms (clinical); Infectious signs/symptoms (clinical); Chest XRay | Symp. Collection | | |
| Callan 2024 | Retros. | Not described | Symp. Collection | | 86 |
| Callea 2022 | NA | Respiratory signs/symptoms (clinical); Infectious signs/symptoms (clinical); Chest XRay; CT scan | DSM + CAM-ICU and RASS | 8 | 12 |
| Capuzzi 2023 | Retros. | Not described | Symp. Collection | | |
| Carr 2019 | Retros. | Respiratory signs/symptoms (clinical); Infectious signs/symptoms (clinical); Chest XRay | Symp. Collection | | |
| Cataneo-Pina 2023 | Retros. | Not described | DeliriumScale: 3D-CAM | | |
| Ceriani 2022 | Retros. | Not described | DeliriumScale: CHART-DEL | | |
| Chang 2024 | Retros. | Respiratory signs/symptoms (clinical); Chest XRay | Symp. Collection | | |
| Chen 2020 | Retros. | Not described | Symp. Collection | | |
| Clemente 2002 | Retros. | Respiratory signs/symptoms (clinical); Infectious signs/symptoms (clinical); Chest XRay | Symp. Collection | | |
| Cooper 2020 | Prosp. | Not described | DeliriumScale: ICDSC | | |
| D'Ardes 2021 | Prosp. | Respiratory signs/symptoms (clinical); Infectious signs/symptoms (clinical); CT scan | DeliriumScale: 4AT | | 14 |
| Damanti 2023 | Prosp. | COVID-BioB protocol | DeliriumScale: 4AT | | |
| deHaan 2023 | Mixed | Not described | DSM + DOS | | |

| Study | Study type | Pneumonia Diagnosis Criteria | Delirium Assessment Method specifications | Delirium onset (*n*=patients) | |
|---|---|---|---|---|---|
| | | | | On admission | During hosp. |
| Denke 2018 | Prosp. | Not described | DeliriumScale: CAM-ICU; RASS | | 37 |
| DiazFuenzalida 1999 | Retros. | Respiratory signs/symptoms (clinical); Infectious signs/symptoms (clinical); Chest XRay | Symp. Collection | | |
| Dravid 2021 | Retros. | Respiratory signs/symptoms (clinical); Infectious signs/symptoms (clinical); Chest XRay; CT scan | Symp. Collection | | |
| Eggers 2004 | Mixed | Respiratory signs/symptoms (clinical); Infectious signs/symptoms (clinical); Chest XRay; Other: CDC Definitions for pneumonia | DSM | | |
| Ekmekyapar 2022 | Retros. | Not described | DeliriumScale: CAM-ICU | | |
| Fernandez-Sabe 2003 | Prosp. | Respiratory signs/symptoms (clinical); Infectious signs/symptoms (clinical); Chest XRay | Symp. Collection | | |
| Fimognari 2022 | Prosp. | Respiratory signs/symptoms (clinical); Infectious signs/symptoms (clinical); Chest XRay; CT scan | Symp. Collection | 69 | |
| Garcia 2021 | Retros. | Chart based only | Symp. Collection | | |
| Garcia-Grimshaw 2022 | Retros. | CT scan | DeliriumScale: CAM-ICU | | 166 |
| Garcia-Vidal 2008 | Prosp. | Respiratory signs/symptoms (clinical); Infectious signs/symptoms (clinical); Chest XRay | Symp. Collection | 314 | |
| Ghaffari 2021 | Retros. | Respiratory signs/symptoms (clinical); Infectious signs/symptoms (clinical); CT scan | Symp. Collection | | |
| Gholi 2022 | Prosp. | Respiratory signs/symptoms (clinical); Infectious signs/symptoms (clinical); CT scan | DeliriumScale: CAM-ICU | | |
| Gil 2006 | Prosp. | Respiratory signs/symptoms (clinical); Infectious signs/symptoms (clinical); Chest XRay | Symp. Collection | | |
| Gogol 2011 | Retros. | Not described | Symp. Collection | | |
| GomezDuque 2023 | Retros. | Not described | DeliriumScale: CAM-ICU | | |
| Goncalves 2023 | Mixed | Respiratory signs/symptoms (clinical); Chest XRay; CT scan | Symp. Collection | | |
| Goss 2003 | Prosp. | Respiratory signs/symptoms (clinical); Infectious signs/symptoms (clinical); Chest XRay | Symp. Collection | 29 | |
| GuimarÃ£es 2023 | Prosp. | Respiratory signs/symptoms (clinical); Infectious signs/symptoms (clinical); Chest XRay | AMS scale: West Haven criteria for hepatic encephalopathy | 7 | 7 |
| Gupta 2024 | Mixed | Respiratory signs/symptoms (clinical); Infectious signs/symptoms (clinical); Chest XRay; CT scan | Symp. Collection | | |
| Gutowski 2023 | Retros. | Severe COVID | Symp. Collection | | |

| Study | Study type | Pneumonia Diagnosis Criteria | Delirium Assessment Method specifications | Delirium onset (*n*=patients) | |
|---|---|---|---|---|---|
| | | | | On admission | During hosp. |
| Hai 2024 | NA | Respiratory signs/symptoms (clinical); Infectious signs/symptoms (clinical); Chest XRay | Symp. Collection | | |
| Helms 1979 | Retros. | Respiratory signs/symptoms (clinical); Infectious signs/symptoms (clinical); Chest Xray | Symp. Collection | | |
| Helms 2020 | Prosp. | Respiratory signs/symptoms (clinical); Infectious signs/symptoms (clinical); CT scan | DeliriumScale: CAM-ICU; RASS | 22 | 97 |
| Hoogewerf 2006 | Prosp. | Respiratory signs/symptoms (clinical); Infectious signs/symptoms (clinical); Chest XRay | Symp. Collection | | |
| Hwang 2020 | Retros. | Chart based only | Symp. Collection | | |
| Iribarren-Diarasarri 2023 | Prosp. | Not described | DeliriumScale: CAM-ICU | | |
| Johnson 2000 | Retros. | ICD code only | Symp. Collection | | |
| Jolley 2023 | Retros. | Chart based only | ICD | | |
| Jones 1993 | Retros. | Chest XRay; Chart based only | Symp. Collection | | |
| Kaneko 2022 | Retros. | Not described | Symp. Collection | | |
| Kelly 2009 | Prosp. | Respiratory signs/symptoms (clinical); Infectious signs/symptoms (clinical); Chest XRay | Symp. Collection | | |
| Kolditz 2015 | Prosp. | Respiratory signs/symptoms (clinical); Infectious signs/symptoms (clinical); Chest XRay; ATS/IDSA 2007 minor criteria for severe pneumonia | Symp. Collection | | |
| Laurichesse 2001 | Prosp. | Respiratory signs/symptoms (clinical); Infectious signs/symptoms (clinical); Chest XRay | Symp. Collection | | |
| Lee 2023 | Retros. | Not described | DSM + CAM-ICU and RASS | | |
| Leijte 2020 | Retros. | Respiratory signs/symptoms (clinical); Infectious signs/symptoms (clinical); CT scan | Symp. Collection | 41 | |
| Lima 2021 | Prosp. | Not described | Symp. Collection | | 26 |
| Limpawattana 2016 | Prosp. | Not described | DeliriumScale: CAM-ICU | | |
| Lin 2008 | NA | Not described | DeliriumScale: CAM-ICU; RASS; GCS | | |
| Lin 2010 | Retros. | Diagnosis-related groups [DRGs] categories for pneumonia | ICD | | |
| Lin 2010 | Retros. | Center for Medicare and Medicaid Services DRG (CMS-DRG classifications) categories fro pneumonia | ICD | 3241 | 668 |

| Study | Study type | Pneumonia Diagnosis Criteria | Delirium Assessment Method specifications | Delirium onset (*n*=patients) | |
|---|---|---|---|---|---|
| | | | | **On admission** | **During hosp.** |
| Liu 2007 | Retros. | Respiratory signs/symptoms (clinical); Infectious signs/symptoms (clinical); Chest XRay | Symp. Collection | | |
| Liu 2021 | Retros. | Respiratory signs/symptoms (clinical); Infectious signs/symptoms (clinical) | Symp. Collection | | |
| Liu 2024 | Retros. | ICD code only | ICD | | |
| Loponen 2008 | Mixed | Not described | Symp. Collection | | |
| Luna 2016 | Retros. | Respiratory signs/symptoms (clinical); Infectious signs/symptoms (clinical); Chest Xray | Symp. Collection | | |
| Manali 2011 | Retros. | Respiratory signs/symptoms (clinical); Infectious signs/symptoms (clinical); Chest XRay; CT scan | Symp. Collection | | |
| Marrie 2002 | Prosp. | Respiratory signs/symptoms (clinical); Infectious signs/symptoms (clinical); Chest XRay | Symp. Collection | | |
| Marrie 2005 | Prosp. | Respiratory signs/symptoms (clinical); Infectious signs/symptoms (clinical); Chest XRay | Symp. Collection | | |
| Marrie 2007 | Prosp. | Respiratory signs/symptoms (clinical); Infectious signs/symptoms (clinical); Chest XRay | Symp. Collection | | |
| Martinez 2021 | Mixed | Infectious signs/symptoms (clinical); Chest Xray | AMS scale: hepatic encephalopathy grade III-IV | | |
| Martins 2022 | Retros. | Not described | Symp. Collection | | |
| Matkovska 2019 | NA | Not described | NA | | |
| Melchio 2021 | Mixed | Respiratory signs/symptoms (clinical); Infectious signs/symptoms (clinical); Chest Xray | AMS scale: Abbrev. mental test | 102 | |
| Mendes 2021 | Retros. | Respiratory signs/symptoms (clinical); Infectious signs/symptoms (clinical); Chest XRay; CT scan | DSM + CAM | 48 | |
| Mendez 2021 | Prosp. | Infectious signs/symptoms (clinical); Chest Xray | Symp. Collection | | |
| Morandi 2021 | Retros. | Infectious signs/symptoms (clinical); Chest XRay; CT scan; Recorded from the chart | DeliriumScale: 4AT and CHART-DEL | 39 | |
| Mortensen 2002 | Prosp. | Respiratory signs/symptoms (clinical); Infectious signs/symptoms (clinical); Chest XRay | Symp. Collection | | |
| Otani 2022 | NA | Not described | Symp. Collection | | |
| Ozlu 2013 | Retros. | Respiratory signs/symptoms (clinical); Infectious signs/symptoms (clinical); Chest XRay | Symp. Collection | | |
| Penafiel 2023 | Prosp. | Respiratory signs/symptoms (clinical); Infectious signs/symptoms (clinical); Chest XRay | Symp. Collection | | |
| Pieralli 2014 | Retros. | Respiratory signs/symptoms (clinical); Infectious signs/symptoms (clinical); Chest XRay; CT scan | DeliriumScale: CAM | | 110 |

| Study | Study type | Pneumonia Diagnosis Criteria | Delirium Assessment Method specifications | Delirium onset (*n*=patients) | |
|---|---|---|---|---|---|
| | | | | On admission | During hosp. |
| Portela-Sanchez 2021 | Prosp. | Infectious signs/symptoms (clinical); Chest XRay | Symp. Collection | | |
| Prabhahar 2024 | Retros. | Respiratory signs/symptoms (clinical); Infectious signs/symptoms (clinical); Chest XRay; CT scan | Symp. Collection | | |
| Premkumar 2019 | Prosp. | Respiratory signs/symptoms (clinical); Infectious signs/symptoms (clinical); Chest XRay | AMS | | |
| Quah 2021 | Mixed | Respiratory signs/symptoms (clinical); Infectious signs/symptoms (clinical); Chest XRay | Symp. Collection | | |
| Regueiro-Mira 2015 | Retros. | Respiratory signs/symptoms (clinical); Infectious signs/symptoms (clinical); Chest XRay; Criteria approved by the Spanish Society of Pulmonology and Thoracic Surgery (SEPAR). Confirmation by antigen determination of L. pneumophila in urine | Symp. Collection | | |
| Riquelme 1997 | Prosp. | Respiratory signs/symptoms (clinical); Infectious signs/symptoms (clinical); Chest XRay | DSM | 45 | |
| Riquelme 2006 | Prosp. | Respiratory signs/symptoms (clinical); Infectious signs/symptoms (clinical); Chest XRay | Symp. Collection | | |
| Rothberg 2013 | Retros. | ICD code only | Symp. Collection | | |
| Ruiz 2014 | Prosp. | Respiratory signs/symptoms (clinical); Infectious signs/symptoms (clinical); Chest XRay | Symp. Collection | | |
| Sabzwari 2014 | Retros. | Not described | Symp. Collection | | |
| Sakakibara 2022 | Retros. | Not described | Symp. Collection | | |
| Saldias 2002 | Prosp. | Respiratory signs/symptoms (clinical); Infectious signs/symptoms (clinical); Chest XRay | Symp. Collection | | |
| Serrano 2023 | Prosp. | Respiratory signs/symptoms (clinical); Infectious signs/symptoms (clinical); Chest XRay; Other: urinary antigen tests | Symp. Collection | | |
| SerranoFernandez 2022 | Prosp. | Respiratory signs/symptoms (clinical); Infectious signs/symptoms (clinical); Chest XRay | Symp. Collection | | |
| Shirakawa 2022 | Retros. | ICD code only | Symp. Collection | | |
| Soares 2022 | Retros. | Stroke-associated pneumonia- based on CDC Criteria | Symp. Collection | | |
| Sorrell 2023 | Retros. | Not described | Symp. Collection | | |
| SousaMatias 2024 | Retros. | Respiratory signs/symptoms (clinical); Infectious signs/symptoms (clinical); Chest XRay | Symp. Collection | | |

| Study | Study type | Pneumonia Diagnosis Criteria | Delirium Assessment Method specifications | Delirium onset (*n*=patients) | |
|---|---|---|---|---|---|
| | | | | On admission | During hosp. |
| Suwanpasu 2016 | Prosp. | Not described | DeliriumScale: CAM | | |
| Szylinska 2020 | Retros. | Not described | DSM + CAM-ICU | | |
| Tasci 2022 | Retros. | CT scan | AMS scale: RASS | | |
| Thabet 2022 | Prosp. | Respiratory signs/symptoms (clinical); Infectious signs/symptoms (clinical); Chest XRay; CT scan; ATS/IDSA citation | AMS scale: GCS | | |
| Ticinesi 2020 | Retros. | Respiratory signs/symptoms (clinical); Infectious signs/symptoms (clinical); CT scan | DeliriumScale: CAM shortened version | | 94 |
| Tomasi 2017 | Prosp. | Chart based only | DeliriumScale: CAM | | |
| Trevisan 2023 | Retros. | Chest XRay; CT scan | DSM + RASS | | |
| Tuma 2021 | Retros. | CT scan | AMS Scale:RASS | | |
| Uginet 2021 | Retros. | Respiratory signs/symptoms (clinical); Infectious signs/symptoms (clinical) | Symp. Collection | | |
| vanderKnaap 2024 | Prosp. | Respiratory signs/symptoms (clinical); Infectious signs/symptoms (clinical); CT scan | Symp. Collection | | |
| Viasus 2012 | Prosp. | Respiratory signs/symptoms (clinical); Infectious signs/symptoms (clinical); Chest XRay | Symp. Collection | | |
| Vinogradov 2021 | Prosp. | CT scan | DSM | | |
| Viscogliosi 2016 | Prosp. | Not described | DSM +CAM | | |
| Watts 2012 | Retros. | Chart based only | Symp. Collection | | |
| Wrenn 2023 | Retros. | Chart based only; ICD code only | Symp. Collection | | |
| Xing 2020 | NA | Not described | DSM | | |
| Yang 2020 | Retros. | ICD code only | ICD | | |
| Yang 2022 | Retros. | ICD code only | ICD | | |
| Yang 2023 | Retros. | ICD code only | ICD | | |
| Yavuz 2021 | Retros. | Respiratory signs/symptoms (clinical); Infectious signs/symptoms (clinical); Chest XRay | Symp. Collection | | |
| Yenibertiz 2021 | Retros. | Not described | DSM | | |
| Yuksel 2021 | Retros. | Respiratory signs/symptoms (clinical); Infectious signs/symptoms (clinical); Chest XRay; CT scan | Symp. Collection | | |
| Zerbit 2022 | Retros. | Respiratory signs/symptoms (clinical); Chest XRay; CT scan | Symp. Collection | | |
| Zhang 2018 | Retros. | Respiratory signs/symptoms (clinical); Infectious signs/symptoms (clinical); Chest Xray | Symp. Collection | | |

| Study | Study type | Pneumonia Diagnosis Criteria | Delirium Assessment Method specifications | Delirium onset (*n*=patients) | |
|---|---|---|---|---|---|
| | | | | On admission | During hosp. |
| Zukowska 2023 | Retros. | Respiratory signs/symptoms (clinical); Infectious signs/symptoms (clinical); Chest XRay; CT scan; ECDC criteria | Symp. Collection | | |

Abbreviations: Not Specified (NA). Diagnostic and Statistical Manual (DSM); Delirium Scales: Confusion Assessment Method (CAM, CAM-ICU), Delirium Observation Scale (DOS), 4 A's Delirium Assessment Tool (4AT), Intensive Care Delirium Screening Checklist (ICDSC), and chart-validated scale (CHART-DEL);AMS scales: Richmond Agitation-Sedation Scale (RASS), Glasgow Coma Scale (GCS), Abbreviated Mental Test, and West-Haven Criteria for Hepatic Encephalopathy. International Classification of Diseases (ICD code), Symptom (Symp) Collection. Restrospective (Retros.), Prospective (Prosp.).

**Table S6. Relationship of delirium to baseline characteristics, Comorbidities, measures of Pneumonia Severity and Acute Care, Mortality, and Length of Clinical Care in Studies at Low Risk of Bias.**

| **Baseline characteristics** | **OR** | **95% CI** | ***p*-value** |
|---|---|---|---|
| Female (n=5) | 0.77 | [0.47; 1.25] | 0.204 |
| Nursing home (n=1) | 2.33 | [1.15; 4.74] | 0.019* |
| Age | DOM +6.20 | [1.49;10.92] | 0.01* |
| **Comorbidities** | **OR** | **95% CI** | **p-value** |
| Dementia (n=3) | 4.12 | [1.91; 8.89] | 0.015* |
| Stroke (n=4) | 2.21 | [1.31; 3.74] | 0.017* |
| Respiratory disease (n=3) | 1.97 | [0.75; 5.14] | 0.093 |
| Liver Disease (n=1) | 0.97 | [0.04; 20.80] | 0.982 |
| Kidney Disease (n=3) | 1.45 | [0.08; 24.67] | 0.631 |
| Hypertension (n=3) | 1.21 | [0.33; 4.45] | 0.601 |
| Heart Disease (n=4) | 0.78 | [0.18; 3.51] | 0.64 |
| Diabetes (n=4) | 1.11 | [0.72; 1.72] | 0.484 |
| Cancer (n=3) | 1.06 | [0.08; 13.62] | 0.933 |
| **Pneumonia Severity and Acute Care** | | | |
| ICU Admission (n=3) | 2.67 | [0.57; 12.49] | 0.112 |
| Invasive Ventilation (n=1) | 5.27 | [0.10; 272.37] | 0.409 |
| Noninvasive Ventilation (n=2) | 1.58 | [0.00; 55214680.00] | 0.794 |
| Steroids (n=1) | 2.07 | [0.10; 45.04] | 0.643 |
| **Length of Clinical Care** | **DOM** | **95%CI** | ***p*-value** |
| Length of Invasive ventilation (n = 1) | +6.00 | [1.54; 10.46] | 0.008* |
| Length of ICU stay (n = 1) | +4.67 | [-0.65; 9.98] | 0.085 |
| Length of Hospitalization (n = 1) | +0.80 | [-3.01; 1.41] | 0.470 |
| **Mortality** | | | |
| Overall Death (univariate analysis) (n=6) | 3.70 | [1.14; 12.05] | 0.036* |
| Overall Death (multivariate analysis (n = 5) | 2.16 | [1.19; 3.91], | 0.023* |

All comorbidities reflect reports of chronic disease prior to admission. *n* = studies, OR = odds ratios, DOM = difference of means, CI = Confidence interval. Significant p-value <0.05 (*). Forest plots and measures of heterogeneity for each baseline characteristics, comorbidities, pneumonia severity factors and acute care, length of clinical care, and mortality for studies at low risk of bias are shown in Supplementary Figures S13, S14, S16, S18 and S19 respectively).

#### Supplementary Figures

##### Figure S1. Traffic light plot for risk of bias in individual studies

Risk of bias was assessed over eight domains (D1–D8). Overall risk of bias was classified as "low" *(green circles)* if studies met ≥6/8 domains including clear ascertainment of both pneumonia and delirium using standardized methods or validated clinical scores. Risk of bias was classified as "unclear" *(yellow circles)* if studies met 6/8 domains, but pneumonia and/or delirium diagnosis were not clearly ascertained, and as "high" *(red circles)* if they meet <6/8 domains and no clear ascertainment for pneumonia or delirium using standardized methods were used. Plot created using *robvis*, a web app built in R for visualizing risk-of-bias assessments.[142]

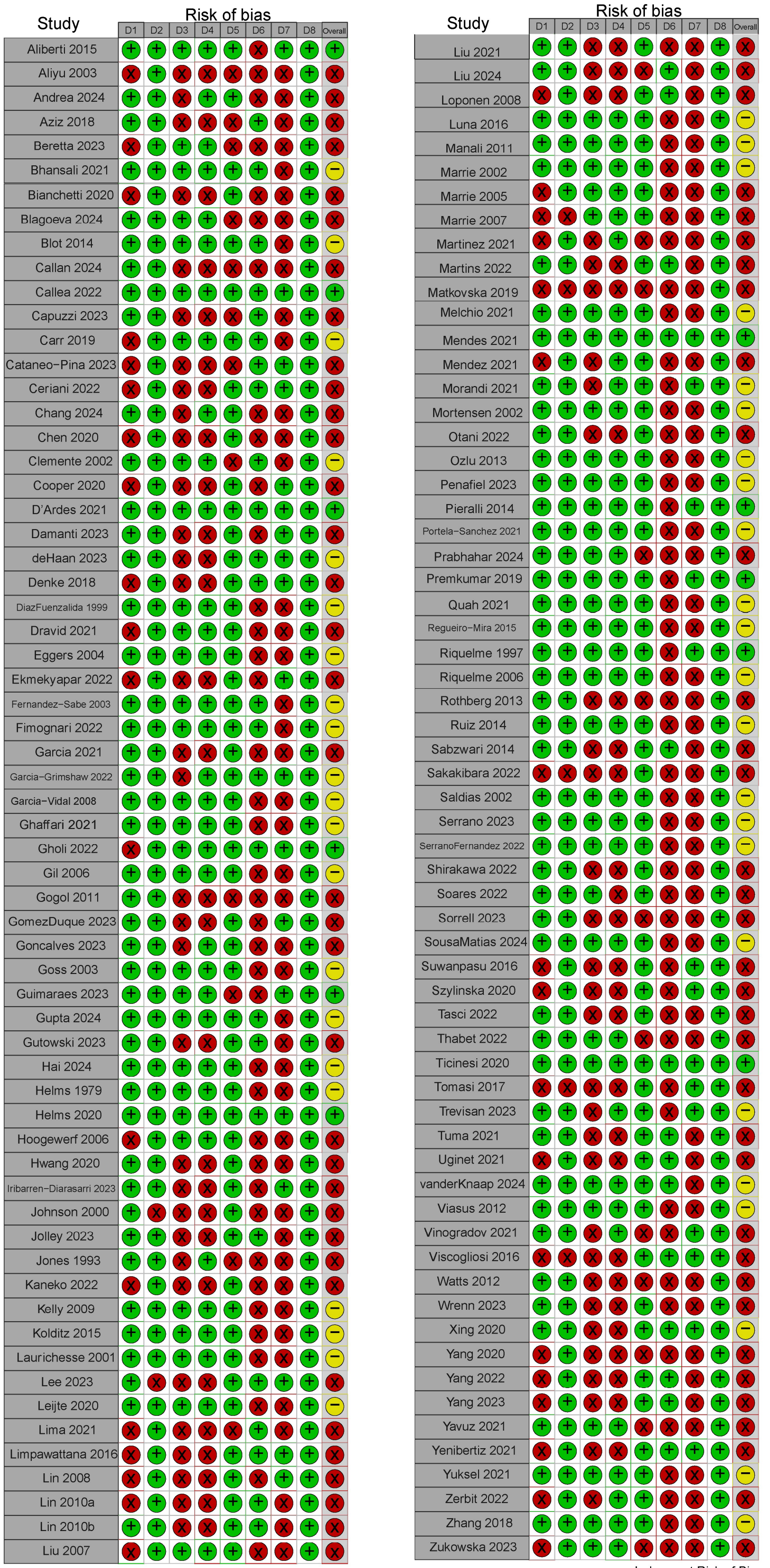


Judgement Risk of Bias
X High
− Moderate
+ Low

Appraisal Checklist domains
D1: Were the criteria for inclusion in the sample clearly defined (as close to all pneumonia patients as possible)?
D2: Were the study subjects and the setting described in detail?
D3: Was the exposure (pneumonia) measured in a valid & reliable way?
D4: Were objective, standard criteria used for measurement of the condition (pneumonia)?
D5: Were confounding factors identified (at least 2/4 of Age, Dementia, ICU, mechanical ventilation?
D6: Were strategies to deal with confounding factors stated?
D7: Were the outcomes (delirium) measured in a valid & reliable way (prospective DSM-based diagnosis by expert or validated clinical scale (CAM, CAM-ICU, ICDSC, 4AT, DRS, CHART-DEL, West Haven criteria)?
D8: Was the appropriate statistical analysis used?

**Figure S2. Funnel plots for all included studies reflect true heterogeneity rather than publication (selection) bias.**

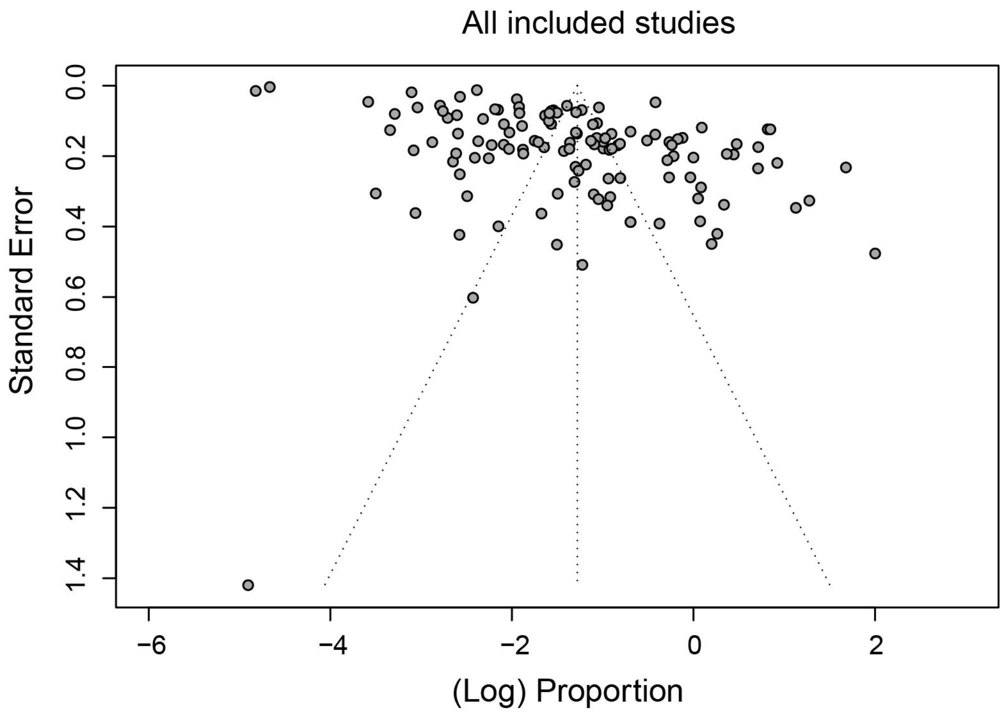


**A.** Assessment for potential publication bias, using 'funnel plot' of standard error by logistic event rate for all included studies.

#### Figure S3. Forest plot by risk of bias assessment.

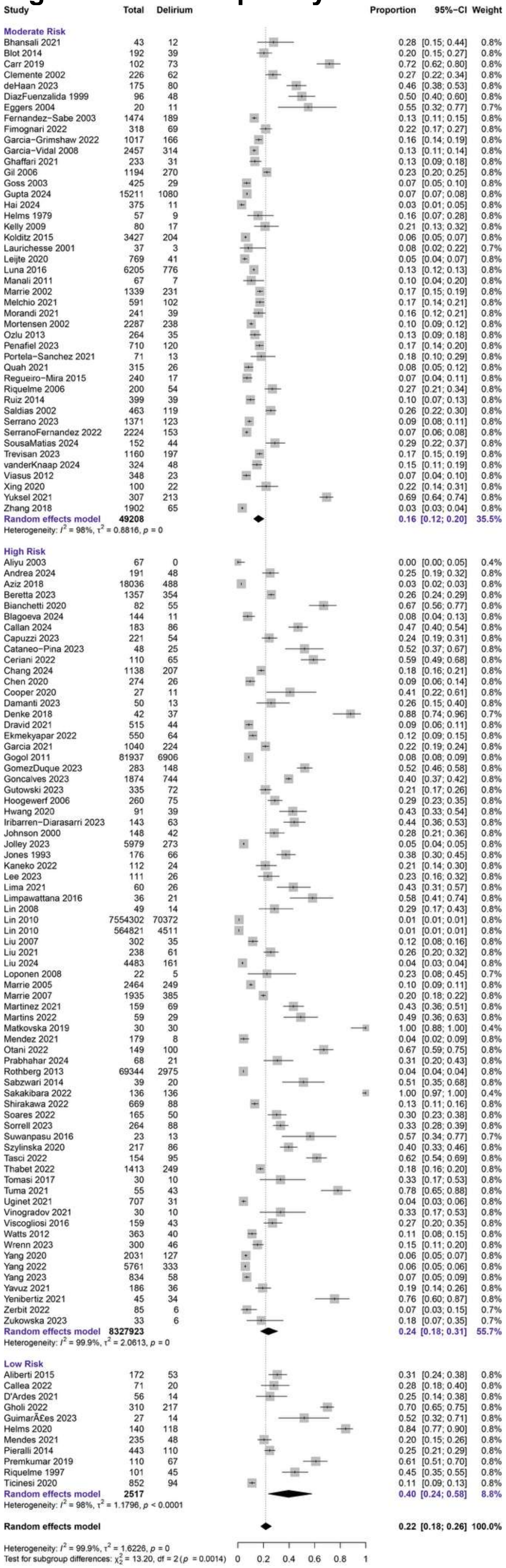

Delirium identification is increased in studies at low risk of bias using validated methods to diagnose delirium and pneumonia. Each paper was graded for risk of bias using the JBI manual for evidence synthesis tool across eight different quality measures (Supplementary Table S2 describes in detail the grading criteria used and Supplementary Figure S1 shows the grading for each primary study). Overall risk of bias was classified as “low” if studies met ≥6/8 domains including clear ascertainment of both pneumonia and delirium using standardized methods or validated clinical scores.

### Figure S4. Forest plot by delirium assessment method

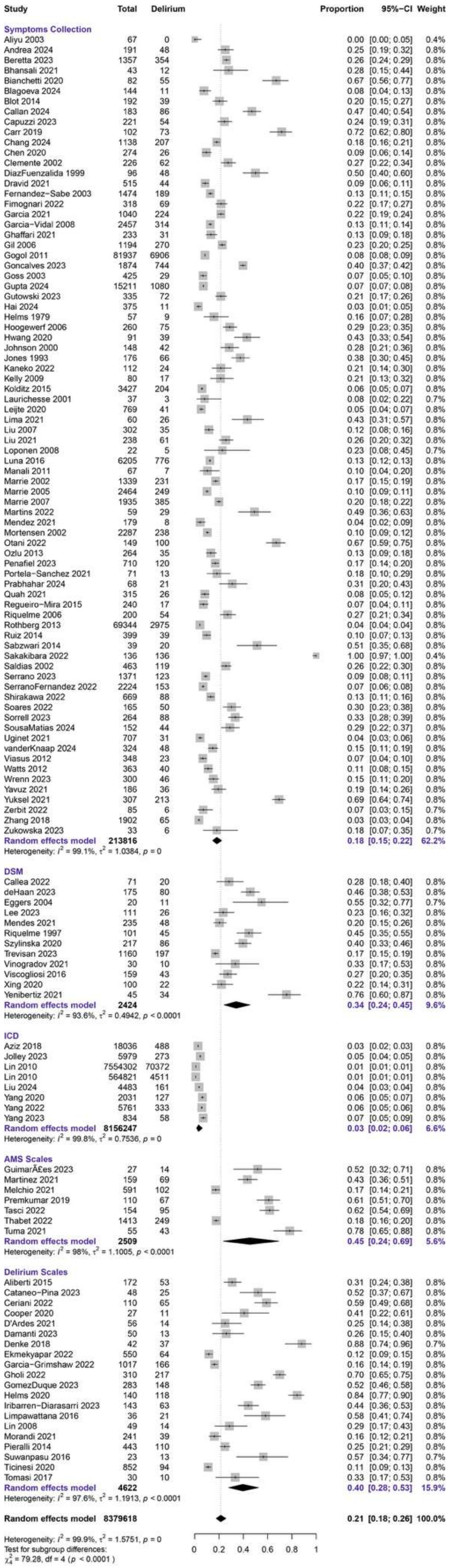

### Figure S5. Forest plot by Hospital Setting

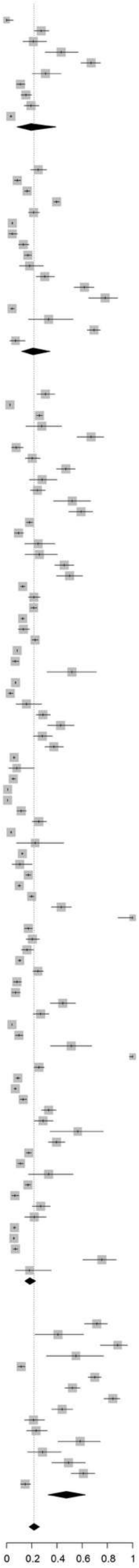

| Study | Total | Delirium | Proportion | 95%-CI | Weight |
|---|---|---|---|---|---|
| **ED** | | | | | |
| Aliyu 2003 | 67 | 0 | 0.00 | [0.00; 0.05] | 0.4% |
| Clemente 2002 | 226 | 62 | 0.27 | [0.22; 0.34] | 0.8% |
| Kelly 2009 | 80 | 17 | 0.21 | [0.13; 0.32] | 0.8% |
| Lima 2021 | 60 | 26 | 0.43 | [0.31; 0.57] | 0.8% |
| Otani 2022 | 149 | 100 | 0.67 | [0.59; 0.75] | 0.8% |
| Prabhahar 2024 | 68 | 21 | 0.31 | [0.20; 0.43] | 0.8% |
| Watts 2012 | 363 | 40 | 0.11 | [0.08; 0.15] | 0.8% |
| Wrenn 2023 | 300 | 46 | 0.15 | [0.11; 0.20] | 0.8% |
| Yavuz 2021 | 186 | 36 | 0.19 | [0.14; 0.26] | 0.8% |
| Zhang 2018 | 1902 | 65 | 0.03 | [0.03; 0.04] | 0.8% |
| **Random effects model** | **3401** | | **0.19** | **[0.08; 0.39]** | **7.6%** |
| Heterogeneity: $I^2 = 97.9\%$, $\tau^2 = 1.5174$, $p < 0.0001$ | | | | | |
| **Mixed (ICU & nonICU)** | | | | | |
| Andrea 2024 | 191 | 48 | 0.25 | [0.19; 0.32] | 0.8% |
| Dravid 2021 | 515 | 44 | 0.09 | [0.06; 0.11] | 0.8% |
| Garcia−Grimshaw 2022 | 1017 | 166 | 0.16 | [0.14; 0.19] | 0.8% |
| Goncalves 2023 | 1874 | 744 | 0.40 | [0.37; 0.42] | 0.8% |
| Gutowski 2023 | 335 | 72 | 0.21 | [0.17; 0.26] | 0.8% |
| Jolley 2023 | 5979 | 273 | 0.05 | [0.04; 0.05] | 0.8% |
| Mendez 2021 | 179 | 8 | 0.04 | [0.02; 0.09] | 0.8% |
| Ozlu 2013 | 264 | 35 | 0.13 | [0.09; 0.18] | 0.8% |
| Penafiel 2023 | 710 | 120 | 0.17 | [0.14; 0.20] | 0.8% |
| Portela−Sanchez 2021 | 71 | 13 | 0.18 | [0.10; 0.29] | 0.8% |
| Soares 2022 | 165 | 50 | 0.30 | [0.23; 0.38] | 0.8% |
| Tasci 2022 | 154 | 95 | 0.62 | [0.54; 0.69] | 0.8% |
| Tuma 2021 | 55 | 43 | 0.78 | [0.65; 0.88] | 0.8% |
| Uginet 2021 | 707 | 31 | 0.04 | [0.03; 0.06] | 0.8% |
| Vinogradov 2021 | 30 | 10 | 0.33 | [0.17; 0.53] | 0.8% |
| Yuksel 2021 | 307 | 213 | 0.69 | [0.64; 0.74] | 0.8% |
| Zerbit 2022 | 85 | 6 | 0.07 | [0.03; 0.15] | 0.7% |
| **Random effects model** | **12638** | | **0.21** | **[0.12; 0.35]** | **13.6%** |
| Heterogeneity: $I^2 = 99.1\%$, $\tau^2 = 1.7008$, $p = 0$ | | | | | |
| **Hospital wards** | | | | | |
| Aliberti 2015 | 172 | 53 | 0.31 | [0.24; 0.38] | 0.8% |
| Aziz 2018 | 18036 | 488 | 0.03 | [0.02; 0.03] | 0.8% |
| Beretta 2023 | 1357 | 354 | 0.26 | [0.24; 0.29] | 0.8% |
| Bhansali 2021 | 43 | 12 | 0.28 | [0.15; 0.44] | 0.8% |
| Bianchetti 2020 | 82 | 55 | 0.67 | [0.56; 0.77] | 0.8% |
| Blagoeva 2024 | 144 | 11 | 0.08 | [0.04; 0.13] | 0.8% |
| Blot 2014 | 192 | 39 | 0.20 | [0.15; 0.27] | 0.8% |
| Callan 2024 | 183 | 86 | 0.47 | [0.40; 0.54] | 0.8% |
| Callea 2022 | 71 | 20 | 0.28 | [0.18; 0.40] | 0.8% |
| Capuzzi 2023 | 221 | 54 | 0.24 | [0.19; 0.31] | 0.8% |
| Cataneo−Pina 2023 | 48 | 25 | 0.52 | [0.37; 0.67] | 0.8% |
| Ceriani 2022 | 110 | 65 | 0.59 | [0.49; 0.68] | 0.8% |
| Chang 2024 | 1138 | 207 | 0.18 | [0.16; 0.21] | 0.8% |
| Chen 2020 | 274 | 26 | 0.09 | [0.06; 0.14] | 0.8% |
| D'Ardes 2021 | 56 | 14 | 0.25 | [0.14; 0.38] | 0.8% |
| Damanti 2023 | 50 | 13 | 0.26 | [0.15; 0.40] | 0.8% |
| deHaan 2023 | 175 | 80 | 0.46 | [0.38; 0.53] | 0.8% |
| DiazFuenzalida 1999 | 96 | 48 | 0.50 | [0.40; 0.60] | 0.8% |
| Fernandez−Sabe 2003 | 1474 | 189 | 0.13 | [0.11; 0.15] | 0.8% |
| Fimognari 2022 | 318 | 69 | 0.22 | [0.17; 0.27] | 0.8% |
| Garcia 2021 | 1040 | 224 | 0.22 | [0.19; 0.24] | 0.8% |
| Garcia−Vidal 2008 | 2457 | 314 | 0.13 | [0.11; 0.14] | 0.8% |
| Ghaffari 2021 | 233 | 31 | 0.13 | [0.09; 0.18] | 0.8% |
| Gil 2006 | 1194 | 270 | 0.23 | [0.20; 0.25] | 0.8% |
| Gogol 2011 | 81937 | 6906 | 0.08 | [0.08; 0.09] | 0.8% |
| Goss 2003 | 425 | 29 | 0.07 | [0.05; 0.10] | 0.8% |
| Guimaraes 2023 | 27 | 14 | 0.52 | [0.32; 0.71] | 0.8% |
| Gupta 2024 | 15211 | 1080 | 0.07 | [0.07; 0.08] | 0.8% |
| Hai 2024 | 375 | 11 | 0.03 | [0.01; 0.05] | 0.8% |
| Helms 1979 | 57 | 9 | 0.16 | [0.07; 0.28] | 0.8% |
| Hoogewerf 2006 | 260 | 75 | 0.29 | [0.23; 0.35] | 0.8% |
| Hwang 2020 | 91 | 39 | 0.43 | [0.33; 0.54] | 0.8% |
| Johnson 2000 | 148 | 42 | 0.28 | [0.21; 0.36] | 0.8% |
| Jones 1993 | 176 | 66 | 0.38 | [0.30; 0.45] | 0.8% |
| Kolditz 2015 | 3427 | 204 | 0.06 | [0.05; 0.07] | 0.8% |
| Laurichesse 2001 | 37 | 3 | 0.08 | [0.02; 0.22] | 0.7% |
| Leijte 2020 | 769 | 41 | 0.05 | [0.04; 0.07] | 0.8% |
| Lin 2010 | 7554302 | 70372 | 0.01 | [0.01; 0.01] | 0.8% |
| Lin 2010 | 564821 | 4511 | 0.01 | [0.01; 0.01] | 0.8% |
| Liu 2007 | 302 | 35 | 0.12 | [0.08; 0.16] | 0.8% |
| Liu 2021 | 238 | 61 | 0.26 | [0.20; 0.32] | 0.8% |
| Liu 2024 | 4483 | 161 | 0.04 | [0.03; 0.04] | 0.8% |
| Loponen 2008 | 22 | 5 | 0.23 | [0.08; 0.45] | 0.7% |
| Luna 2016 | 6205 | 776 | 0.13 | [0.12; 0.13] | 0.8% |
| Manali 2011 | 67 | 7 | 0.10 | [0.04; 0.20] | 0.8% |
| Marrie 2002 | 1339 | 231 | 0.17 | [0.15; 0.19] | 0.8% |
| Marrie 2005 | 2464 | 249 | 0.10 | [0.09; 0.11] | 0.8% |
| Marrie 2007 | 1935 | 385 | 0.20 | [0.18; 0.22] | 0.8% |
| Martinez 2021 | 159 | 69 | 0.43 | [0.36; 0.51] | 0.8% |
| Matkovska 2019 | 30 | 30 | 1.00 | [0.88; 1.00] | 0.4% |
| Melchio 2021 | 591 | 102 | 0.17 | [0.14; 0.21] | 0.8% |
| Mendes 2021 | 235 | 48 | 0.20 | [0.15; 0.26] | 0.8% |
| Morandi 2021 | 241 | 39 | 0.16 | [0.12; 0.21] | 0.8% |
| Mortensen 2002 | 2287 | 238 | 0.10 | [0.09; 0.12] | 0.8% |
| Pieralli 2014 | 443 | 110 | 0.25 | [0.21; 0.29] | 0.8% |
| Quah 2021 | 315 | 26 | 0.08 | [0.05; 0.12] | 0.8% |
| Regueiro−Mira 2015 | 240 | 17 | 0.07 | [0.04; 0.11] | 0.8% |
| Riquelme 1997 | 101 | 45 | 0.45 | [0.35; 0.55] | 0.8% |
| Riquelme 2006 | 200 | 54 | 0.27 | [0.21; 0.34] | 0.8% |
| Rothberg 2013 | 69344 | 2975 | 0.04 | [0.04; 0.04] | 0.8% |
| Ruiz 2014 | 399 | 39 | 0.10 | [0.07; 0.13] | 0.8% |
| Sabzwari 2014 | 39 | 20 | 0.51 | [0.35; 0.68] | 0.8% |
| Sakakibara 2022 | 136 | 136 | 1.00 | [0.97; 1.00] | 0.4% |
| Saldias 2002 | 463 | 119 | 0.26 | [0.22; 0.30] | 0.8% |
| Serrano 2023 | 1371 | 123 | 0.09 | [0.08; 0.11] | 0.8% |
| SerranoFernandez 2022 | 2224 | 153 | 0.07 | [0.06; 0.08] | 0.8% |
| Shirakawa 2022 | 669 | 88 | 0.13 | [0.11; 0.16] | 0.8% |
| Sorrell 2023 | 264 | 88 | 0.33 | [0.28; 0.39] | 0.8% |
| SousaMatias 2024 | 152 | 44 | 0.29 | [0.22; 0.37] | 0.8% |
| Suwanpasu 2016 | 23 | 13 | 0.57 | [0.34; 0.77] | 0.7% |
| Szylinska 2020 | 217 | 86 | 0.40 | [0.33; 0.46] | 0.8% |
| Thabet 2022 | 1413 | 249 | 0.18 | [0.16; 0.20] | 0.8% |
| Ticinesi 2020 | 852 | 94 | 0.11 | [0.09; 0.13] | 0.8% |
| Tomasi 2017 | 30 | 10 | 0.33 | [0.17; 0.53] | 0.8% |
| Trevisan 2023 | 1160 | 197 | 0.17 | [0.15; 0.19] | 0.8% |
| Viasus 2012 | 348 | 23 | 0.07 | [0.04; 0.10] | 0.8% |
| Viscogliosi 2016 | 159 | 43 | 0.27 | [0.20; 0.35] | 0.8% |
| Xing 2020 | 100 | 22 | 0.22 | [0.14; 0.31] | 0.8% |
| Yang 2020 | 2031 | 127 | 0.06 | [0.05; 0.07] | 0.8% |
| Yang 2022 | 5761 | 333 | 0.06 | [0.05; 0.06] | 0.8% |
| Yang 2023 | 834 | 58 | 0.07 | [0.05; 0.09] | 0.8% |
| Yenibertiz 2021 | 45 | 34 | 0.76 | [0.60; 0.87] | 0.8% |
| Zukowska 2023 | 33 | 6 | 0.18 | [0.07; 0.35] | 0.7% |
| **Random effects model** | **8361191** | | **0.18** | **[0.15; 0.23]** | **66.0%** |
| Heterogeneity: $I^2 = 99.9\%$, $\tau^2 = 1.3914$, $p = 0$ | | | | | |
| **ICU** | | | | | |
| Carr 2019 | 102 | 73 | 0.72 | [0.62; 0.80] | 0.8% |
| Cooper 2020 | 27 | 11 | 0.41 | [0.22; 0.61] | 0.8% |
| Denke 2018 | 42 | 37 | 0.88 | [0.74; 0.96] | 0.7% |
| Eggers 2004 | 20 | 11 | 0.55 | [0.32; 0.77] | 0.7% |
| Ekmekyapar 2022 | 550 | 64 | 0.12 | [0.09; 0.15] | 0.8% |
| Gholi 2022 | 310 | 217 | 0.70 | [0.65; 0.75] | 0.8% |
| GomezDuque 2023 | 283 | 148 | 0.52 | [0.46; 0.58] | 0.8% |
| Helms 2020 | 140 | 118 | 0.84 | [0.77; 0.90] | 0.8% |
| Iribarren−Diarasarri 2023 | 143 | 63 | 0.44 | [0.36; 0.53] | 0.8% |
| Kaneko 2022 | 112 | 24 | 0.21 | [0.14; 0.30] | 0.8% |
| Lee 2023 | 111 | 26 | 0.23 | [0.16; 0.32] | 0.8% |
| Limpawattana 2016 | 36 | 21 | 0.58 | [0.41; 0.74] | 0.8% |
| Lin 2008 | 49 | 14 | 0.29 | [0.17; 0.43] | 0.8% |
| Martins 2022 | 59 | 29 | 0.49 | [0.36; 0.63] | 0.8% |
| Premkumar 2019 | 110 | 67 | 0.61 | [0.51; 0.70] | 0.8% |
| vanderKnaap 2024 | 324 | 48 | 0.15 | [0.11; 0.19] | 0.8% |
| **Random effects model** | **2418** | | **0.47** | **[0.33; 0.63]** | **12.7%** |
| Heterogeneity: $I^2 = 97.2\%$, $\tau^2 = 1.2525$, $p < 0.0001$ | | | | | |
| **Random effects model** | | | **0.22** | **[0.18; 0.26]** | **100.0%** |

Heterogeneity: $I^2 = 99.9\%$, $\tau^2 = 1.6226$, $p = 0$
Test for subgroup differences:
$\chi^2_3 = 18.96$, df = 3 ($p = 0.0003$)

#### Figure S6. Forest plot by Pneumonia Infection Origin

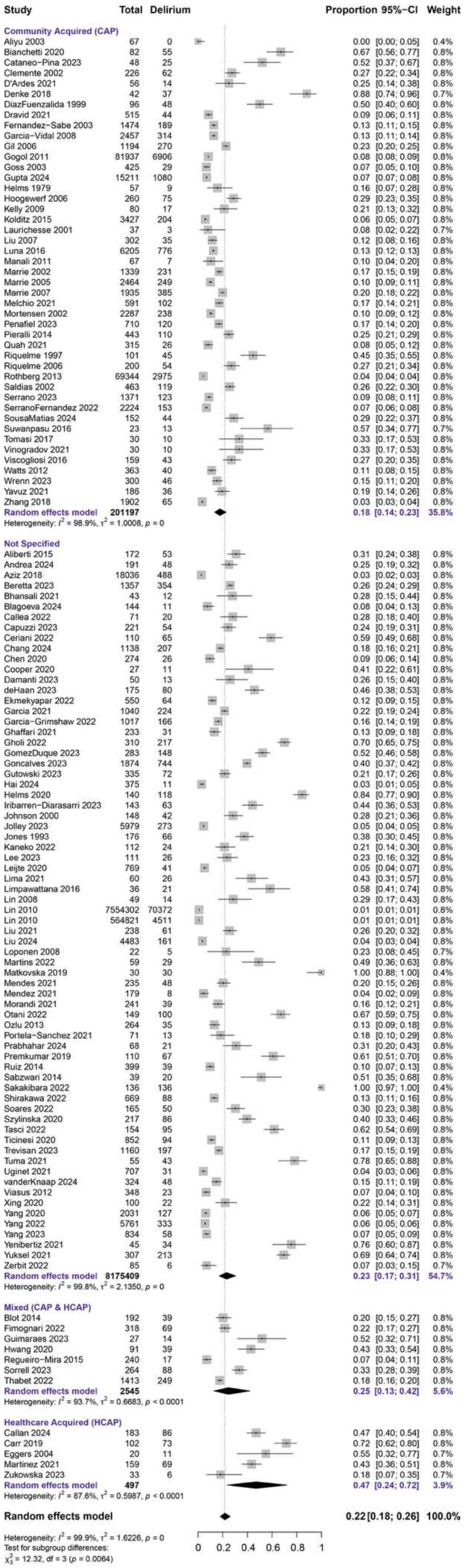

#### Figure S7. Forest plot by Microbiological etiology

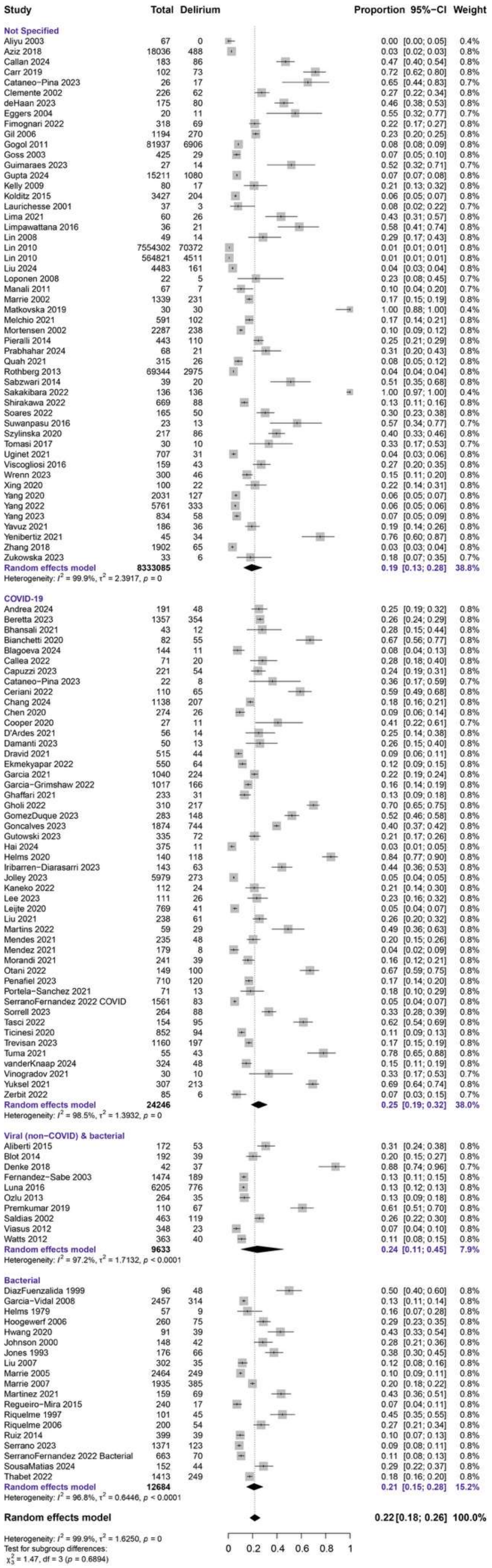

**Figure S8. Subgroup analysis in studies at low risk of bias**

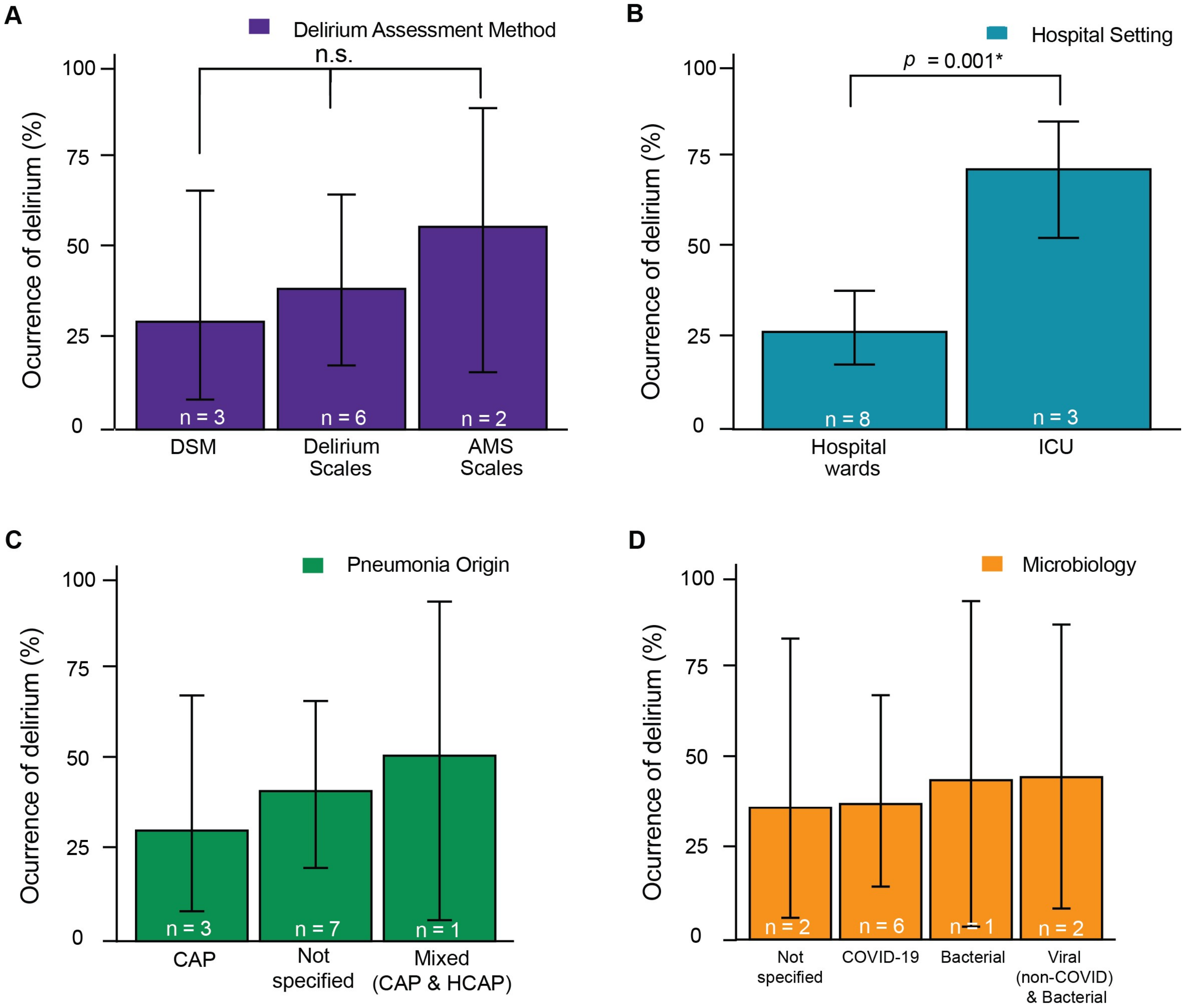


**A.** Ascertainment of delirium through validated methods was a criterion to classify studies as low risk of bias. As expected, delirium rates did not vary significantly according to the validated assessment method used in these studies ($R^2 = 0\%$, $p = 0.60$). Compared to the DSM, delirium rates were similar when assessed using standardized assessments of mental status (AMS Scales, $p$=0.98) and with validated Delirium scales ($p$=0.59). Standardized assessments of mental status included the Glasgow Coma Scale, Richmond Agitation Sedation Scale, Abbreviated Mental Test, and West-Haven criteria for hepatic encephalopathy. Delirium scales included CAM, CAM-ICU, ICSDC, 4AT, DRS, DOS, and CHART-DEL (see abbreviations in Supplementary Methods). Each bar represents a meta-analytic estimate of delirium rates, with the calculated 95% confidence interval. Post-hoc $p$-values in all panels are adjusted for multiple comparisons (Holm). Forest plots of all studies with N, meta-analytic proportion, CI and heterogeneity measures per each subgroup analysis are provided in Supplementary Figure S9A.

**B.** In studies at low risk of bias, delirium rates varied significantly according to study setting, which explained 68.2% of variance in delirium rates across studies ($R^2$ 68.2%, $p$=0.0015). Delirium rates were significantly higher for studies performed in the Intensive Care Unit (ICU) compared to those in hospital wards ($p$=0.001). Conventions as in A, with forest plots in Supplementary figure S9B).

**C.** Delirium rates did not vary significantly according to pneumonia origin in studies at low risk of bias, which 0% of variance in delirium rates across studies ($R^2$ = 0%, $p$=0.77). (CAP=Community Acquired Pneumonia, HCAP=Healthcare Acquired Pneumonia). Conventions as in A, with forest plots in Supplementary Figure S9C).
**D.** Delirium rates did not vary significantly according to microbiological etiologies in studies at low risk of bias, which explained 0.0% of variance in delirium rates across studies ($R^2$ = 0.0%, $p$=0.98). Conventions as in A, with forest plots in Supplementary Figure 9D).

#### Figure S9. Forest plots per subgroup analysis in studies at low risk of bias

##### A. Delirium Assessment method

| Study | Total | Delirium | Proportion | 95%−CI | Weight |
|---|---|---|---|---|---|
| **Delirium scale** | | | | | |
| Aliberti 2015 | 172 | 53 | 0.31 | [0.24; 0.38] | 9.2% |
| D'Ardes 2021 | 56 | 14 | 0.25 | [0.14; 0.38] | 8.8% |
| Gholi 2022 | 310 | 217 | 0.70 | [0.65; 0.75] | 9.3% |
| Helms 2020 | 140 | 118 | 0.84 | [0.77; 0.90] | 9.0% |
| Pieralli 2014 | 443 | 110 | 0.25 | [0.21; 0.29] | 9.4% |
| Ticinesi 2020 | 852 | 94 | 0.11 | [0.09; 0.13] | 9.4% |
| **Random effects model** | **1973** | | **0.39** | **[0.13; 0.74]** | **55.1%** |
| Heterogeneity: $I^2 = 98.9\%$, $\tau^2 = 1.9413$, $p < 0.0001$ | | | | | |
| **DSM** | | | | | |
| Callea 2022 | 71 | 20 | 0.28 | [0.18; 0.40] | 8.9% |
| Mendes 2021 | 235 | 48 | 0.20 | [0.15; 0.26] | 9.3% |
| Riquelme 1997 | 101 | 45 | 0.45 | [0.35; 0.55] | 9.1% |
| **Random effects model** | **407** | | **0.30** | **[0.09; 0.65]** | **27.3%** |
| Heterogeneity: $I^2 = 89.8\%$, $\tau^2 = 0.3081$, $p < 0.0001$ | | | | | |
| **AMS** | | | | | |
| Guimaraes 2023 | 27 | 14 | 0.52 | [0.32; 0.71] | 8.4% |
| Premkumar 2019 | 110 | 67 | 0.61 | [0.51; 0.70] | 9.2% |
| **Random effects model** | **137** | | **0.59** | **[0.18; 0.91]** | **17.6%** |
| Heterogeneity: $I^2 = 0\%$, $\tau^2 = 0$, $p = 0.3924$ | | | | | |
| **Random effects model** | **2517** | | **0.40** | **[0.24; 0.58]** | **100.0%** |

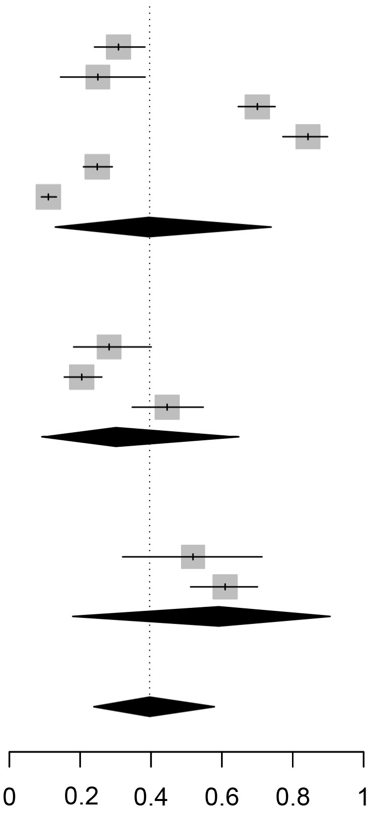


Heterogeneity: $I^2 = 98.0\%$, $\tau^2 = 1.1796$, $p < 0.0001$
Test for subgroup differences: $\chi^2_2 = 11.80$, df = 2 ($p = 0.0027$)

##### B. Hospital Setting

| Study | Total | Delirium | Proportion | 95%−CI | Weight |
|---|---|---|---|---|---|
| **Hospital wards** | | | | | |
| Aliberti 2015 | 172 | 53 | 0.31 | [0.24; 0.38] | 9.2% |
| Callea 2022 | 71 | 20 | 0.28 | [0.18; 0.40] | 8.9% |
| D'Ardes 2021 | 56 | 14 | 0.25 | [0.14; 0.38] | 8.8% |
| Guimaraes 2023 | 27 | 14 | 0.52 | [0.32; 0.71] | 8.4% |
| Mendes 2021 | 235 | 48 | 0.20 | [0.15; 0.26] | 9.3% |
| Pieralli 2014 | 443 | 110 | 0.25 | [0.21; 0.29] | 9.4% |
| Riquelme 1997 | 101 | 45 | 0.45 | [0.35; 0.55] | 9.1% |
| Ticinesi 2020 | 852 | 94 | 0.11 | [0.09; 0.13] | 9.4% |
| **Random effects model** | **1957** | | **0.27** | **[0.18; 0.39]** | **72.5%** |
| Heterogeneity: $I^2 = 93.5\%$, $\tau^2 = 0.3853$, $p < 0.0001$ | | | | | |
| **ICU** | | | | | |
| Gholi 2022 | 310 | 217 | 0.70 | [0.65; 0.75] | 9.3% |
| Helms 2020 | 140 | 118 | 0.84 | [0.77; 0.90] | 9.0% |
| Premkumar 2019 | 110 | 67 | 0.61 | [0.51; 0.70] | 9.2% |
| **Random effects model** | **560** | | **0.73** | **[0.36; 0.93]** | **27.5%** |
| Heterogeneity: $I^2 = 88.2\%$, $\tau^2 = 0.3384$, $p = 0.0002$ | | | | | |
| **Random effects model** | **2517** | | **0.40** | **[0.24; 0.58]** | **100.0%** |

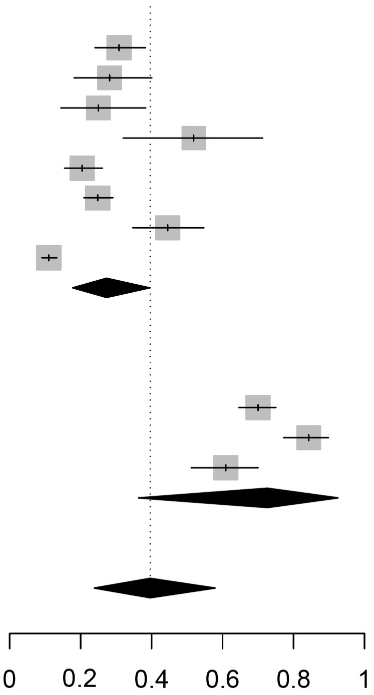


Heterogeneity: $I^2 = 98.0\%$, $\tau^2 = 1.1796$, $p < 0.0001$
Test for subgroup differences: $\chi^2_1 = 20.96$, df = 1 ($p < 0.0001$)

#### Figure S9. Forest plots per subgroup analysis in studies at low risk of bias. *(continued)*

##### C. Pneumonia Infection origin

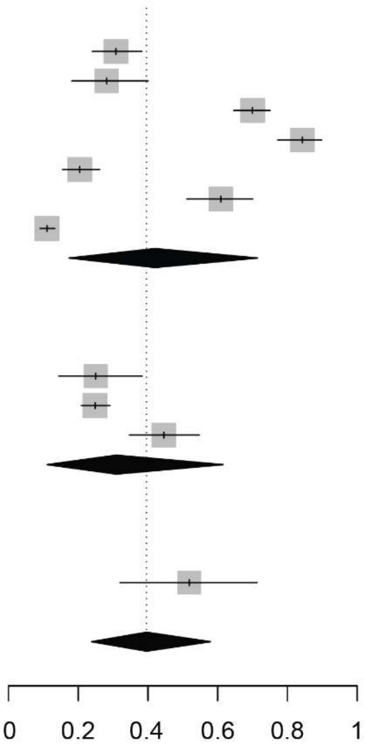

| Study | Total | Delirium | Proportion | 95%−CI | Weight |
|---|---|---|---|---|---|
| **Not Specified** | | | | | |
| Aliberti 2015 | 172 | 53 | 0.31 | [0.24; 0.38] | 9.2% |
| Callea 2022 | 71 | 20 | 0.28 | [0.18; 0.40] | 8.9% |
| Gholi 2022 | 310 | 217 | 0.70 | [0.65; 0.75] | 9.3% |
| Helms 2020 | 140 | 118 | 0.84 | [0.77; 0.90] | 9.0% |
| Mendes 2021 | 235 | 48 | 0.20 | [0.15; 0.26] | 9.3% |
| Premkumar 2019 | 110 | 67 | 0.61 | [0.51; 0.70] | 9.2% |
| Ticinesi 2020 | 852 | 94 | 0.11 | [0.09; 0.13] | 9.4% |
| **Random effects model** | **1890** | | **0.42** | **[0.17; 0.71]** | **64.3%** |
| Heterogeneity: $I^2 = 98.7\%$, $\tau^2 = 1.7633$, $p < 0.0001$ | | | | | |
| **Community Acquired (CAP)** | | | | | |
| D'Ardes 2021 | 56 | 14 | 0.25 | [0.14; 0.38] | 8.8% |
| Pieralli 2014 | 443 | 110 | 0.25 | [0.21; 0.29] | 9.4% |
| Riquelme 1997 | 101 | 45 | 0.45 | [0.35; 0.55] | 9.1% |
| **Random effects model** | **600** | | **0.31** | **[0.11; 0.62]** | **27.3%** |
| Heterogeneity: $I^2 = 87.1\%$, $\tau^2 = 0.2253$, $p = 0.0004$ | | | | | |
| **Mixed (CAP & HCAP)** | | | | | |
| Guimaraes 2023 | 27 | 14 | **0.52** | **[0.32; 0.71]** | **8.4%** |
| **Random effects model** | **2517** | | **0.40** | **[0.24; 0.58]** | **100.0%** |

Heterogeneity: $I^2 = 98.0\%$, $\tau^2 = 1.1796$, $p < 0.0001$
Test for subgroup differences: $\chi^2_2 = 3.34$, df = 2 ($p = 0.1885$)

##### D. Microbiological etiology

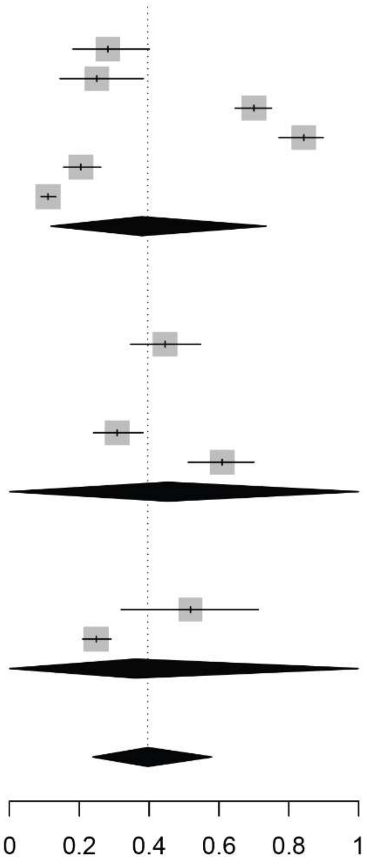

| Study | Total | Delirium | Proportion | 95%−CI | Weight |
|---|---|---|---|---|---|
| **COVID-19** | | | | | |
| Callea 2022 | 71 | 20 | 0.28 | [0.18; 0.40] | 8.9% |
| D'Ardes 2021 | 56 | 14 | 0.25 | [0.14; 0.38] | 8.8% |
| Gholi 2022 | 310 | 217 | 0.70 | [0.65; 0.75] | 9.3% |
| Helms 2020 | 140 | 118 | 0.84 | [0.77; 0.90] | 9.0% |
| Mendes 2021 | 235 | 48 | 0.20 | [0.15; 0.26] | 9.3% |
| Ticinesi 2020 | 852 | 94 | 0.11 | [0.09; 0.13] | 9.4% |
| **Random effects model** | **1664** | | **0.38** | **[0.12; 0.74]** | **54.7%** |
| Heterogeneity: $I^2 = 98.9\%$, $\tau^2 = 2.0470$, $p < 0.0001$ | | | | | |
| **Bacterial** | | | | | |
| Riquelme 1997 | 101 | 45 | **0.45** | **[0.35; 0.55]** | **9.1%** |
| **Viral (non−COVID) & bacterial** | | | | | |
| Aliberti 2015 | 172 | 53 | 0.31 | [0.24; 0.38] | 9.2% |
| Premkumar 2019 | 110 | 67 | 0.61 | [0.51; 0.70] | 9.2% |
| **Random effects model** | **282** | | **0.45** | **[0.00; 1.00]** | **18.4%** |
| Heterogeneity: $I^2 = 95.8\%$, $\tau^2 = 0.7514$, $p < 0.0001$ | | | | | |
| **Not Specified** | | | | | |
| Guimaraes 2023 | 27 | 14 | 0.52 | [0.32; 0.71] | 8.4% |
| Pieralli 2014 | 443 | 110 | 0.25 | [0.21; 0.29] | 9.4% |
| **Random effects model** | **470** | | **0.36** | **[0.00; 1.00]** | **17.8%** |
| Heterogeneity: $I^2 = 88.5\%$, $\tau^2 = 0.6181$, $p = 0.0032$ | | | | | |
| **Random effects model** | **2517** | | **0.40** | **[0.24; 0.58]** | **100.0%** |

Heterogeneity: $I^2 = 98.0\%$, $\tau^2 = 1.1796$, $p < 0.0001$
Test for subgroup differences: $\chi^2_3 = 0.49$, df = 3 ($p = 0.9201$)

**Figure S10. Older age is a predisposing factor for delirium.**

**A**

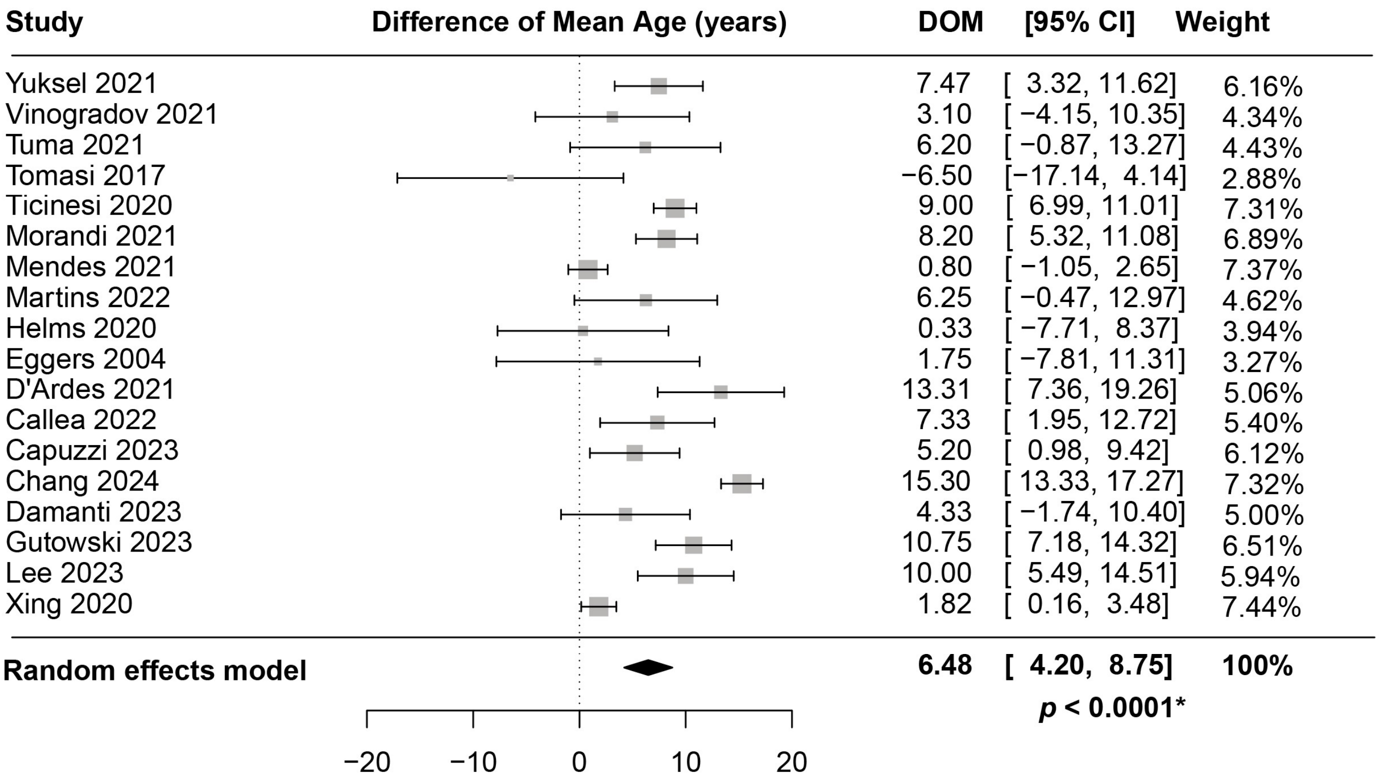


**B**

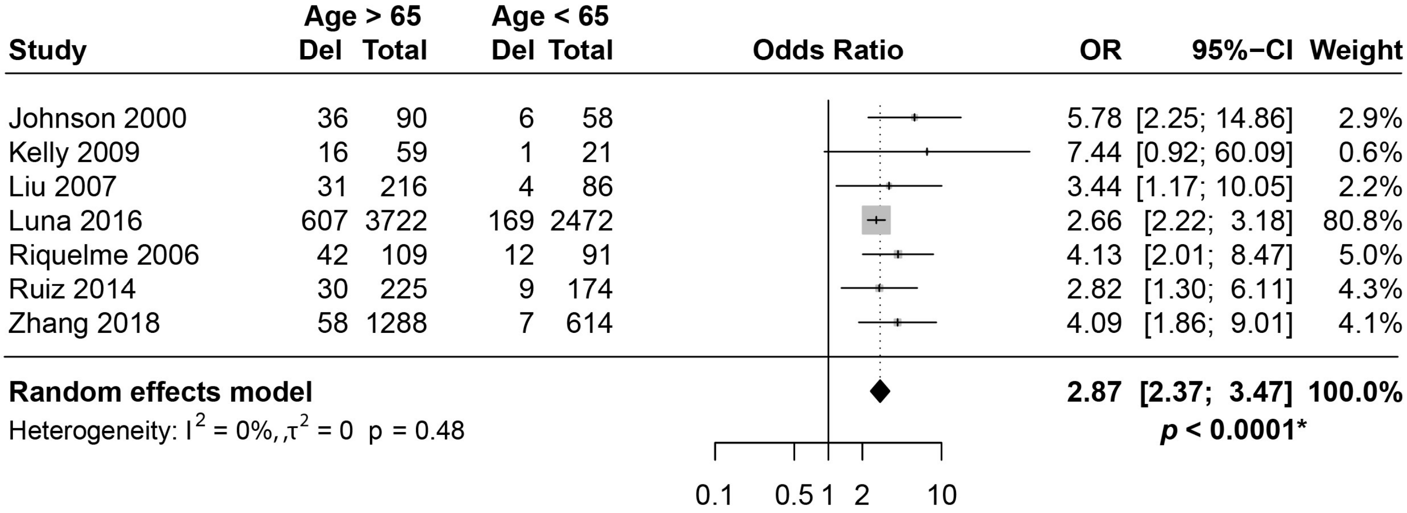


**A.** Patients with pneumonia and delirium are older than those without delirium (n=18 studies, DOM +6.5 years, 95% CI [4.2; 8.7], $p$<0.0001). **B.** Subgroup analysis by age showed that patients >65 years-old had increased odds of delirium compared to those <65 years old ($n$=7 studies, OR 2.87, 95% CI [2.37;3.47], p<0.0001).

#### Figure S11. Forest plots by demographics as predisposing factors of delirium

##### A. Gender (female)

| Study | Delirium Events | Delirium Total | No Del Events | No Del Total | OR | 95%-CI | Weight |
|---|---|---|---|---|---|---|---|
| Callea 2022 | 9 | 20 | 21 | 51 | 1.17 | [0.41; 3.32] | 3.5% |
| Capuzzi 2023 | 12 | 54 | 46 | 167 | 0.75 | [0.36; 1.55] | 5.5% |
| Chang 2024 | 109 | 207 | 399 | 931 | 1.48 | [1.10; 2.01] | 9.9% |
| D'Ardes 2021 | 9 | 14 | 28 | 42 | 0.90 | [0.25; 3.20] | 2.6% |
| Damanti 2023 | 5 | 13 | 23 | 37 | 0.38 | [0.10; 1.40] | 2.5% |
| Eggers 2004 | 2 | 11 | 1 | 9 | 1.78 | [0.13; 23.52] | 0.8% |
| Garcia 2021 | 58 | 224 | 315 | 816 | 0.56 | [0.40; 0.77] | 9.5% |
| Gutowski 2023 | 42 | 72 | 99 | 263 | 2.32 | [1.36; 3.94] | 7.3% |
| Helms 2020 | 29 | 118 | 11 | 22 | 0.33 | [0.13; 0.83] | 4.1% |
| Lee 2023 | 11 | 26 | 41 | 85 | 0.79 | [0.32; 1.91] | 4.4% |
| Martins 2022 | 11 | 29 | 9 | 30 | 1.43 | [0.48; 4.21] | 3.3% |
| Mendes 2021 | 24 | 48 | 109 | 187 | 0.72 | [0.38; 1.35] | 6.3% |
| Morandi 2021 | 21 | 39 | 77 | 202 | 1.89 | [0.95; 3.78] | 5.8% |
| Portela−Sanchez 2021 | 3 | 13 | 18 | 58 | 0.67 | [0.16; 2.72] | 2.2% |
| Tasci 2022 | 33 | 95 | 22 | 59 | 0.90 | [0.46; 1.76] | 5.9% |
| Ticinesi 2020 | 42 | 94 | 360 | 758 | 0.89 | [0.58; 1.37] | 8.4% |
| Tomasi 2017 | 4 | 10 | 13 | 20 | 0.36 | [0.08; 1.71] | 1.9% |
| Tuma 2021 | 14 | 43 | 6 | 12 | 0.48 | [0.13; 1.77] | 2.5% |
| Vinogradov 2021 | 3 | 10 | 11 | 20 | 0.35 | [0.07; 1.76] | 1.8% |
| Xing 2020 | 12 | 22 | 40 | 78 | 1.14 | [0.44; 2.95] | 4.0% |
| Yuksel 2021 | 92 | 213 | 33 | 94 | 1.41 | [0.85; 2.32] | 7.6% |
| **Random effects model** | | **1375** | | **3941** | **0.93** | **[0.73; 1.20]** | **100.0%** |

Heterogeneity: $I^2 = 59\%$, $\tau^2 = 0.1202$, $p < 0.01$

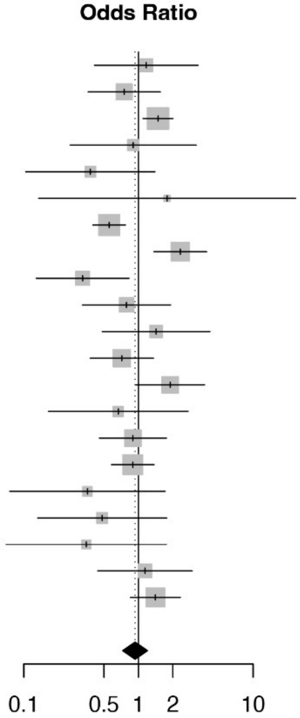


***p* = 0.57**

##### B. Nursing Home

| Study | Delirium Events | Delirium Total | NoDel Events | NoDel Total | OR | 95%-CI | Weight |
|---|---|---|---|---|---|---|---|
| Chang 2024 | 141 | 207 | 178 | 931 | 9.04 | [6.47; 12.63] | 50.5% |
| Damanti 2023 | 1 | 13 | 1 | 37 | 3.00 | [0.17; 51.75] | 8.6% |
| Mendes 2021 | 16 | 48 | 33 | 187 | 2.33 | [1.15; 4.74] | 40.9% |
| **Random effects model** | | **268** | | **1155** | **4.73** | **[0.64; 35.04]** | **100.0%** |

Heterogeneity: $I^2 = 83\%$, $\tau^2 = 0.4001$, $p < 0.01$

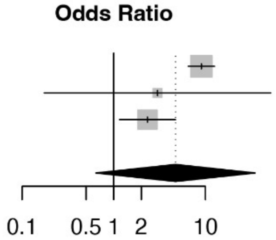


***p* = 0.08**

##### C. Smoking

| Study | Delirium Events | Delirium Total | NoDel Events | NoDel Total | OR | 95%-CI | Weight |
|---|---|---|---|---|---|---|---|
| Garcia−Grimshaw 2022 | 21 | 166 | 129 | 851 | 0.81 | [0.49; 1.33] | 45.4% |
| Gutowski 2023 | 5 | 72 | 22 | 263 | 0.82 | [0.30; 2.24] | 10.9% |
| Lee 2023 | 7 | 26 | 27 | 85 | 0.79 | [0.30; 2.11] | 11.6% |
| Morandi 2021 | 8 | 39 | 53 | 202 | 0.73 | [0.31; 1.68] | 15.8% |
| Tuma 2021 | 10 | 43 | 2 | 12 | 1.52 | [0.28; 8.09] | 4.0% |
| Xing 2020 | 11 | 22 | 42 | 78 | 0.86 | [0.33; 2.21] | 12.4% |
| **Random effects model** | | **368** | | **1491** | **0.82** | **[0.70; 0.96]** | **100.0%** |

Heterogeneity: $I^2 = 0\%$, $\tau^2 = 0$, $p = 0.99$

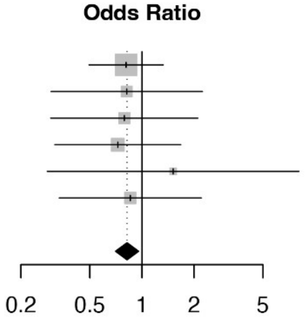


***p* = 0.02***

##### D. Alcohol intake

| Study | Delirium Events | Delirium Total | NoDel Events | NoDel Total | OR | 95%-CI | Weight |
|---|---|---|---|---|---|---|---|
| Jolley 2023 | 23 | 273 | 221 | 5706 | 2.28 | [1.46; 3.57] | 80.3% |
| Tuma 2021 | 4 | 43 | 0 | 12 | 2.85 | [0.14; 56.65] | 1.8% |
| Xing 2020 | 12 | 22 | 38 | 78 | 1.26 | [0.49; 3.26] | 17.9% |
| **Random effects model** | | **338** | | **5796** | **2.06** | **[1.02; 4.16]** | **100.0%** |

Heterogeneity: $I^2 = 0\%$, $\tau^2 = 0$, $p = 0.53$

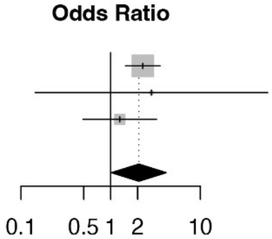


***p* = 0.047***

#### Figure S12. Forest plots by comorbidities as predisposing factors of delirium

##### A. Dementia

| Study | Delirium Events | Delirium Total | NoDel Events | NoDel Total | OR | 95%-CI | Weight |
|---|---|---|---|---|---|---|---|
| Bianchetti 2020 | 55 | 55 | 27 | 27 | 2.02 | [0.04; 104.45] | 0.4% |
| Callea 2022 | 9 | 20 | 4 | 51 | 9.61 | [2.50; 37.02] | 3.6% |
| Chang 2024 | 119 | 207 | 205 | 931 | 4.79 | [3.49; 6.57] | 32.8% |
| Garcia–Grimshaw 2022 | 3 | 166 | 11 | 851 | 1.41 | [0.39; 5.09] | 3.9% |
| Gutowski 2023 | 6 | 72 | 9 | 263 | 2.57 | [0.88; 7.46] | 5.6% |
| Lee 2023 | 3 | 26 | 2 | 85 | 5.41 | [0.85; 34.35] | 2.0% |
| Martins 2022 | 5 | 29 | 5 | 30 | 1.04 | [0.27; 4.06] | 3.6% |
| Mendes 2021 | 37 | 48 | 82 | 187 | 4.31 | [2.07; 8.96] | 10.8% |
| Morandi 2021 | 19 | 39 | 23 | 202 | 7.39 | [3.45; 15.86] | 10.0% |
| Ticinesi 2020 | 38 | 94 | 118 | 758 | 3.68 | [2.33; 5.81] | 21.7% |
| Tuma 2021 | 5 | 43 | 0 | 12 | 3.57 | [0.18; 69.23] | 0.8% |
| Xing 2020 | 7 | 22 | 8 | 78 | 4.08 | [1.28; 12.99] | 4.8% |
| **Random effects model** | | **821** | | **3475** | **4.13** | **[3.07; 5.55]** | **100.0%** |

Heterogeneity: $I^2 = 11\%$, $\tau^2 = 0.0294$, $p = 0.34$

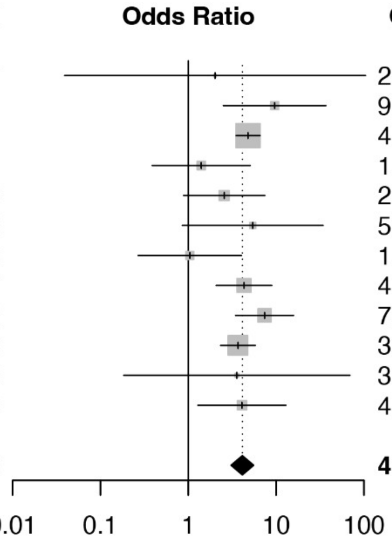


***p* <0.001***

##### B. Stroke

| Study | Delirium Events | Delirium Total | NoDel Events | NoDel Total | OR | 95%-CI | Weight |
|---|---|---|---|---|---|---|---|
| Callea 2022 | 7 | 20 | 6 | 51 | 4.04 | [1.15; 14.14] | 6.6% |
| Chang 2024 | 21 | 207 | 65 | 931 | 1.50 | [0.90; 2.52] | 39.0% |
| Damanti 2023 | 0 | 13 | 4 | 37 | 0.28 | [0.01; 5.48] | 1.2% |
| Helms 2020 | 9 | 118 | 0 | 22 | 3.90 | [0.22; 69.53] | 1.3% |
| Lee 2023 | 2 | 26 | 1 | 85 | 7.00 | [0.61; 80.55] | 1.7% |
| Mendes 2021 | 13 | 48 | 33 | 187 | 1.73 | [0.83; 3.63] | 19.0% |
| Ticinesi 2020 | 11 | 94 | 43 | 758 | 2.20 | [1.09; 4.44] | 21.2% |
| Tuma 2021 | 8 | 43 | 0 | 12 | 5.99 | [0.32; 111.44] | 1.2% |
| Xing 2020 | 8 | 22 | 10 | 78 | 3.89 | [1.30; 11.59] | 8.7% |
| **Random effects model** | | **591** | | **2161** | **2.01** | **[1.40; 2.91]** | **100.0%** |

Heterogeneity: $I^2 = 0\%$, $\tau^2 = 0$, $p = 0.49$

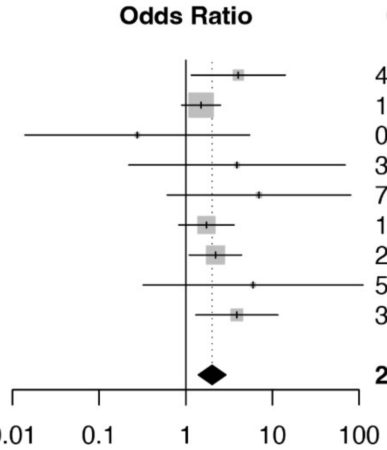


***p* = 0.002***

##### C. COPD

| Study | Delirium Events | Delirium Total | NoDel Events | NoDel Total | OR | 95%-CI | Weight |
|---|---|---|---|---|---|---|---|
| Callea 2022 | 3 | 20 | 9 | 51 | 0.82 | [0.20; 3.42] | 4.8% |
| Capuzzi 2023 | 18 | 54 | 32 | 167 | 2.11 | [1.06; 4.18] | 20.8% |
| Garcia–Grimshaw 2022 | 8 | 166 | 30 | 851 | 1.39 | [0.62; 3.08] | 15.3% |
| Gutowski 2023 | 3 | 72 | 14 | 263 | 0.77 | [0.22; 2.77] | 6.0% |
| Helms 2020 | 20 | 118 | 2 | 22 | 2.04 | [0.44; 9.43] | 4.2% |
| Lee 2023 | 2 | 26 | 4 | 85 | 1.69 | [0.29; 9.78] | 3.2% |
| Morandi 2021 | 6 | 39 | 21 | 202 | 1.57 | [0.59; 4.18] | 10.1% |
| Ticinesi 2020 | 20 | 94 | 82 | 758 | 2.23 | [1.29; 3.84] | 32.9% |
| Tuma 2021 | 5 | 43 | 1 | 12 | 1.45 | [0.15; 13.72] | 1.9% |
| Vinogradov 2021 | 1 | 10 | 0 | 20 | 6.47 | [0.24; 174.08] | 0.9% |
| **Random effects model** | | **642** | | **2431** | **1.75** | **[1.35; 2.27]** | **100.0%** |

Heterogeneity: $I^2 = 0\%$, $\tau^2 = 0$, $p = 0.86$

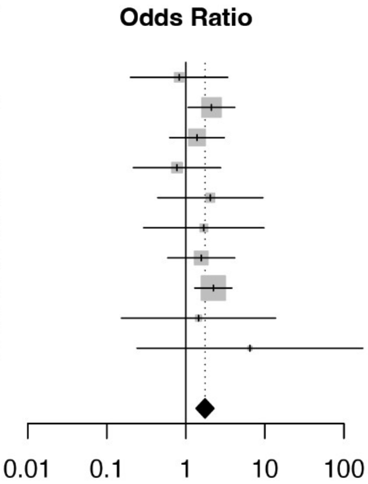


***p* = 0.001***

**Figure S12. Forest plots by comorbidities as predisposing factors of delirium.** *(continued)*

**D. Kidney disease**

| Study | Delirium Events | Delirium Total | NoDel Events | NoDel Total | OR | 95%-CI | Weight |
|---|---|---|---|---|---|---|---|
| Callea 2022 | 4 | 20 | 4 | 51 | 2.94 | [0.66; 13.13] | 5.1% |
| Chang 2024 | 20 | 207 | 74 | 931 | 1.24 | [0.74; 2.08] | 27.6% |
| Garcia–Grimshaw 2022 | 5 | 166 | 17 | 851 | 1.52 | [0.55; 4.19] | 10.3% |
| Gutowski 2023 | 13 | 72 | 30 | 263 | 1.71 | [0.84; 3.48] | 18.1% |
| Helms 2020 | 6 | 118 | 3 | 22 | 0.34 | [0.08; 1.47] | 5.3% |
| Lee 2023 | 2 | 26 | 2 | 85 | 3.46 | [0.46; 25.86] | 2.9% |
| Tasci 2022 | 8 | 95 | 0 | 59 | 11.56 | [0.65; 204.11] | 1.5% |
| Ticinesi 2020 | 16 | 94 | 59 | 758 | 2.43 | [1.33; 4.43] | 23.0% |
| Tuma 2021 | 11 | 43 | 3 | 12 | 1.03 | [0.24; 4.51] | 5.3% |
| Vinogradov 2021 | 0 | 10 | 0 | 20 | 1.95 | [0.04; 105.52] | 0.8% |
| **Random effects model** | | **851** | | **3052** | **1.62** | **[1.08; 2.43]** | **100.0%** |

Heterogeneity: $I^2 = 14\%$, $\tau^2 = 0.0477$, $p = 0.31$

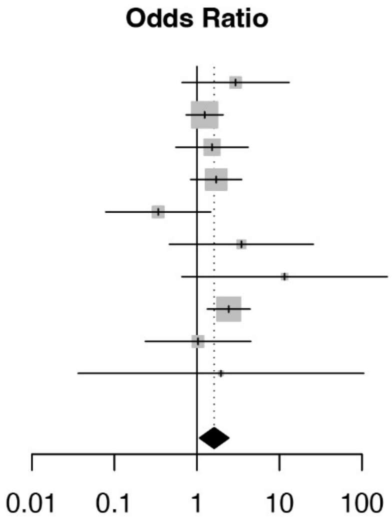


***p* <0.026***

**E. Liver disease**

| Study | Delirium Events | Delirium Total | NoDel Events | NoDel Total | OR | 95%-CI | Weight |
|---|---|---|---|---|---|---|---|
| Capuzzi 2023 | 6 | 54 | 11 | 167 | 1.77 | [0.62; 5.05] | 46.8% |
| Chang 2024 | 4 | 207 | 10 | 931 | 1.81 | [0.56; 5.84] | 37.4% |
| Damanti 2023 | 0 | 13 | 1 | 37 | 0.90 | [0.03; 23.50] | 4.8% |
| Helms 2020 | 2 | 118 | 0 | 22 | 0.97 | [0.04; 20.80] | 5.4% |
| Tasci 2022 | 2 | 95 | 0 | 59 | 3.18 | [0.15; 67.43] | 5.5% |
| **Random effects model** | | **487** | | **1216** | **1.73** | **[1.23; 2.43]** | **100.0%** |

Heterogeneity: $I^2 = 0\%$, $\tau^2 = 0$, $p = 0.98$

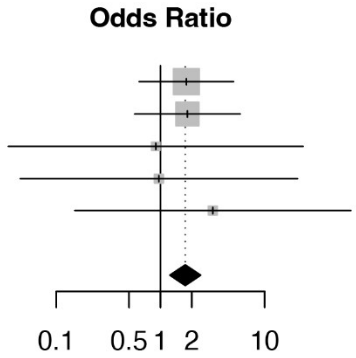


***p* = 0.011***

**F. Hypertension**

| Study | Delirium Events | Delirium Total | NoDel Events | NoDel Total | OR | 95%-CI | Weight |
|---|---|---|---|---|---|---|---|
| Callea 2022 | 12 | 20 | 25 | 51 | 1.56 | [0.55; 4.46] | 4.9% |
| Capuzzi 2023 | 33 | 54 | 83 | 167 | 1.59 | [0.85; 2.97] | 9.0% |
| Chang 2024 | 66 | 207 | 316 | 931 | 0.91 | [0.66; 1.26] | 13.6% |
| Damanti 2023 | 11 | 13 | 23 | 37 | 3.35 | [0.65; 17.37] | 2.4% |
| Garcia–Grimshaw 2022 | 54 | 166 | 250 | 851 | 1.16 | [0.81; 1.66] | 13.1% |
| Gutowski 2023 | 44 | 72 | 148 | 263 | 1.22 | [0.72; 2.08] | 10.3% |
| Lee 2023 | 21 | 26 | 34 | 85 | 6.30 | [2.17; 18.32] | 4.8% |
| Mendes 2021 | 31 | 48 | 137 | 187 | 0.67 | [0.34; 1.31] | 8.4% |
| Morandi 2021 | 25 | 39 | 118 | 202 | 1.27 | [0.62; 2.59] | 7.9% |
| Tasci 2022 | 54 | 95 | 25 | 59 | 1.79 | [0.93; 3.45] | 8.6% |
| Ticinesi 2020 | 65 | 94 | 433 | 758 | 1.68 | [1.06; 2.67] | 11.4% |
| Tuma 2021 | 31 | 43 | 8 | 12 | 1.29 | [0.33; 5.10] | 3.2% |
| Vinogradov 2021 | 7 | 10 | 8 | 20 | 3.50 | [0.69; 17.71] | 2.5% |
| **Random effects model** | | **887** | | **3623** | **1.41** | **[1.04; 1.91]** | **100.0%** |

Heterogeneity: $I^2 = 47\%$, $\tau^2 = 0.1175$, $p = 0.03$

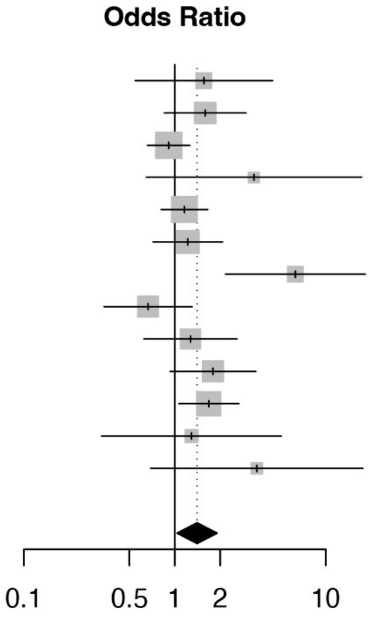


***p* = 0.032***

**Figure S12. Forest plots by comorbidities as predisposing factors of delirium.** *(continued)*

**G. Heart disease**

| Study | Delirium Events | Delirium Total | NoDel Events | NoDel Total | OR | 95%-CI | Weight |
|---|---|---|---|---|---|---|---|
| Callea 2022 | 10 | 20 | 19 | 51 | 1.68 | [0.59; 4.79] | 6.0% |
| Capuzzi 2023 | 29 | 54 | 67 | 167 | 1.73 | [0.93; 3.21] | 8.8% |
| Chang 2024 | 13 | 207 | 64 | 931 | 0.91 | [0.49; 1.68] | 8.8% |
| Damanti 2023 | 4 | 13 | 5 | 37 | 2.84 | [0.63; 12.86] | 3.9% |
| Garcia–Grimshaw 2022 | 10 | 166 | 41 | 851 | 1.27 | [0.62; 2.58] | 8.1% |
| Gutowski 2023 | 18 | 72 | 36 | 263 | 2.10 | [1.11; 3.98] | 8.7% |
| Helms 2020 | 58 | 118 | 12 | 22 | 0.81 | [0.32; 2.01] | 6.8% |
| Lee 2023 | 7 | 26 | 10 | 85 | 2.76 | [0.93; 8.21] | 5.8% |
| Mendes 2021 | 24 | 48 | 154 | 187 | 0.21 | [0.11; 0.42] | 8.4% |
| Morandi 2021 | 18 | 39 | 75 | 202 | 1.45 | [0.73; 2.90] | 8.3% |
| Tasci 2022 | 65 | 95 | 28 | 59 | 2.40 | [1.23; 4.69] | 8.4% |
| Ticinesi 2020 | 32 | 94 | 204 | 758 | 1.40 | [0.89; 2.21] | 10.0% |
| Tuma 2021 | 8 | 43 | 0 | 12 | 5.99 | [0.32; 111.44] | 1.4% |
| Xing 2020 | 12 | 22 | 40 | 78 | 1.14 | [0.44; 2.95] | 6.6% |
| **Random effects model** | | **1017** | | **3703** | **1.33** | **[0.89; 1.99]** | **100.0%** |

Heterogeneity: $I^2 = 68\%$, $\tau^2 = 0.2964$, $p < 0.01$

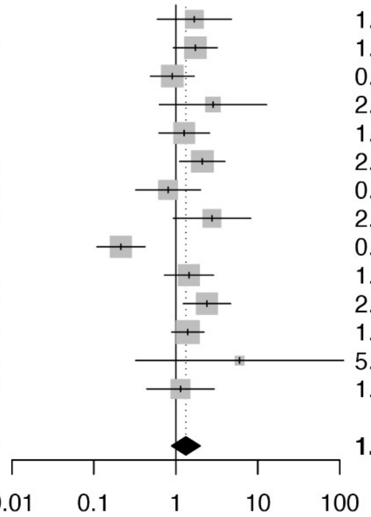


***p* = 0.15**

**H. Diabetes**

| Study | Delirium Events | Delirium Total | NoDel Events | NoDel Total | OR | 95%-CI | Weight |
|---|---|---|---|---|---|---|---|
| Callea 2022 | 7 | 20 | 14 | 51 | 1.42 | [0.47; 4.30] | 3.2% |
| Capuzzi 2023 | 11 | 54 | 37 | 167 | 0.90 | [0.42; 1.91] | 6.9% |
| Damanti 2023 | 2 | 13 | 11 | 37 | 0.43 | [0.08; 2.27] | 1.4% |
| Garcia–Grimshaw 2022 | 45 | 166 | 233 | 851 | 0.99 | [0.68; 1.43] | 28.4% |
| Gutowski 2023 | 22 | 72 | 63 | 263 | 1.40 | [0.79; 2.48] | 12.0% |
| Helms 2020 | 18 | 118 | 3 | 22 | 1.14 | [0.31; 4.25] | 2.3% |
| Lee 2023 | 7 | 26 | 22 | 85 | 1.06 | [0.39; 2.85] | 4.0% |
| Mendes 2021 | 9 | 48 | 45 | 187 | 0.73 | [0.33; 1.62] | 6.2% |
| Morandi 2021 | 14 | 39 | 43 | 202 | 2.07 | [0.99; 4.32] | 7.3% |
| Tasci 2022 | 28 | 95 | 11 | 59 | 1.82 | [0.83; 4.02] | 6.4% |
| Ticinesi 2020 | 23 | 94 | 156 | 758 | 1.25 | [0.76; 2.07] | 15.8% |
| Tuma 2021 | 19 | 43 | 4 | 12 | 1.58 | [0.41; 6.06] | 2.2% |
| Vinogradov 2021 | 2 | 10 | 3 | 20 | 1.42 | [0.20; 10.23] | 1.0% |
| Xing 2020 | 5 | 22 | 10 | 78 | 2.00 | [0.60; 6.63] | 2.8% |
| **Random effects model** | | **820** | | **2792** | **1.19** | **[0.99; 1.43]** | **100.0%** |

Heterogeneity: $I^2 = 0\%$, $\tau^2 = 0$, $p = 0.76$

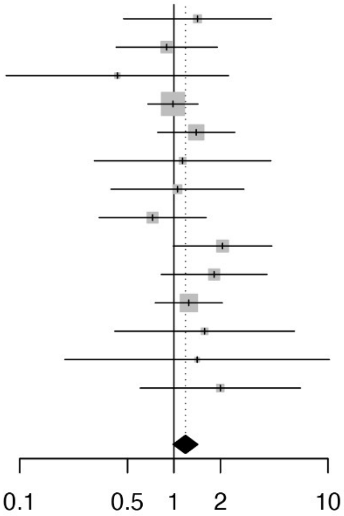


***p* = 0.06**

**i. Cancer**

| Study | Delirium Events | Delirium Total | NoDel Events | NoDel Total | OR | 95%-CI | Weight |
|---|---|---|---|---|---|---|---|
| Callea 2022 | 6 | 20 | 5 | 51 | 3.94 | [1.04; 14.89] | 10.4% |
| Capuzzi 2023 | 12 | 54 | 19 | 167 | 2.23 | [1.00; 4.95] | 15.9% |
| Chang 2024 | 14 | 207 | 43 | 931 | 1.50 | [0.80; 2.79] | 18.0% |
| Damanti 2023 | 0 | 13 | 10 | 37 | 0.10 | [0.01; 1.78] | 3.4% |
| Gutowski 2023 | 7 | 72 | 37 | 263 | 0.66 | [0.28; 1.54] | 15.2% |
| Helms 2020 | 16 | 118 | 5 | 22 | 0.53 | [0.17; 1.65] | 12.3% |
| Ticinesi 2020 | 11 | 94 | 119 | 758 | 0.71 | [0.37; 1.38] | 17.6% |
| Tuma 2021 | 5 | 43 | 2 | 12 | 0.66 | [0.11; 3.91] | 7.3% |
| **Random effects model** | | **621** | | **2241** | **1.03** | **[0.51; 2.09]** | **100.0%** |

Heterogeneity: $I^2 = 54\%$, $\tau^2 = 0.3910$, $p = 0.03$

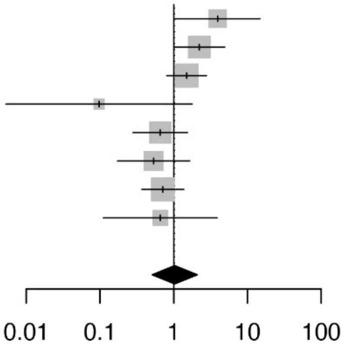


***p* = 0.91**

**Figure S13. Forest plots by demographics as predisposing factors of delirium in studies at low risk of bias**

**A. Gender (Female)**

| Study | Delirium Events | Delirium Total | No Del Events | No Del Total | OR | 95%−CI | Weight |
|---|---|---|---|---|---|---|---|
| Callea 2022 | 9 | 20 | 21 | 51 | 1.17 | [0.41; 3.32] | 10.2% |
| D'Ardes 2021 | 9 | 14 | 28 | 42 | 0.90 | [0.25; 3.20] | 7.0% |
| Helms 2020 | 29 | 118 | 11 | 22 | 0.33 | [0.13; 0.83] | 12.5% |
| Mendes 2021 | 24 | 48 | 109 | 187 | 0.72 | [0.38; 1.35] | 24.7% |
| Ticinesi 2020 | 42 | 94 | 360 | 758 | 0.89 | [0.58; 1.37] | 45.6% |
| **Random effects model** | | **294** | | **1060** | **0.77** | **[0.47; 1.25]** | **100.0%** |

***p* = 0.20**

Heterogeneity: $I^2 = 9.6\%$, $\tau^2 = 0.0190$, $p = 0.3516$

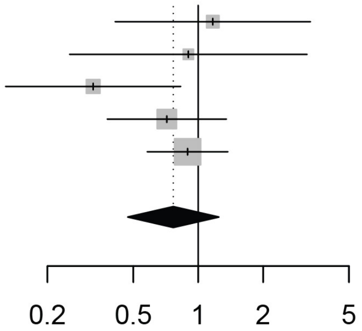


**B. Age difference**

| Study | DOM | 95% CI | Weight |
|---|---|---|---|
| Ticinesi 2020 | 9.00 | [ 6.99, 11.01] | 24.14% |
| Mendes 2021 | 0.80 | [−1.05, 2.65] | 24.30% |
| Helms 2020 | 0.33 | [−7.71, 8.37] | 14.56% |
| D'Ardes 2021 | 13.31 | [ 7.36, 19.26] | 18.01% |
| Callea 2022 | 7.33 | [ 1.95, 12.72] | 18.99% |
| **Random−Effects Model** | **6.20** | **[ 1.49, 10.92]** | **100%** |

***p* = 0.01***

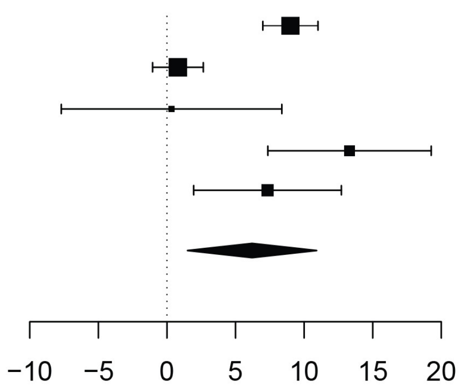


**C. Nursing home**

| Study | Delirium Events | Delirium Total | No Del Events | No Del Total | OR | 95%−CI |
|---|---|---|---|---|---|---|
| Mendes 2021 | 16 | 48 | 33 | 187 | 2.33 | [1.15; 4.74] |

***p* = 0.02***

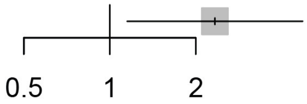

**Figure S14. Forest plots by comorbidities as predisposing factors of delirium in studies at low risk of bias**

**A. Dementia**

| Study | Delirium Events | Delirium Total | No Del Events | No Del Total | OR | 95%−CI | Weight |
|---|---|---|---|---|---|---|---|
| Callea 2022 | 9 | 20 | 4 | 51 | 9.61 | [2.50; 37.02] | 7.6% |
| Mendes 2021 | 37 | 48 | 82 | 187 | 4.31 | [2.07; 8.96] | 25.8% |
| Ticinesi 2020 | 38 | 94 | 118 | 758 | 3.68 | [2.33; 5.81] | 66.5% |
| **Random effects model** | | **162** | | **996** | **4.12** | **[1.91; 8.89]** | **100.0%** |
| | | | | | | ***p* = 0.015*** | |

Heterogeneity: $I^2 = 0.0\%$, $\tau^2 = 0$, $p = 0.4136$

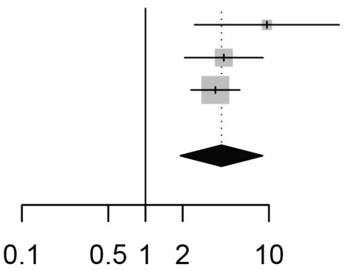


**B. Stroke**

| Study | Delirium Events | Delirium Total | No Del Events | No Del Total | OR | 95%−CI | Weight |
|---|---|---|---|---|---|---|---|
| Callea 2022 | 7 | 20 | 6 | 51 | 4.04 | [1.15; 14.14] | 13.8% |
| Helms 2020 | 9 | 118 | 0 | 22 | 3.90 | [0.22; 69.53] | 2.6% |
| Mendes 2021 | 13 | 48 | 33 | 187 | 1.73 | [0.83; 3.63] | 39.5% |
| Ticinesi 2020 | 11 | 94 | 43 | 758 | 2.20 | [1.09; 4.44] | 44.1% |
| **Random effects model** | | **280** | | **1018** | **2.21** | **[1.31; 3.74]** | **100.0%** |
| | | | | | | ***p* = 0.017*** | |

Heterogeneity: $I^2 = 0.0\%$, $\tau^2 = 0$, $p = 0.6929$

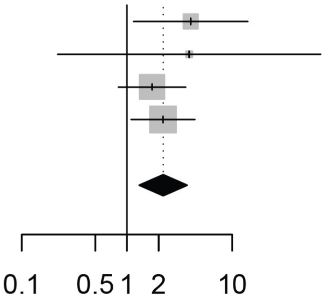


**C. COPD**

| Study | Delirium Events | Delirium Total | No Del Events | No Del Total | OR | 95%−CI | Weight |
|---|---|---|---|---|---|---|---|
| Callea 2022 | 3 | 20 | 9 | 51 | 0.82 | [0.20; 3.42] | 11.5% |
| Helms 2020 | 20 | 118 | 2 | 22 | 2.04 | [0.44; 9.43] | 9.9% |
| Ticinesi 2020 | 20 | 94 | 82 | 758 | 2.23 | [1.29; 3.84] | 78.6% |
| **Random effects model** | | **232** | | **831** | **1.97** | **[0.75; 5.14]** | **100.0%** |
| | | | | | | ***p* = 0.093** | |

Heterogeneity: $I^2 = 0.0\%$, $\tau^2 = 0$, $p = 0.4401$

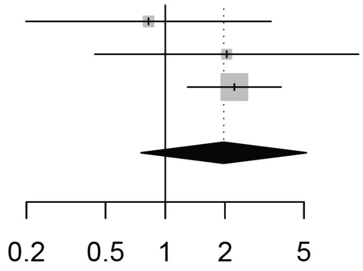


**D. Kidney disease**

| Study | Delirium Events | Delirium Total | No Del Events | No Del Total | OR | 95%−CI | Weight |
|---|---|---|---|---|---|---|---|
| Callea 2022 | 4 | 20 | 4 | 51 | 2.94 | [0.66; 13.13] | 28.6% |
| Helms 2020 | 6 | 118 | 3 | 22 | 0.34 | [0.08; 1.47] | 29.1% |
| Ticinesi 2020 | 16 | 94 | 59 | 758 | 2.43 | [1.33; 4.43] | 42.3% |
| **Random effects model** | | **232** | | **831** | **1.45** | **[0.08; 24.67]** | **100.0%** |
| | | | | | | ***p* = 0.631** | |

Heterogeneity: $I^2 = 68.0\%$, $\tau^2 = 0.9330$, $p = 0.0439$

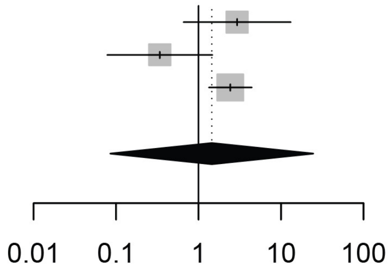

**Figure S14. Forest plots by comorbidities as predisposing factors of delirium in studies at low risk of bias.** *(continued)*

**E. Hypertension**

| Study | Delirium Events | Delirium Total | No Del Events | No Del Total | OR | 95%−CI | Weight |
|---|---|---|---|---|---|---|---|
| Callea 2022 | 12 | 20 | 25 | 51 | 1.56 | [0.55; 4.46] | 21.1% |
| Mendes 2021 | 31 | 48 | 137 | 187 | 0.67 | [0.34; 1.31] | 34.2% |
| Ticinesi 2020 | 65 | 94 | 433 | 758 | 1.68 | [1.06; 2.67] | 44.7% |
| **Random effects model** | | **162** | | **996** | **1.21** | **[0.33; 4.45]** | **100.0%** |

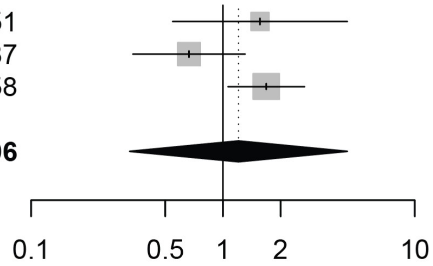


Heterogeneity: $I^2 = 60.8\%$, $\tau^2 = 0.1511$, $p = 0.0780$

***p* = 0.60**

**F. Heart disease**

| Study | Delirium Events | Delirium Total | No Del Events | No Del Total | OR | 95%−CI | Weight |
|---|---|---|---|---|---|---|---|
| Callea 2022 | 10 | 20 | 19 | 51 | 1.68 | [0.59; 4.79] | 21.9% |
| Helms 2020 | 58 | 118 | 12 | 22 | 0.81 | [0.32; 2.01] | 23.5% |
| Mendes 2021 | 24 | 48 | 154 | 187 | 0.21 | [0.11; 0.42] | 26.2% |
| Ticinesi 2020 | 32 | 94 | 204 | 758 | 1.40 | [0.89; 2.21] | 28.4% |
| **Random effects model** | | **280** | | **1018** | **0.78** | **[0.18; 3.51]** | **100.0%** |

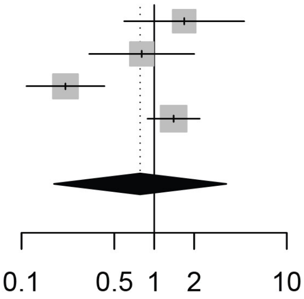


Heterogeneity: $I^2 = 86.4\%$, $\tau^2 = 0.7262$, $p < 0.0001$

***p* = 0.64**

**G. Diabetes**

| Study | Delirium Events | Delirium Total | No Del Events | No Del Total | OR | 95%−CI | Weight |
|---|---|---|---|---|---|---|---|
| Callea 2022 | 7 | 20 | 14 | 51 | 1.42 | [0.47; 4.30] | 11.8% |
| Helms 2020 | 18 | 118 | 3 | 22 | 1.14 | [0.31; 4.25] | 8.3% |
| Mendes 2021 | 9 | 48 | 45 | 187 | 0.73 | [0.33; 1.62] | 22.6% |
| Ticinesi 2020 | 23 | 94 | 156 | 758 | 1.25 | [0.76; 2.07] | 57.2% |
| **Random effects model** | | **280** | | **1018** | **1.11** | **[0.72; 1.72]** | **100.0%** |

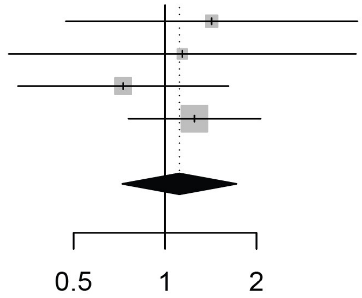


Heterogeneity: $I^2 = 0.0\%$, $\tau^2 = 0$, $p = 0.6867$

***p* = 0.48**

**H. Cancer**

| Study | Delirium Events | Delirium Total | No Del Events | No Del Total | OR | 95%−CI | Weight |
|---|---|---|---|---|---|---|---|
| Callea 2022 | 6 | 20 | 5 | 51 | 3.94 | [1.04; 14.89] | 28.5% |
| Helms 2020 | 16 | 118 | 5 | 22 | 0.53 | [0.17; 1.65] | 31.8% |
| Ticinesi 2020 | 11 | 94 | 119 | 758 | 0.71 | [0.37; 1.38] | 39.6% |
| **Random effects model** | | **232** | | **831** | **1.06** | **[0.08; 13.62]** | **100.0%** |

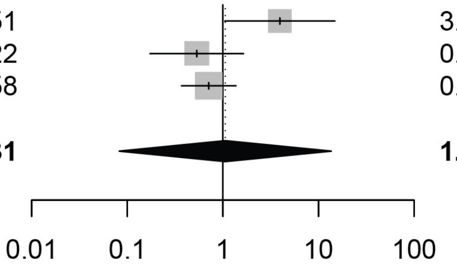


Heterogeneity: $I^2 = 66.9\%$, $\tau^2 = 0.7766$, $p = 0.0486$

***p* = 0.93**

#### Figure S15. Forest plots by pneumonia-severity factors

##### A. ICU Admission

| Study | Delirium Events | Delirium Total | NoDel Events | NoDel Total | OR | 95%−CI | Weight |
|---|---|---|---|---|---|---|---|
| Carr 2019 | 73 | 73 | 29 | 29 | 2.49 | [0.05; 128.51] | 1.8% |
| Cooper 2020 | 11 | 11 | 16 | 16 | 0.70 | [0.01; 37.73] | 1.8% |
| Denke 2018 | 37 | 37 | 5 | 5 | 6.82 | [0.12; 380.08] | 1.8% |
| Eggers 2004 | 11 | 11 | 9 | 9 | 1.21 | [0.02; 66.96] | 1.8% |
| Ekmekyapar 2022 | 64 | 64 | 486 | 486 | 0.13 | [0.00; 6.74] | 1.9% |
| Gholi 2022 | 217 | 217 | 93 | 93 | 2.33 | [0.05; 118.11] | 1.9% |
| GomezDuque 2023 | 148 | 148 | 135 | 135 | 1.10 | [0.02; 55.61] | 1.9% |
| Gutowski 2023 | 8 | 72 | 63 | 263 | 0.40 | [0.18; 0.87] | 19.1% |
| Helms 2020 | 118 | 118 | 22 | 22 | 5.27 | [0.10; 272.37] | 1.8% |
| Iribarren−Diarasarri 2023 | 63 | 63 | 80 | 80 | 0.79 | [0.02; 40.31] | 1.8% |
| Kaneko 2022 | 24 | 24 | 88 | 88 | 0.28 | [0.01; 14.31] | 1.8% |
| Lee 2023 | 26 | 26 | 85 | 85 | 0.31 | [0.01; 16.00] | 1.8% |
| Limpawattana 2016 | 21 | 21 | 15 | 15 | 1.39 | [0.03; 73.79] | 1.8% |
| Lin 2008 | 14 | 14 | 35 | 35 | 0.41 | [0.01; 21.58] | 1.8% |
| Martins 2022 | 29 | 29 | 30 | 30 | 0.97 | [0.02; 50.36] | 1.8% |
| Portela−Sanchez 2021 | 3 | 13 | 10 | 58 | 1.44 | [0.33; 6.19] | 9.8% |
| Premkumar 2019 | 67 | 67 | 43 | 43 | 1.55 | [0.03; 79.66] | 1.8% |
| Regueiro−Mira 2015 | 6 | 17 | 33 | 223 | 3.14 | [1.09; 9.08] | 14.5% |
| Tomasi 2017 | 4 | 10 | 0 | 20 | 28.38 | [1.34; 600.58] | 3.0% |
| vanderKnaap 2024 | 48 | 48 | 276 | 276 | 0.18 | [0.00; 8.95] | 1.8% |
| Yuksel 2021 | 163 | 213 | 32 | 94 | 6.32 | [3.71; 10.75] | 24.3% |
| **Random effects model** | | **1296** | | **2105** | **1.73** | **[0.96; 3.10]** | **100.0%** |

Heterogeneity: $I^2 = 55.0\%$, $\tau^2 = 0.2518$, $p = 0.0013$

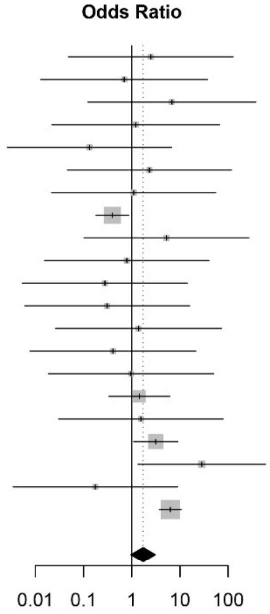


***p* = 0.07**

##### B. Invasive Ventilation

| Study | Delirium Events | Delirium Total | NoDel Events | NoDel Total | OR | 95%−CI | Weight |
|---|---|---|---|---|---|---|---|
| Carr 2019 | 73 | 73 | 29 | 29 | 2.49 | [ 0.05; 128.51] | 3.3% |
| Damanti 2023 | 0 | 13 | 0 | 37 | 2.78 | [ 0.05; 147.03] | 3.2% |
| Eggers 2004 | 11 | 11 | 9 | 9 | 1.21 | [ 0.02; 66.96] | 3.2% |
| Garcia−Grimshaw 2022 | 131 | 166 | 116 | 851 | 23.72 | [15.56; 36.14] | 14.2% |
| GomezDuque 2023 | 148 | 148 | 135 | 135 | 1.10 | [ 0.02; 55.61] | 3.3% |
| Helms 2020 | 118 | 118 | 22 | 22 | 5.27 | [ 0.10; 272.37] | 3.3% |
| Iribarren−Diarasarri 2023 | 63 | 63 | 80 | 80 | 0.79 | [ 0.02; 40.31] | 3.3% |
| Kaneko 2022 | 24 | 24 | 88 | 88 | 0.28 | [ 0.01; 14.31] | 3.3% |
| Lee 2023 | 22 | 26 | 10 | 85 | 41.25 | [11.78; 144.43] | 10.9% |
| Lin 2008 | 14 | 14 | 35 | 35 | 0.41 | [ 0.01; 21.58] | 3.2% |
| Martins 2022 | 26 | 29 | 18 | 30 | 5.78 | [ 1.42; 23.44] | 10.2% |
| Tasci 2022 | 38 | 95 | 18 | 59 | 1.52 | [ 0.76; 3.03] | 13.4% |
| Tuma 2021 | 19 | 43 | 5 | 12 | 1.11 | [ 0.30; 4.05] | 10.7% |
| vanderKnaap 2024 | 48 | 48 | 276 | 276 | 0.18 | [ 0.00; 8.95] | 3.3% |
| Yuksel 2021 | 52 | 213 | 3 | 94 | 9.80 | [ 2.98; 32.26] | 11.2% |
| **Random effects model** | | **1084** | | **1842** | **3.86** | **[ 1.60; 9.33]** | **100.0%** |

Heterogeneity: $I^2 = 81\%$, $\tau^2 = 1.1436$, $p < 0.01$

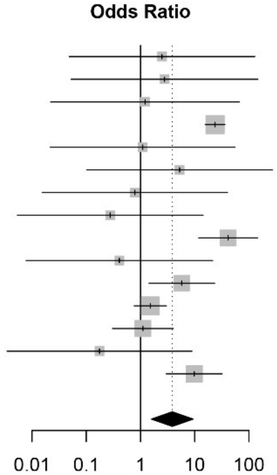


***p* = 0.005***

##### C. Noninvasive Ventilation

| Study | Delirium Events | Delirium Total | NoDel Events | NoDel Total | OR | 95%−CI | Weight |
|---|---|---|---|---|---|---|---|
| Callea 2022 | 9 | 20 | 9 | 51 | 3.82 | [1.22; 11.91] | 8.9% |
| Carr 2019 | 0 | 73 | 0 | 29 | 0.40 | [0.01; 20.70] | 2.9% |
| Damanti 2023 | 0 | 13 | 4 | 37 | 0.28 | [0.01; 5.48] | 4.2% |
| Eggers 2004 | 0 | 11 | 0 | 9 | 0.83 | [0.01; 45.69] | 2.8% |
| Garcia−Grimshaw 2022 | 35 | 166 | 735 | 851 | 0.04 | [0.03; 0.06] | 10.6% |
| GomezDuque 2023 | 0 | 148 | 0 | 135 | 0.91 | [0.02; 46.30] | 2.9% |
| Helms 2020 | 0 | 118 | 0 | 22 | 0.19 | [0.00; 9.82] | 2.9% |
| Iribarren−Diarasarri 2023 | 0 | 63 | 0 | 80 | 1.27 | [0.02; 64.78] | 2.9% |
| Kaneko 2022 | 0 | 24 | 0 | 88 | 3.61 | [0.07; 186.74] | 2.9% |
| Lee 2023 | 4 | 26 | 75 | 85 | 0.02 | [0.01; 0.08] | 8.5% |
| Lin 2008 | 0 | 14 | 0 | 35 | 2.45 | [0.05; 129.36] | 2.9% |
| Martins 2022 | 3 | 29 | 12 | 30 | 0.17 | [0.04; 0.70] | 8.1% |
| Morandi 2021 | 5 | 39 | 25 | 202 | 1.04 | [0.37; 2.91] | 9.2% |
| Tasci 2022 | 57 | 95 | 41 | 59 | 0.66 | [0.33; 1.31] | 10.1% |
| Tuma 2021 | 24 | 43 | 7 | 12 | 0.90 | [0.25; 3.30] | 8.4% |
| vanderKnaap 2024 | 0 | 48 | 0 | 276 | 5.70 | [0.11; 290.74] | 2.9% |
| Yuksel 2021 | 161 | 213 | 91 | 94 | 0.10 | [0.03; 0.34] | 8.7% |
| **Random effects model** | | **1143** | | **2095** | **0.39** | **[0.17; 0.92]** | **100.0%** |

Heterogeneity: $I^2 = 87\%$, $\tau^2 = 1.4799$, $p < 0.01$

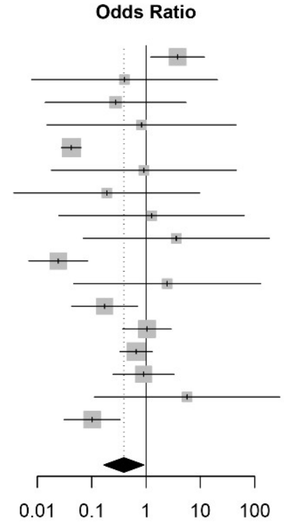


***p* = 0.03***

#### Figure S15. Forest plots by pneumonia-severity factors. *(continued)*

**D. Multilobar pneumonia**

| Study | Delirium Events | Delirium Total | NoDel Events | NoDel Total | OR | 95%–CI | Weight |
|---|---|---|---|---|---|---|---|
| Garcia–Grimshaw 2022 | 129 | 166 | 425 | 851 | 3.49 | [2.37; 5.16] | 26.0% |
| Morandi 2021 | 10 | 39 | 64 | 202 | 0.74 | [0.34; 1.62] | 17.0% |
| Portela–Sanchez 2021 | 11 | 13 | 45 | 58 | 1.59 | [0.31; 8.09] | 6.6% |
| Trevisan 2023 | 158 | 197 | 717 | 963 | 1.39 | [0.95; 2.03] | 26.2% |
| Vinogradov 2021 | 10 | 10 | 20 | 20 | 0.51 | [0.01; 27.68] | 1.3% |
| Yuksel 2021 | 153 | 213 | 62 | 94 | 1.32 | [0.78; 2.22] | 22.8% |
| **Random effects model** | | **638** | | **2188** | **1.56** | **[0.84; 2.90]** | **100.0%** |

Heterogeneity: $I^2 = 75\%$, $\tau^2 = 0.1829$, $p < 0.01$

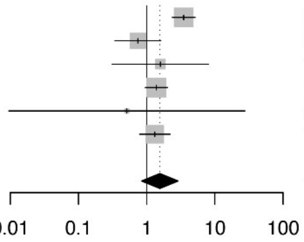


***p* = 0.12**

**E. Dialysis**

| Study | Delirium Events | Delirium Total | NoDel Events | NoDel Total | OR | 95%–CI | Weight |
|---|---|---|---|---|---|---|---|
| Garcia–Grimshaw 2022 | 43 | 166 | 64 | 851 | 4.30 | [2.79; 6.61] | 89.2% |
| Lee 2023 | 3 | 26 | 2 | 85 | 5.41 | [0.85; 34.35] | 4.8% |
| Tuma 2021 | 11 | 43 | 2 | 12 | 1.72 | [0.32; 9.09] | 6.0% |
| **Random effects model** | | **235** | | **948** | **4.12** | **[2.07; 8.17]** | **100.0%** |

Heterogeneity: $I^2 = 0\%$, $\tau^2 = 0$, $p = 0.55$

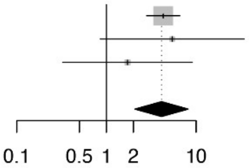


***p* = 0.01***

**F. Steroids**

| Study | Delirium Events | Delirium Total | NoDel Events | NoDel Total | OR | 95%–CI | Weight |
|---|---|---|---|---|---|---|---|
| Callea 2022 | 20 | 20 | 49 | 51 | 2.07 | [0.10; 45.04] | 10.1% |
| Chang 2024 | 33 | 207 | 199 | 931 | 0.70 | [0.47; 1.04] | 55.3% |
| Damanti 2023 | 3 | 13 | 27 | 37 | 0.11 | [0.03; 0.49] | 28.0% |
| Lee 2023 | 26 | 26 | 85 | 85 | 0.31 | [0.01; 16.00] | 6.6% |
| **Random effects model** | | **266** | | **1104** | **0.44** | **[0.08; 2.52]** | **100.0%** |

Heterogeneity: $I^2 = 52\%$, $\tau^2 = 0.4985$, $p = 0.10$

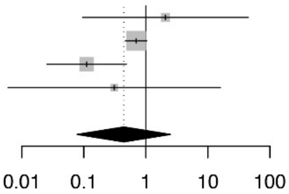


***p* = 0.23**

**Figure S16. Forest plots by pneumonia-severity factors in studies at low risk of bias.**

**A. ICU Admision**

| Study | Delirium Events | Delirium Total | No Del Events | No Del Total | OR | 95%−CI | Weight |
|---|---|---|---|---|---|---|---|
| Gholi 2022 | 217 | 217 | 93 | 93 | 2.33 | [0.05; 118.11] | 33.5% |
| Helms 2020 | 118 | 118 | 22 | 22 | 5.27 | [0.10; 272.37] | 33.2% |
| Premkumar 2019 | 67 | 67 | 43 | 43 | 1.55 | [0.03; 79.66] | 33.3% |
| **Random effects model** | | **402** | | **158** | **2.67** | **[0.57; 12.49]** | **100.0%** |

Heterogeneity: $I^2 = 0.0\%$, $\tau^2 = 0$, $p = 0.9087$

***p* = 0.11**

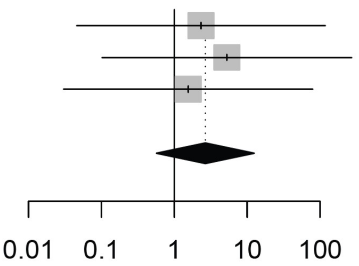


**B. Invasive ventilation**

| Study | Delirium Events | Delirium Total | No Del Events | No Del Total | OR | 95%−CI |
|---|---|---|---|---|---|---|
| Helms 2020 | 118 | 118 | 22 | 22 | 5.27 | [0.10; 272.37] |

***p* = 0.41**

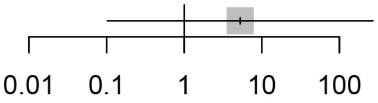


**C. Steroids**

| Study | Delirium Events | Delirium Total | No Del Events | No Del Total | OR | 95%−CI |
|---|---|---|---|---|---|---|
| Callea 2022 | 20 | 20 | 49 | 51 | 2.07 | [0.10; 45.04] |

***p* = 0.63**

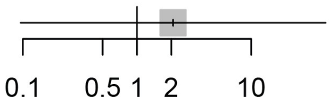

#### Figure S17. Forest plots of the associations between delirium and clinical course

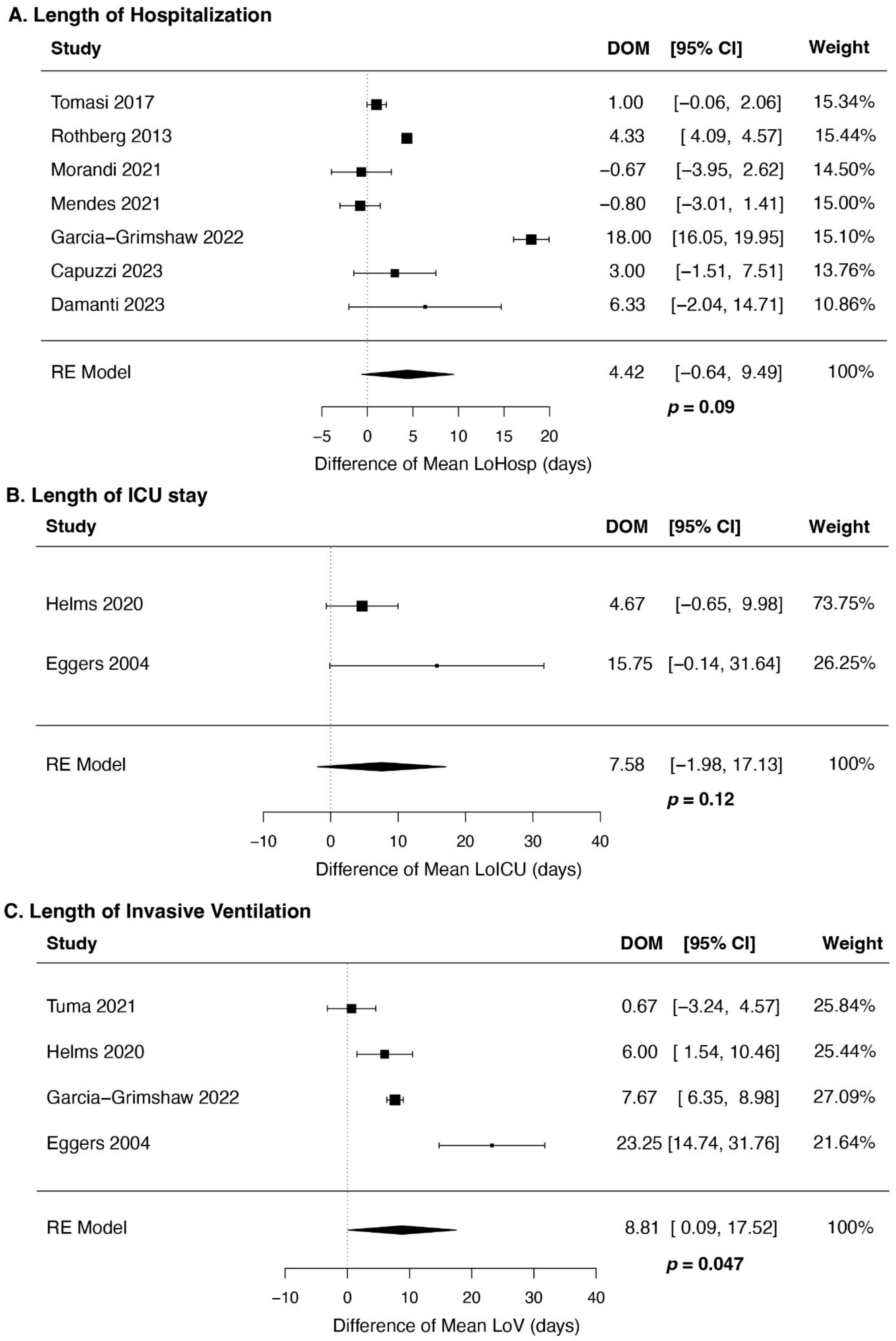


**A.** Patients with pneumonia and delirium did not show significant differences in hospitalization length or in ICU stay (**B**), but they did show increased length of invasive ventilation (**C**, DOM 8.81, 95% CI [0.09, 17.52], *p*=0.047). Difference of means (DOM) was calculated using the Hozo/Wan/Bland method.

**Figure S18. Forest plots of the associations between delirium and clinical course in studies at low risk of bias**

**A. Length of Hospitalization**

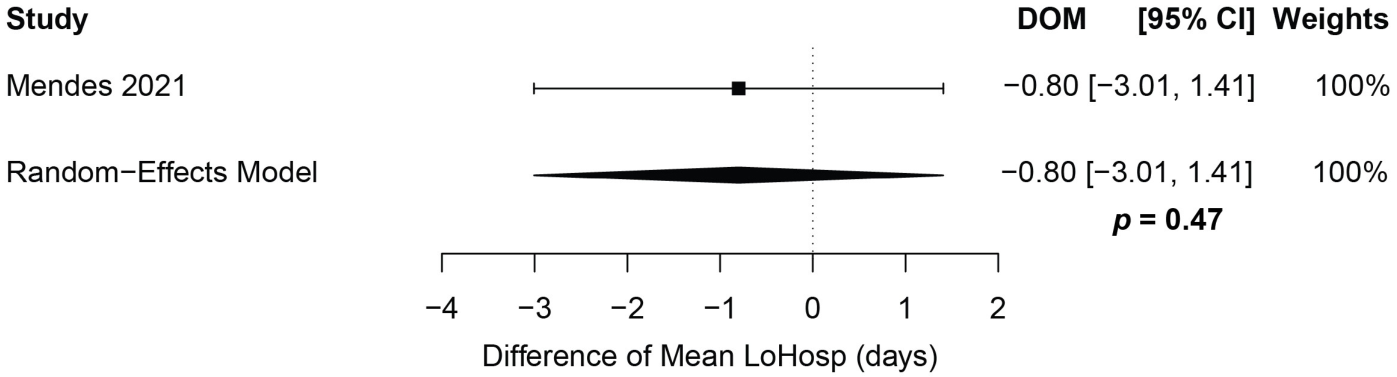


**B. Length of ICU stay**

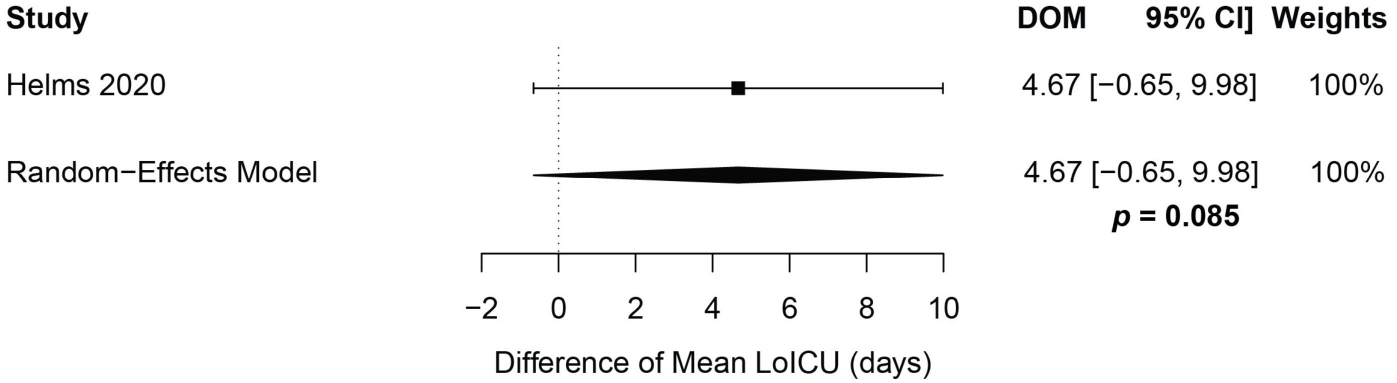


**C. Length of Invasive Ventilation**

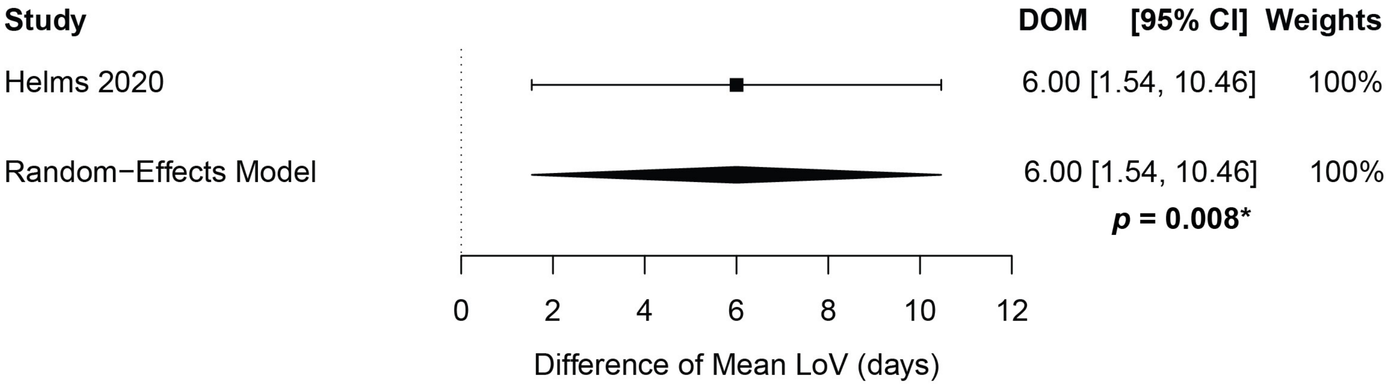

**Figure S19. Delirium is associated with significantly increased mortality in patients with pneumonia in studies at low risk of bias.**

**A. Overall death univariate**

| Study | Delirium Events | Delirium Total | No Del Events | No Del Total | OR | 95%−CI | Weight |
|---|---|---|---|---|---|---|---|
| Aliberti 2015 | 45 | 53 | 22 | 119 | 24.80 | [10.26; 59.98] | 17.1% |
| Callea 2022 | 7 | 20 | 3 | 51 | 8.62 | [ 1.95; 38.03] | 13.2% |
| Helms 2020 | 19 | 118 | 2 | 22 | 1.92 | [ 0.41; 8.90] | 12.9% |
| Mendes 2021 | 20 | 48 | 56 | 187 | 1.67 | [ 0.87; 3.21] | 18.5% |
| Pieralli 2014 | 17 | 110 | 37 | 333 | 1.46 | [ 0.79; 2.72] | 18.7% |
| Ticinesi 2020 | 54 | 94 | 227 | 758 | 3.16 | [ 2.04; 4.89] | 19.6% |
| **Random effects model** | | **443** | | **1470** | **3.70** | **[ 1.14; 12.05]** | **100.0%** |

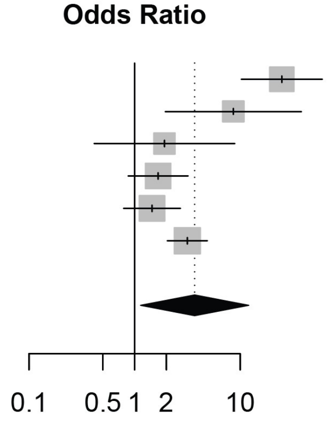


Heterogeneity: $I^2 = 84.6\%$, $\tau^2 = 1.0258$, $p < 0.0001$

***p* = 0.036***

**B. Overall death multivariate**

| Study | logOR | SE(logOR) | OR | 95%−CI | Weight |
|---|---|---|---|---|---|
| Aliberti 2015 | 0.8544 | 0.3742 | 2.35 | [1.13; 4.90] | 25.3% |
| Callea 2022 | 1.9592 | 0.9898 | 7.09 | [1.02; 49.38] | 3.6% |
| Mendes 2021 | 0.7419 | 0.2873 | 2.10 | [1.20; 3.70] | 43.0% |
| Pieralli 2014 | 1.7387 | 0.7237 | 5.69 | [1.36; 23.20] | 6.8% |
| Ticinesi 2020 | 0.2078 | 0.4076 | 1.23 | [0.55; 2.74] | 21.3% |
| **Random effects model (HK)** | | | **2.16** | **[1.19; 3.91]** | **100.0%** |

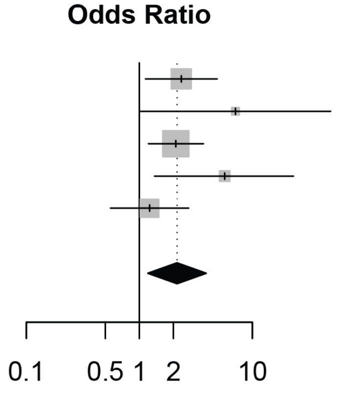


Heterogeneity: $I^2 = 23.0\%$, $\tau^2 < 0.0001$, $p = 0.2676$

***p* = 0.023***
